# Differential effects of glucagon-like peptide-1 receptor agonist classes on blood pressure: a systematic review and network meta-analysis of randomised controlled trials with meta-regression

**DOI:** 10.1101/2025.07.05.25330933

**Authors:** Oscar Hou In Chou, Haixin Zhou, Hamza Waraich, Derek Wu, Keanu Razzaghi, Jeffrey Shi Kai Chan, Tong Liu, Bernard Man Yung Cheung, Gary Tse, Carmel M McEniery, Ian B Wilkinson

## Abstract

**Background and Aims:** Recent clinical trials have reported blood pressure (BP)-lowering effects of glucagon-like peptide-1 receptor agonists (GLP1Ra). A recent systematic review has focused on the effects of semaglutide. However, there has been no comprehensive evaluation of the BP effects of all GLP1Ra available, including the double agonist tirzepatide and the triple agonist retatrutide. Additionally, the extent to which BP reduction is mediated by weight loss remains unclear. This systematic review and network meta-analysis aimed to evaluate the impact of GLP1Ra on systolic and diastolic BP across randomized controlled trials (RCTs).

**Methods:** PubMed/MEDLINE, Web of Science and Ovid/Embase were searched from their inception until 31^st^ July 2024. RCTs involving adult patients treated with GLP1Ra that reported BP and weight changes were included. Pair-wise meta-analysis and meta-regression models were utilised. Network meta-analysis was conducted. Mean difference (MD) and its 95% confidence intervals (CIs) were reported.

**Results:** A total of 75 RCTs, including 114352 participants, were included. Retatrutide demonstrated the greatest reduction in systolic BP (MD: −7.0 mmHg; 95% CI: −10.5 to −3.5, followed by tirzepatide (MD: −5.2 mmHg; 95% CI: −6.9 to −3.5) and semaglutide (MD: −3.4 mmHg; 95% CI: −4.7 to −2.1). For diastolic BP, tirzepatide showed the largest reduction (MD: −1.7 mmHg; 95% CI: −2.6 to −0.8), followed by semaglutide (MD: −0.8 mmHg; 95% CI: −1.4 to −0.2). Mediation analysis indicated that weight loss partially mediated the BP-lowering effects of GLP1Ra.

**Conclusion:** Retatrutide, tirzepatide and semaglutide reduced systolic blood pressure compared to placebo. Tirzepatide and semaglutide also led to significant diastolic BP reductions. The triple agonist retatrutide emerged as the most effective agent for lowering systolic BP among all GLP1Ra classes.

## Introduction

Hypertension, affecting over 1.3 billion people worldwide, remains a leading cause of adverse cardiovascular events, contributing to significant morbidity, mortality, and economic burden.^1^ Type 2 diabetes mellitus (T2DM) is an established risk factor for cardiovascular disease and is associated with concomitant hypertension.^2^ Obesity is likely the commonest modifiable risk factor for both hypertension and T2DM.^3^ Despite the numerous anti-hypertensive agents available, achieving optimal blood pressure targets remains a major challenge. Glucagon-like peptide-1 receptor agonists (GLP1Ra) have emerged as a promising therapeutic option, not only for their glucose-lowering and weight-reducing effects but also for their potential to address multiple cardiovascular risk factors, including hypertension.^4^ GLP1 is an incretin hormone that triggers the release of insulin from the pancreas. GLP1Ra has also demonstrated substantial cardiovascular and renal protective effects in multiple randomised controlled trials (RCTs) for primary and secondary prevention.^5^

GLP1Ra appear to reduce systolic blood pressure (SBP) but their effects on diastolic blood pressure (DBP) is less consistent.^6,7^ The mechanisms by which GLP1Ra lowers BP are unknown but besides weight loss *per se*, GLP1Ra may achieve it via natriuresis, inhibition of the renin-angiotensin (RAAS) and sympathetic nervous systems,^8^ and vasodilatation^9.^ Given the diverse efficacy and dosing requirements for GLP1Ra, the advantages and disadvantages of each GLP1Ra drug in terms of cardiovascular risks and blood pressure control should be evaluated and understood.^5^

In recent years, newer classes of GLP1Ra, such as dual and triple agonists, have been launched. They have offered additional advantages by targeting multiple pathways involved in satiety and metabolism. Recent advancements in GLP1Ra therapy include the development of dual and triple agonists, such as tirzepatide (GLP-1/GIP receptor agonist) and retatrutide (GLP-1/GIP/glucagon receptor agonist), which offer additional metabolic benefits and superior weight loss. ^10–12^ Yet, few studies have evaluated the effects of different classes of GLP1 agonists on blood pressure.^13^ Given the increasing use of GLP1Ra in clinical practice, and the introduction of newer agents, we undertook a systematic review and network meta-analysis of RCTs to provide critical insights into the blood pressure-lowering effects of all GLP1Ra classes, including the newer dual and triple agonists.

## Research Design and Methods

This study was reported under the Preferred Reporting Items for a Review and Meta-Analysis (PRISMA), and the checklist was followed. This study was registered in the International Prospective Register of Systematic Reviews (PROSPERO). PubMed/MEDLINE, Web of Science and Ovid/Embase were searched from their inception until July 2024. The search was restricted to studies evaluating GLP-1 RAs, regardless of Food and Drug Administration (FDA) approval.

The search terms included ‘glucagon-like peptide-1 receptor agonists’, ‘GLP-1 agonist’, ‘GLP-1RA’, ‘semaglutide’, ‘dulaglutide’, ‘exenatide’, ‘liraglutide’, ‘tirzepatide’, ‘blood pressure’, ‘systolic blood pressure’, ‘diastolic blood pressure’ and synonyms. The citations of selected articles and any relevant studies that evaluated GLP-1RA and BP reduction were reviewed. After removing duplicates, records were reviewed at the title and abstract level, followed by the screening of full text based on our study criteria. We included randomized-controlled trials from peer-reviewed articles and excluded conference abstracts and non-English literature. The references to the articles and relevant systematic reviews included have also been screened for potentially relevant studies.

### Search Strategy and Eligibility Criteria

RCTs that reported on the use of GLP-1 agonists in adult patients and evaluated blood pressure and weight change were included compared to either placebo or other drug groups. The studies were excluded if 1) they were case reports, editorials, opinion pieces, and review articles, 2) using duplicated populations, such as a post hoc analysis of an included trial, and 3) they were not available as full texts.

### Data Extraction and Quality Assessment

Key participant and intervention characteristics and reported data on efficacy outcomes were extracted independently by two investigators using standard data extraction templates. Any disagreements were resolved by discussion or, if required, by a third author. Data on the following variables were extracted: first author’s name, year of publication, interventional and control treatments, number of randomised patients, and demographic and clinical data (e.g. age, sex).

### Data Synthesis and Analysis

We pooled all estimates using a random-effects model based on the restricted maximum likelihood model using approaches described in the Cochrane Handbook. Effect sizes were expressed using mean differences with 95% confidence intervals (CIs). In studies that did not report SD, CIs were converted to SDs. A funnel plot and the Egger test were used to estimate publication bias. Both Cochran’s Q and Higgins and Thompson’s I2 statistics were generated to describe the heterogeneities among the studies. Forest plots were used to plot the effect size, either for each study or overall. Publication bias was evaluated by graphical inspection of the funnel plot; the estimation of publication bias was quantified by means of the Egger linear regression test.

### Outcome measures

The primary endpoint of this network meta-analysis and meta-regression was the change in SBP and DBP compared to either another drug group or to placebo. The secondary endpoints of this study included changes in weight and HbA1c. Meta-regression analysis was performed to investigate the relationship between the blood pressure-lowering effects and the potential effect modifiers, including duration of type 2 diabetes, weight loss and HbA1c changes. The subgroup analysis was performed based on the 1) dose and 2) single, double or triple agonist status.

### Quality evaluation and sensitivity analysis

The funnel plot was generated, and the Egger’s test was conducted to estimate the potential publication bias. The Trimfill approach was used to estimate where the publication biases didn’t exist. Sensitivity analysis was conducted by including all trials comparing GLP1Ra against other classes of glucose-lowering drugs and also removing high heterogeneity studies.

## Results

### Literature selection and study characteristics

A literature search through July 2024 yielded 981 outputs from PubMed, 998 results from Web of Science Core Collection, and 830 results from Ovid/Embase. After the records were removed for duplication, 849 studies were screened and ultimately, potentially relevant references on GLP1Ra were selected **(Supplementary Figure 1)**.

Finally, 119 articles were included in the meta-analysis, including 115,730 participants. The duration of the intervention varied between 3 weeks and 260 weeks. The mean age was 55.7 years (standard deviation [SD] of 4.2), 54. 6% were male and the mean duration of diabetes was 7.9 years (SD: 4.6). Based on our retrieved publications, 14 GLP1Ra agents were identified. The study characteristics are shown in **Supplementary Table 2**. Most of the studies included were multinational RCTs.

### Meta-analysis: Blood pressure control

For SBP, the network meta-analysis continued of 53 trials involving 38,558 subjects. The results demonstrated that most GLP1Ra drugs did not significantly alter SBP compared to placebo. However, the newer weight loss drugs were associated with significant reductions in SBP. Retatrutide had the largest effect (mean difference [MD]: 7.0 mmHg; 95% Confidence Interval [CI]: 3.4-10.5), followed by tirzepatide (MD: 5.2 mmHg; 95% CI: 3.5-6.9), semaglutide (MD: 3.4 mmHg; 95% CI: 2.1-4.7), liraglutide (MD: 2.7 mmHg; 95% CI: 1.5-3.8), orforglipron (MD: 2.5 mmHg; 95% CI: 0.5-4.5) and dulaglutide (MD: 1.7 mmHg; 95% CI: 0.1-3.4) **(Figure 1–2)**.. Retatrutide resulted in a more significant SBP drop compared to the other drugs as illustrated in **Figure 3**.

**Figure 1.**
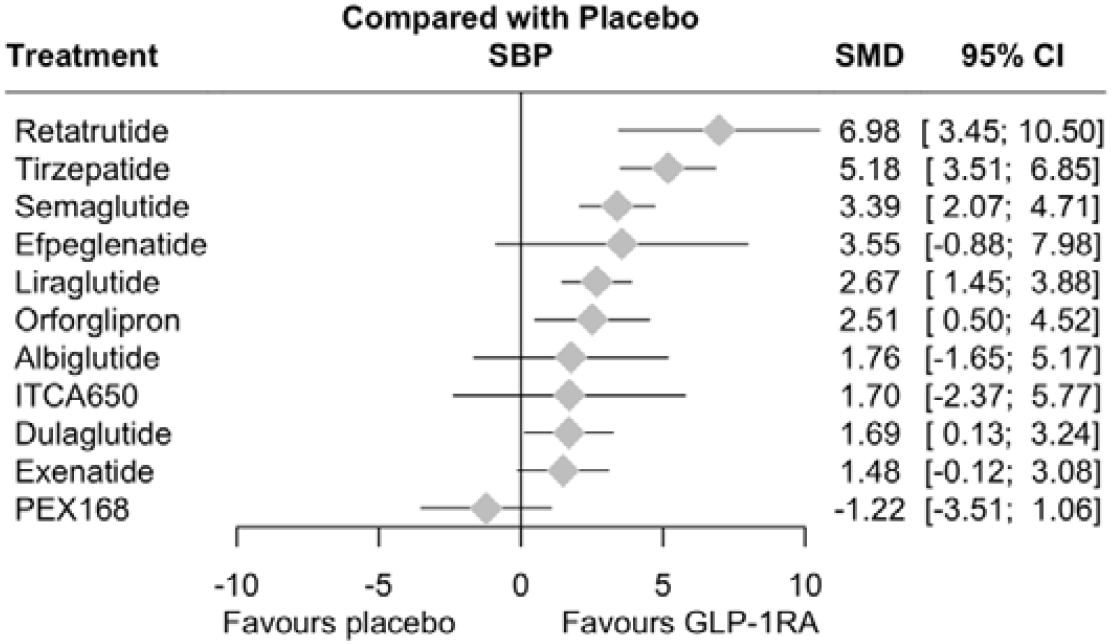
Forest plot of the network analysis of the effects of GLP1Ra on systolic blood pressure stratified by drug types. Unit: mmHg

**Figure 2.**
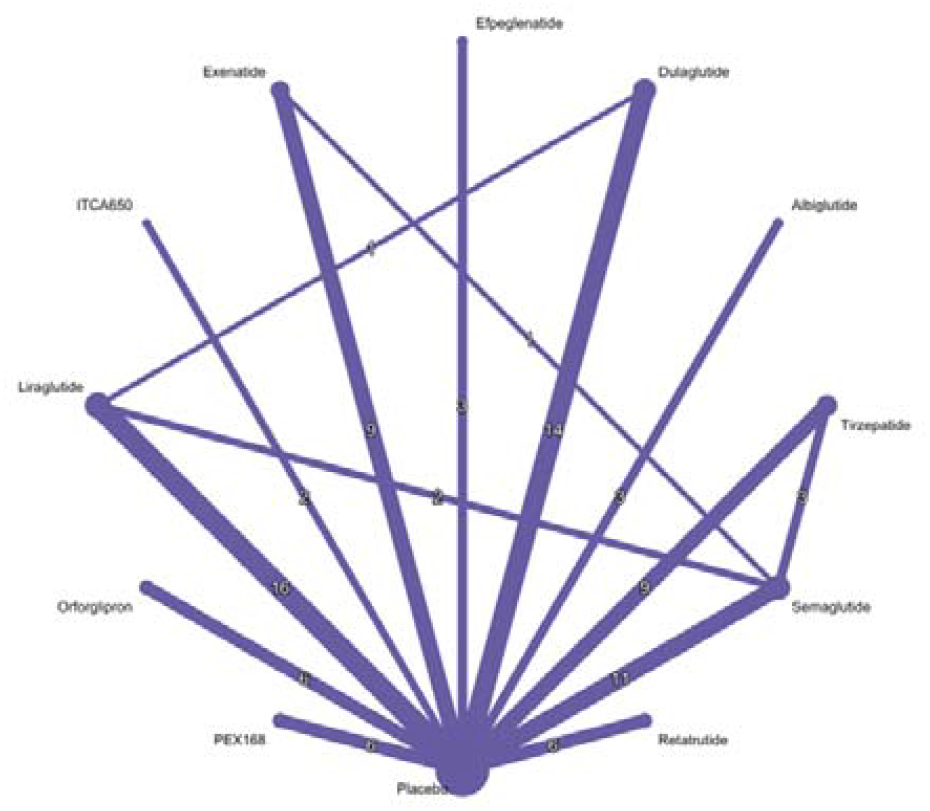
Netgraph of the network analysis of the effects of GLP1Ra on systolic blood pressure stratified by drug types

**Figure 3.**
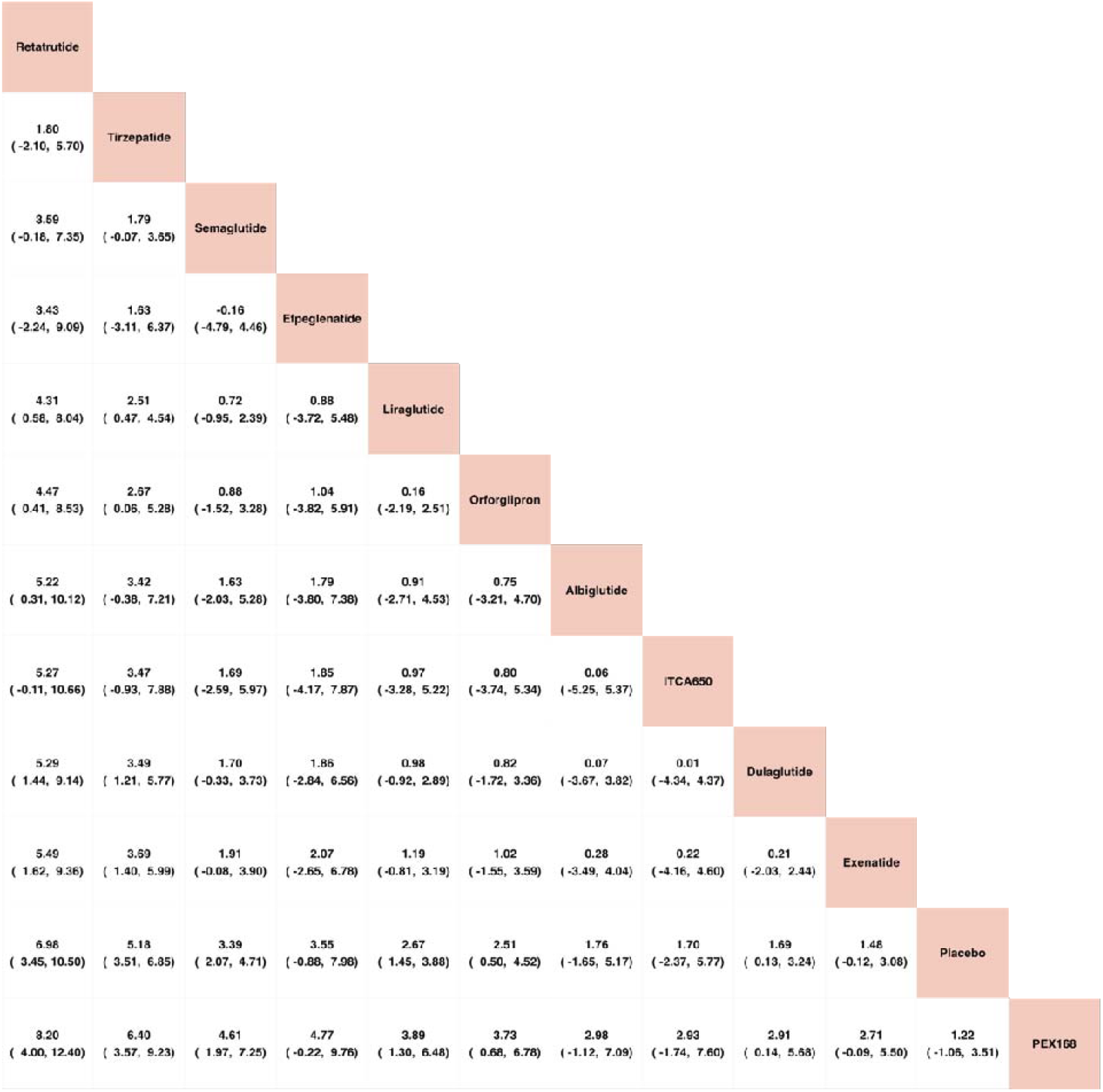
League table of the network analysis of effects of GLP1Ra on systolic blood pressure stratified by drug types. Unit: mmHg

The network meta-analysis included 48 trials involving 35,848 subjects, which reported DBP. Again, most of the GLP1Ra drugs did not significantly alter DBP compared to placebo. Tirzepatide (MD: 1.7 mmHg; 95% CI: 0.8-2.6) was the GLP1Ra that resulted in the most significant reduction in DBP. This was followed by semaglutide (MD: 0.8 mmHg; 95% CI: 0.2-1.4) **(Figure 4–5)**. The head-to-head comparisons of different GLP1Ra drugs on DBP is demonstrated in **Figure 6**.

**Figure 4.**
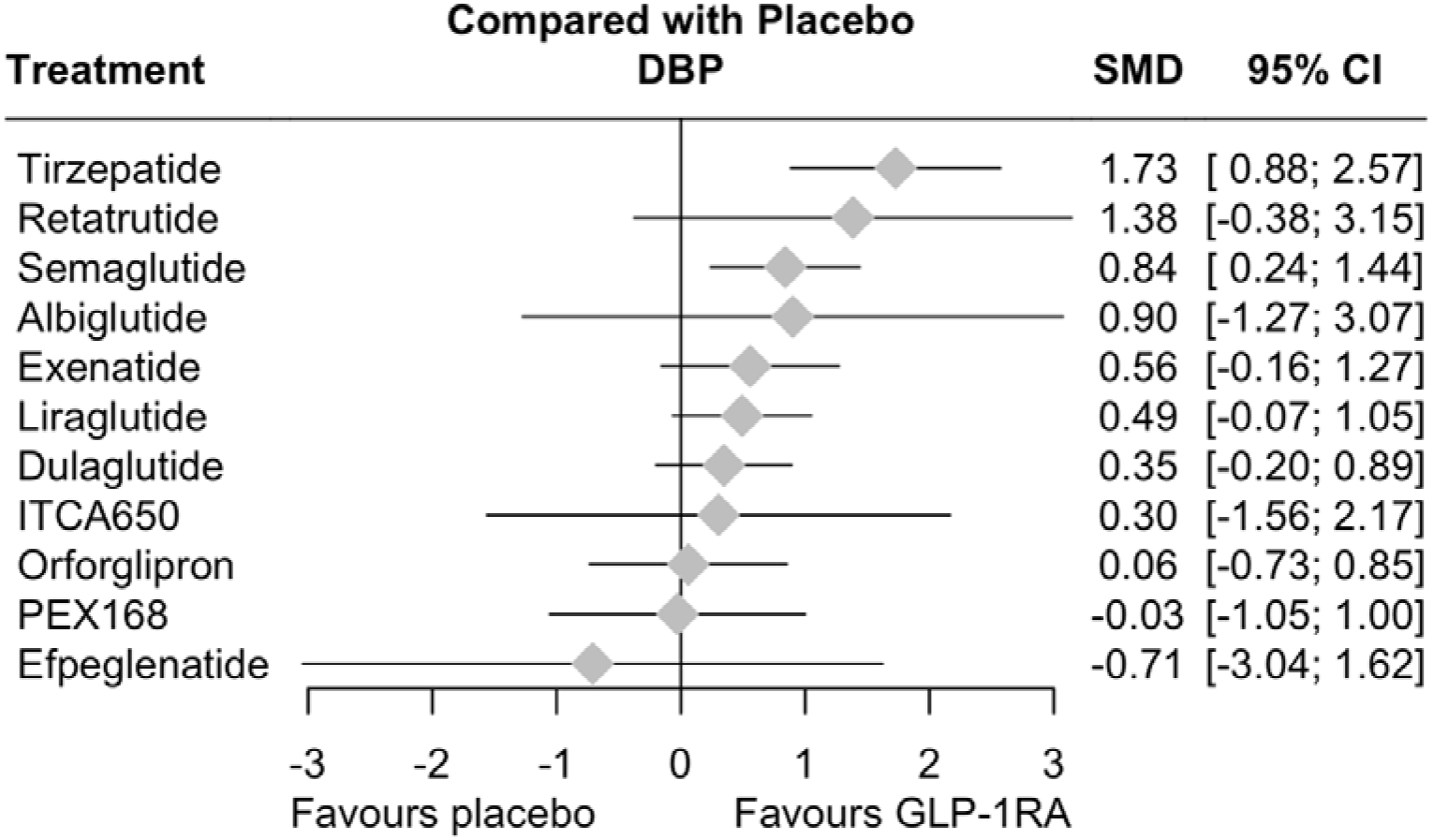
Forest plot of the network analysis of the effects of GLP1Ra on diastolic blood pressure stratified by drug types. Unit: mmHg

**Figure 5.**
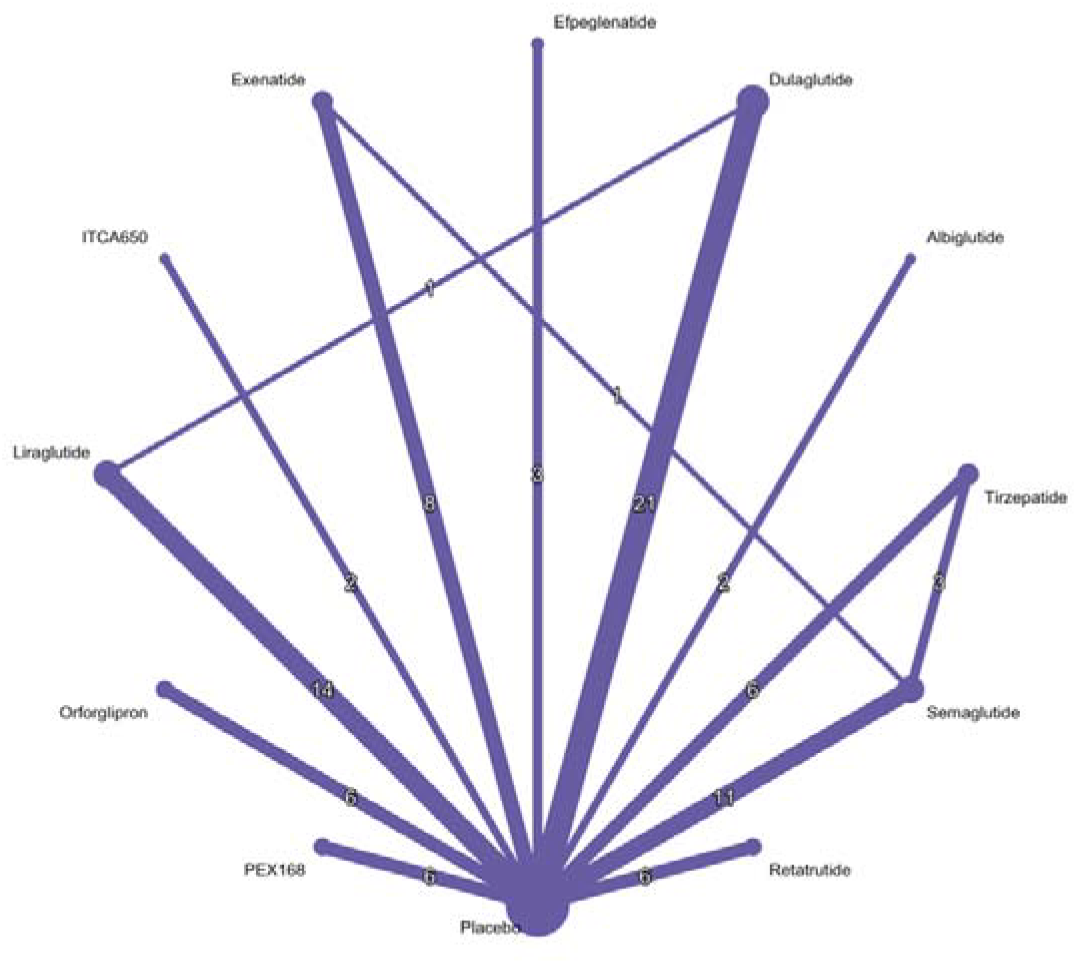
Forest plot of the network analysis of the effects of GLP1Ra on diastolic blood pressure stratified by drug types.

**Figure 6.**
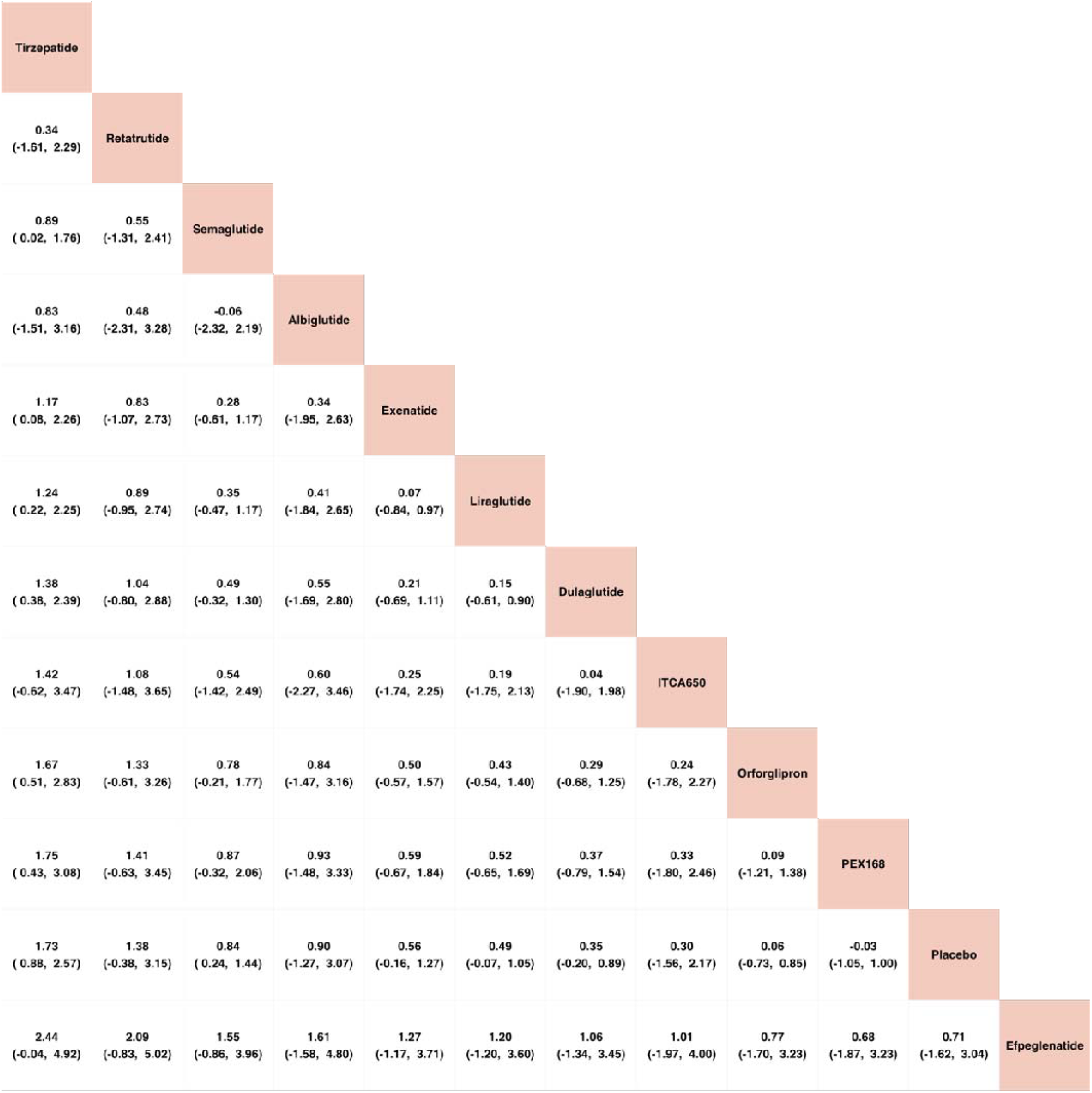
League table of the network analysis of effects of GLP1Ra on diastolic blood pressure stratified by drug types. Unit: mmHg

### Meta-analysis: Body weight change and glycaemic control

The effects of the GLP1Ra on body weight were evaluated by the MD of body weight loss in 67 trials with 41037 participants. The greatest weight loss was observed with tirzepatide (MD: 8.1 kg; 95% CI: 6.5-9.6), followed by orforglipron, retatrutide, semaglutide, then liraglutide. However, GLP1Ra such as mazdutide, ITCA 650, exenatide, efpeglenatide, lixisenatide, PB119, dulaglutide, PEX168 did not significantly lower body weight **(Supplementary Figure 2-3)**. The head-to-head comparisons of different GLP1Ra drugs on weight loss are shown in **Supplementary Figure 4**.

Regarding glycaemic control, 68 trials with 42,648 participants reported HbA1c lowering effects. The greatest HbA1c changes from GLP1Ra rested in tirzepatide (MD: 1.6%; 95% CI: 1.3-1.8), followed by orforglipron, retatrutide, semaglutide, albiglutide, PB119, mazdutide, PEX168, liraglutide, efpeglenatide, exenatide, and lixisenatide and dulaglutide **(Supplementary Figure 5-6)**. The head-to-head comparisons of different GLP1Ra drugs on HbA1c lowering are shown in **Supplementary Figure 7**. In summary, most of the GLP1Ra showed significant efficacy in reducing HbA1c levels.

### Sensitivity analysis and quality assessment

The effects of the GLP1Ra on SBP were hampered by the publication biases such that a significant intercept existed (z = 2.7228, p = 0.0065) **(Supplementary Figure 8)**. Meanwhile, the publication biases against diastolic blood pressure were not statistically significant (z = 0.3674, p = 0.7133) **(Supplementary Figure 9)**. Meanwhile, sensitivity analysis was conducted by including all trials comparing GLP1Ra against other classes of glucose-lowering drugs and by removing high heterogeneity studies **(Supplementary Result 4)**. After including the extra trials, the result remained consistent with the main findings.

### Meta-regressions and Mediation analysis

The impact of baseline effect modifiers on the primary outcomes was assessed using meta-regression analyses. The effects of diabetes duration on the effects of GLP1Ra on SBP were investigated **(Supplementary Figure 12)**. Diabetes duration was positively correlated with SBP lowering effects in semaglutide, dulaglutide, tirzepatide, retatrutide and orforglipron. Meanwhile, diabetes duration was negatively correlated with SBP lowering effects of exenatide, liraglutide, PEX168 and albuglutide. The effects of SBP lowering were positively correlated with age amongst PEX168, orforglipron, retratrutide and tirzepatide but not with other groups **(Supplementary Figure 13)**.

The effects of BP reduction could be secondary to weight changes. Meta-regression was thus conducted to investigate whether weight changes may contribute to BP lowering. The result demonstrated that weight reduction was positively correlated with SBP lowering effects with semaglutide and exenatide, but not on liraglutide or dulaglutide **(Supplementary Figure 14)**. Meanwhile, weight changes were positively correlated with diastolic blood pressure lowering effects amongst semaglutide and exenatide **(Supplementary Figure 15)**.

Mediation analysis was also conducted to attest to how much the BP-lowering effects were mediated by metabolic syndrome **(Supplementary Figure 16-17)**. In the causal mediation analysis regarding the impact of GLP1Ra on BP, the ACME for both weight change and HbA1c change was small and non-significant, indicating a minimal mediation effect **(Supplementary Figure 18)**. Indeed, weight change has a partial mediation effect on SBP reduction, but change in HbA1c did not. Meanwhile, weight change has a partial mediation effect on DBP reduction, but the effect is statistically insignificant **(Supplementary Figure 19-21)**.

### Subgroup analyses

Subgroup analyses of single versus dual or triple agonists status of GLP1Ra were conducted **(Supplementary Figure 22-24)**. The result demonstrated that triple GLP1Ra (MD: 7.0 mmHg; 95% CI: 3.2-10.9) were associated with the greatest SBP lowering effects compared to dual (MD: 5.1 mmHg; 95% CI: 2.8-7.4) and single agonist (MD: 2.2 mmHg; 95% CI: 1.30-3.1). Meanwhile, dual GLP1Ra agonist (MD: 1.2 mmHg; 95% CI: 0.1-2.4) and single agonist (MD: 0. 5 mmHg; 95% CI: 0.1-0.8) but not triple agonist (MD: 1.4 mmHg; 95% CI: −0.4-3.2) was associated with significant DBP lowering effects **(Supplementary Figure 25-27)**. The subgroup analysis of SBP and DBP lowering effects stratified by each drug dosage was also conducted **(Supplementary Figure 34-37)**. Given that the effects of tirzepatide on blood pressure were significant, a subgroup analysis was conducted to investigate the effects of tirzepatide dosage on the BP changes (**Supplementary Figure 42-43)**. The result suggested a direct dose-response between tirzepatide and SBP reduction. However, there was no obvious dose-responsive relationship between tirzepatide and DBP (**Supplementary Figure 44-45)**. Lastly, the effects of the mean age of the study on the BP changes are shown in **Supplementary Figure 46**.

## Discussion

### Main findings

In this network meta-analysis and meta-regression of 11 RCTs, the efficacy of the GLP1Ra on the reduction of blood pressure on adult patients was evaluated. The findings showed that i) retatrutide, tirzepatide, semaglutide, liraglutide, orforglipron and dulaglutide reduced SBP, ii) tirzepatide and semaglutide alone reduced DBP. Notably, retatrutide, tirzepatide, and semaglutide led to the most significant reductions in SBP, while tirzepatide and semaglutide showed the greatest reductions in DBP. The triple agonist retatrutide had the greatest systolic blood pressure lowering effects compared to dual and single agonists. To the best of our knowledge, this is the first network meta-analysis investigating the evidence for SBP and DBP lowering amongst GLP1Ra users.

### Comparisons with previous studies

In a meta-analysis focusing on obese or overweight patients, GLP1Ra reduce SBP by 3.4 and diastolic blood pressure by 1.1 mmHg.^14^ Our results are consistent with a recent meta-analysis which showed that semaglutide decreased SBP by 4.8□mmHg (95% CI −5.65 to −4.02).^15,16^ In the STEP 1 trial, semaglutide 2.4 mg significantly reduced SBP. The SUSTAIN-6 trial (Evaluate Cardiovascular and Other Long-term Outcomes with semaglutide in Subjects with Type 2 Diabetes) showed a dose-dependent SBP decrease.^15^ Liraglutide also exhibited a blood pressure-lowering effect, as evidenced by the SCALE Diabetes trial, which showed that liraglutide 3 mg was more effective in lowering SBP than liraglutide 1.8 mg and placebo.^17^

In the previous meta-analysis, only exenatide reduced DBP.^18^ In our meta-analysis, 3 agents of GLP1Ra, tirzepatide, semaglutide and exenatide, were associated with significant DBP reduction. In a meta-analysis investigating GLP1Ra amongst overweight or obese patients, semaglutide resulted in a reduction of DBP by 1.5□mmHg, while liraglutide resulted in a significant reduction of 0.7 mmHg □.^14^ The effects of GLP1Ra on SBP and DBP were not necessarily in the same direction. In the LEADER study (the Liraglutide Effect and Action in Diabetes: Evaluation of Cardiovascular Outcome), it was suggested that liraglutide resulted in SBP reduction by 1.2 mmHg but DBP elevation by 0.6 mmHg.^19^

Newer GLP1Ra that modulate the incretin pathways are currently entering the treatment paradigm for diabetes and obesity. In our study, triple GLP1Ra had the greatest SBP lowering effects compared to dual or single agonists. Tirzepatide is a dual agonist that acts on both GLP-1 and the GIP receptors. In the SURPASS-1 trial, SBP dropped by 4.7 to 5.2 mmHg compared to 2 mmHg for the placebo.^20^ In the SURMOUNT-1 trial, the reduction in SBP was 6.2 mmHg for tirzepeptide compared to placebo.^11^ In a meta-analysis, tirzepatide, significantly reduced SBP, with a median effect of 4.2 for 5mg, 5.3 for 10 mg and 5.7 for 15 mg.^21^ The greater effect of tirzepatide may be due to the GIP moiety which can stimulate the release of nitric oxide from the endothelial cells and resulting in vasodilatation.^22,23^

Kennedy *et al*., suggested that the reduction in SBP by semaglutide was mediated mainly by weight loss. However, this study showed that this phenomenon was only observed with semaglutide and not in other GLP1Ra, such as liraglutide or tirzepatide. Wong *et al*., GLP1Ra produced significant BP reduction regardless of mean weight loss.^14^ Indeed, some studies suggested that the BP-lowering effects occurred prior to weight reduction^24,25^ GLP1Ra might directly act on endothelial cells and vascular smooth muscle cells where GLP1 receptors are expressed.^26^ In animal studies, GLP1Ra use led to higher expression of endothelial nitric oxide synthase, decreased vascular remodelling, and reduced vascular inflammation.^27,28^

### Clinical implications

In the American Diabetes Association (ADA) and the European Association for the Study of Diabetes (EASD) guidelines, it was highlighted that GLP1Ra should be considered amongst T2DM patients with established or increased cardiovascular risks.^29^ BP reduction of 5 □mmHg lowers the risk of major adverse cardiovascular events by at least 10%. ^30^ The BP lowering effects of some classes of GLP1Ra may be an additional benefit on top of its known cardioprotective effects and weight loss effects. Thus, weight loss with GLP1Ra medications may be a treatment option for hypertension and randomised controlled trials will be helpful for evaluating

We have included the latest available GLP1Ra and provided updated available information regarding GLP1Ra’s effect on BP reduction.^5^ This might aid policy formulation and drug selection for clinical practices. According to the subgroup analysis results, using triple agonists might have greater BP-controlling effects compared to double or single agonists.

The fact that for some classes of drugs, the blood pressure-lowering effects were independent of its weight-reducing effects demonstrated its applicability even amongst populations that were not obese.

Recently, a review conducted by the Korean Society of Hypertension recommended forward-looking policies for GLP1Ra treatment in managing metabolic syndrome.^31^ However, the Korean Society of Hypertension recommendation that the high costs associated with GLP1Ra and its gastrointestinal side effects may result in limited usage of GLP1Ra in controlling BP. For instance, given that the monthly medication costs of using GLP1Ra were over 700 USD, the price of tirzepatide have to be reduced by at least 38% to become cost-effective in terms of weight reduction.^32^

### Limitations

There are several limitations to be acknowledged. Firstly, the exact BP and the lipid profile of each study were not thoroughly provided, such that a meta-regression based on the baseline blood pressure was not conducted. Secondly, most of the studies did not provide the temporal relationship between weight loss and BP reduction, such that its relationship with weight loss maintenance could not be determined. Thirdly, the use and adherence of concomitant BP and lipid-lowering medications were not available. Thus, whether the blood pressure-lowering effects from GLP1Ra were a result of the different drug profiles remained unknown. However, given that the baseline comorbidities between the medication and the placebo group should be randomised, it is believable that the blood pressure-lowering effects of the covariates should be balanced out. Last but not least, some drugs involved few articles, and results must be interpreted with caution when compared with other drugs and in subgroup analyses.

## Conclusions

This network meta-analysis found that GLP1Ra significantly lowered SBP and DBP. Retatrutide, tirzepatide and semaglutide produced in the most marked SBP reduction, whilst tirzepatide and semaglutide resulted in the most marked reductions in DBP. Furthermore, triple GLP1Ra were associated with the greatest SBP lowering effects compared to dual and single agonists.

## Data Availability

All data produced in the present work are contained in the manuscript

## Authors contribution

OHIC: conceptualization, data curation, methodology, validation, writing of original draft, writing of review and editing; HZ: data curation, methodology, validation, writing of original draft; HW: data curation, methodology, validation, writing of review and editing; KR: data curation, methodology, validation, writing of review and editing; DW: data curation, methodology, validation, writing of review and editing; QL: writing of review and editing; JSKC: data interpretation, writing of review and editing; TL: data interpretation, writing of review and editing; GT: data interpretation, writing of review and editing, supervision; CM: data interpretation, writing of review and editing, supervision; IBW: data interpretation, writing of review and editing, supervision

