## Supplementary Appendix for "Differential effects of glucagon-like peptide-1 receptor agonist classes on blood pressure: a systematic review and network meta-analysis of randomised controlled trials with meta-regression"

#### Supplementary Method 1. Search strategy

(glucagon-like peptide-1 receptor agonists OR GLP-1 agonist OR GLP-1RA) AND (blood pressure OR systolic blood pressure OR diastolic blood pressure)

*PubMed search term: 981 output*

*(”glucagon like peptide 1 receptor agonists”[Pharmacological Action] OR ”glucagon like peptide 1 receptor agonists”[MeSH Terms] OR (”glucagon like”[All Fields] AND ”peptide 1”[All Fields] AND ”receptor”[All Fields] AND ”agonists”[All Fields]) OR ”glucagon like peptide 1 receptor agonists”[All Fields] OR ”glucagon like peptide 1 receptor agonists”[All Fields] OR ((”glucagon like peptide 1”[MeSH Terms] OR ”glucagon like peptide 1”[All Fields] OR ”glp 1”[All Fields]) AND (”agonist”[All Fields] OR ”agonist s”[All Fields] OR ”agonistic”[All Fields] OR ”agonistically”[All Fields] OR ”agonistics”[All Fields] OR ”agonists”[MeSH Subheading] OR ”agonists”[All Fields])) OR ”GLP-1RA”[All Fields]) AND (”blood pressure”[MeSH Terms] OR (”blood”[All Fields] AND ”pressure”[All Fields]) OR ”blood pressure”[All Fields] OR ”blood pressure determination”[MeSH Terms] OR (”blood”[All Fields] AND ”pressure”[All Fields] AND ”determination”[All Fields]) OR ”blood pressure determination”[All Fields] OR ”arterial pressure”[MeSH Terms] OR (”arterial”[All Fields] AND ”pressure”[All Fields]) OR ”arterial pressure”[All Fields] OR (”blood pressure”[MeSH Terms] OR (”blood”[All Fields] AND ”pressure”[All Fields]) OR ”blood pressure”[All Fields] OR (”systolic”[All Fields] AND ”blood”[All Fields] AND ”pressure”[All Fields]) OR ”systolic blood pressure”[All Fields]) OR (”blood pressure”[MeSH Terms] OR (”blood”[All Fields] AND ”pressure”[All Fields]) OR ”blood pressure”[All Fields] OR (”diastolic”[All Fields] AND ”blood”[All Fields] AND ”pressure”[All Fields]) OR ”diastolic blood pressure”[All Fields]))*

*998 results from Web of Science Core Collection*

*ALL((glucagon-like peptide-1 receptor agonists OR GLP-1 agonist OR GLP-1RA) AND (blood pressure OR systolic blood pressure OR diastolic blood pressure))*

*830 results from Ovid/Embase*

*((glucagon-like peptide-1 receptor agonists or GLP-1 agonist or GLP-1RA) and (blood pressure or systolic blood pressure or diastolic blood pressure)).mp. [mp=title, abstract, heading word, drug trade name, original title, device manufacturer, drug manufacturer, device trade name, keyword heading word, floating subheading word, candidate term word]*

### Supplementary Table 1. Characteristics of included study

| Study | Country and region | Design | Follow-up duration (weeks) | Number of participants | Randomised treatment | Dose and frequency | Diabetes duration (years) | Age | Male (%) |
|---|---|---|---|---|---|---|---|---|---|
| Ahmadi 2019[1] | Sweden | RCT | 24.0 | 122 | Liraglutide<br>Placebo | 1.80<br>- | 17.3<br>- | 63.8<br>- | 63.5<br>- |
| Ahmann 2015[2] | Multinational | RCT | 26.0 | 451 | Liraglutide<br>Placebo | 1.80<br>- | 12.1<br>- | 59.3<br>- | 52.0<br>- |
| Ahmann 2018[3] | Multinational | RCT | 56.0 | 809 | Semaglutide<br>Exenatide | 1.00<br>- | 9.0<br>- | 56.4<br>- | 54.2<br>- |
| Ahren 2014[4] | Sweden | RCT | 104.0 | 1049 | Albiglutide<br>Sitagliptin<br>Glimepiride | 30Qwsc<br>100.00<br>2.00 | 6.0<br>5.8<br>6.0 | 54.3<br>54.3<br>54.4 | 44.7<br>46.0<br>51.5 |
| Ahren 2017[5] | Multinational | RCT | 56.0 | 1231 | Semaglutide<br>Semaglutide<br>Sitagliptin | 0.50<br>1.00<br>- | 6.4<br>6.7<br>- | 54.8<br>56.0<br>- | 51.0<br>50.0<br>- |
| Aroda 2023[6] | Multinational | RCT | 52.0 | 2294 | Semaglutide<br>Semaglutide<br>Semaglutide | 25.00<br>50.00<br>- | 9.7<br>8.9<br>- | 65.0<br>-<br>- | 57.0<br>-<br>- |
| Aroda 2019[7] | Multinational | RCT | 26.0 | 703 | Semaglutide<br>Semaglutide<br>Semaglutide<br>Placebo | 3.00<br>7.00<br>14.00<br>- | -<br>-<br>-<br>- | 55.0<br>55.0<br>54.0<br>- | 50.0<br>50.0<br>49.0<br>- |
| Astrup 2009[8] | Denmark | RCT | 20.0 | 564 | Liraglutide<br>Liraglutide<br>Liraglutide<br>Liraglutide<br>Orlistat | 3.00<br>2.40<br>1.80<br>1.20<br>- | -<br>-<br>-<br>-<br>- | 49.5<br>45.0<br>45.5<br>47.2<br>- | 25.0<br>24.0<br>24.0<br>24.0<br>23.0 |
| Barnett 2007 [9] | UnitedKingdom | RCT | 0.00 | 138 | Exenatide<br>InsulinGlargine | 0.00<br>- | 7.4<br>- | -<br>- | 47.1<br>- |
| Blackman 2016[10] | Multinational | RCT | 32.0 | 359 | Liraglutide<br>Placebo | 3.00<br>- | -<br>- | 48.5<br>- | 71.9<br>- |
| Diamant 2010[11] | Netherland | RCT | 26.0 | 29 | Exenatide<br>InsulinGlargine | 2.00<br>- | 8<br>- | 58.0<br>- | 42.0<br>- |

| Study | Country and region | Design | Follow-up duration (weeks) | Number of participants | Randomised treatment | Dose and frequency | Diabetes duration (years) | Age | Male (%) |
|---|---|---|---|---|---|---|---|---|---|
| Blevins 2011[12] | USA | RCT | 24.0 | 252 | Exenatide<br>Exenatide | 2.00<br>- | 7<br>- | -<br>- | 60.0<br>- |
| Blonde 2015[13] | Multinational | RCT | 52.0 | 884 | Dulaglutide<br>Dulaglutide | 1.50<br>0.75 | 12.8<br>12.4 | 58.9<br>59.3 | 54.0<br>50.0 |
| Blonde 2020[14] | Multinational | RCT | 26.0 | 302 | Liraglutide<br>Placebo | 1.80<br>- | 10.1<br>- | 54.7<br>- | 62.0<br>- |
| Bunck 2009[15] | Multinational | RCT | 52.0 | 69 | Exenatide<br>InsulinGlargine | 1.20<br>- | 5.7<br>- | 58.4<br>- | 64.0<br>- |
| Buse 2004[16] | USA | RCT | 52.0 | 377 | Exenatide<br>Placebo | 0.01<br>- | 9.3<br>- | 55.0<br>- | 61.0<br>- |
| Buse 2011[17] | Multinational | RCT | 30.0 | 261 | Exenatide<br>Placebo | 12.0<br>- | 5.9<br>- | 59.0<br>- | 51.0<br>- |
| Capehorn 2020[18] | Multinational | RCT | 30.0 | 577 | Semaglutide<br>Liraglutide | 1.00<br>1.20 | 9.6<br>8.9 | 60.1<br>58.9 | 58.2<br>- |
| Chen 2016[19] | China | RCT | 12 | 120 | PEX168<br>PEX168 | Placebo<br>- | 0.10<br>0.20 | 4.3<br>4.0 | 53□7<br>56.4 |
| Dahl 2022[20] | Multinational | RCT | 40 | 475 | Tirzepatide<br>Tirzepatide<br>Tirzepatide<br>Placebo | 5.00<br>10.00<br>15.00<br>- | 14.1<br>12.6<br>13.7<br>- | 62<br>61<br>61<br>- | 53.0<br>61.0<br>54.0<br>- |
| Davies 2009[21] | UnitedKingdom | RCT | 9.0 | 235 | Exenatide<br>InsulinGlargine | 0.01<br>- | 7.5<br>- | 55.0<br>- | 68.0<br>- |
| Davies 2015[22] | Multinational | RCT | 56.0 | 846 | Liraglutide<br>Liraglutide<br>Placebo | 1.80<br>- | 7.4<br>7.5 | 54.9<br>- | 51.2<br>- |
| Defronzo 2005[23] | USA | RCT | 30.0 | 336 | Exenatide<br>Exenatide<br>Placebo | 0.01<br>- | 6.2<br>4.9 | 53.0<br>52.0 | 51.8<br>60.2 |
| DelPrato 2020[24] | USA | RCT | 17 | 207 | Efpeglenatide<br>Efpeglenatide<br>Efpeglenatide<br>Placebo | 8.00<br>12.00<br>16.00 | 9.1<br>7.4<br>7.2 | 56.7<br>56.0<br>56.4 | 36.5<br>53.8<br>47.2 |
| Drucker 2008[25] | USA | RCT | 30.0 | 295 | Exenatide<br>Exenatide | 2.00<br>- | 7<br>- | 55<br>- | 55.0<br>- |

| Study | Country and region | Design | Follow-up duration (weeks) | Number of participants | Randomised treatment | Dose and frequency | Diabetes duration (years) | Age | Male (%) |
|---|---|---|---|---|---|---|---|---|---|
| Dungan 2014[26] | Multinational | RCT | 26 | 599 | Dulaglutide | 1.50 | 7.1 | 56.5 | 46.0 |
| | | | | | Liraglutide | - | - | - | - |
| Frias 2018[27] | Multinational | RCT | 26.0 | 316 | LY3298176 | 1.00 | 7.8 | 57.4 | 56.0 |
| | | | | | LY3298176 | 5.00 | 8.9 | 57.9 | 62.0 |
| | | | | | LY3298176 | 10.00 | 7.9 | 56.5 | 59.0 |
| | | | | | LY3298176 | 15.00 | 8.5 | 56 | 42.0 |
| | | | | | Dulaglutide | 1.50 | 9.3 | 58.7 | 44.0 |
| | | | | | Placebo | | | | |
| Frias 2023[28] | USA | RCT | 32 | 92 | CagriSema | 2.40 | 6.4 | 56 | 58.0 |
| | | | | | Semaglutide | 2.40 | 9.2 | 57 | 58.0 |
| | | | | | Cagrilintide | 2.40 | 10.7 | 62 | 77.0 |
| Frias 2021[29] | Multinational | RCT | 40 | 1842 | Semaglutide | 2.00 | 9.2 | 57.9 | 58.0 |
| | | | | | Semaglutide | - | - | - | - |
| Frias 2021[30] | Multinational | RCT | 40 | 1842 | Dulaglutide | 3.00 | 7.6 | 56.9 | 53.2 |
| | | | | | Dulaglutide | 4.50 | 7.7 | 56.6 | 51.8 |
| Frias 2021[31] | Multinational | RCT | 40 | 1878 | Tirzepatide | 5.00 | 9.1 | 56.3 | 46.3 |
| | | | | | Tirzepatide | 10.00 | 8.4 | 57.2 | 50.7 |
| | | | | | Tirzepatide | 15.00 | 8.7 | 55.9 | 45.5 |
| | | | | | Semaglutide | | | | |
| Frias 2023[32] | Multinational | RCT | 26 | 383 | Orforglipron | 3.00 | 5.0 | 59 | 51.0 |
| | | | | | Orforglipron | 12.00 | 7.1 | 57.4 | 64.0 |
| | | | | | Orforglipron | 24.00 | 5.9 | 60.5 | 64.0 |
| | | | | | Orforglipron | 36.00 | 5.9 | 59.7 | 59.0 |
| | | | | | Orforglipron | 45.00 | 6.8 | 58.5 | 60.0 |
| | | | | | Dulaglutide | 1.50 | 7.9 | 58.8 | 44.0 |
| | | | | | Placebo | | | | |
| Frias 2022[33] | USA | RCT | 56 | 406 | Efpeglenatide | 2.00 | 5.3 | 58.6 | 55.0 |
| | | | | | Efpeglenatide | 4.00 | 4.9 | 56.3 | 51.5 |
| | | | | | Efpeglenatide | 6.00 | 5.2 | 59.6 | 59.2 |
| | | | | | Placebo | | | | |
| Frias 2019[34] | Multinational | RCT | 18 | 318 | Dulaglutide | 1.50 | 7.9 | 57.7 | 48.1 |
| | | | | | Dulaglutide | 3.00 | 7.5 | 55.9 | 44.3 |
| | | | | | Dulaglutide | 4.50 | 9.0 | 52.6 | 47.4 |
| | | | | | Placebo | | | | |
| Gao 2009[35] | China | RCT | 16.0 | 466 | Exenatide | 0.02 | - | 53.6 | 48.0 |
| | | | | | Placebo | | | | |

| Study | Country and region | Design | Follow-up duration (weeks) | Number of participants | Randomised treatment | Dose and frequency | Diabetes duration (years) | Age | Male (%) |
|---|---|---|---|---|---|---|---|---|---|
| Gao 2020[36] | China | RCT | 24.0 | 533 | PEX168<br>PEX168<br>Placebo | 0.10<br>0.20 | 4.3<br>4.8 | 53.6<br>52.8 | 57.0<br>60.6 |
| Garber 2008[37] | USA | RCT | 30.0 | 746 | Liraglutide<br>Liraglutide<br>Glimepiride | 1.80<br>1.20 | -<br>- | -<br>- | 49.0<br>47.0 |
| Garvey 2023[38] | Multinational | RCT | 72 | 938 | Tirzepatide<br>Tirzepatide<br>Placebo | 10.00<br>15.00 | 17.6<br>17.5 | 54.3<br>53.6 | 49.0<br>49.0 |
| Gill 2010[39] | Multinational | RCT | 52.0 | 54 | Exenatide<br>Placebo | 0.01 | 7 | 57 | 68.0 |
| Giorgino 2015[40] | Multinational | RCT | 78.0 | 810 | Dulaglutide<br>Dulaglutide<br>InsulinGlargine | 1.50<br>0.75 | 9<br>9 | 56<br>57 | 53.0<br>50.0 |
| Grunberger 2012[41] | Multinational | RCT | 12.0 | 164 | Dulaglutide<br>Dulaglutide<br>Dulaglutide<br>Dulaglutide<br>Placebo | 0.10<br>0.50<br>1.00<br>1.50 | 3.9<br>3.7<br>3.3<br>4.6 | 56.3<br>56.9<br>57.2<br>57.5 | 31.4<br>47.1<br>47.1<br>44.8 |
| Heise 2022[42] | Germany | RCT | 28.0 | 127 | Tirzepatide<br>Semaglutide<br>Placebo | 15.00<br>1.00 | 10.24<br>12.73 | 61.1<br>63.7 | 69.0<br>77.0 |
| Horber 2020[43] | Switzerland | RCT | 96.0 | 95 | Liraglutide<br>Placebo | 1.80 | - | 54 | 11.6 |
| Inagaki 2022[44] | Japan | RCT | 52.0 | 636 | Tirzepatide<br>Tirzepatide<br>Tirzepatide<br>Dulaglutide | 5.00<br>10.00<br>15.00 | 4.5<br>5.1<br>5.1 | 56.8<br>56.2<br>56.0 | 71.0<br>75.0<br>83.0 |
| Iwamoto 2009[45] | China | RCT | 10.0 | 29 | Exenatide<br>Exenatide<br>Placebo | 0.80<br>2.00 | 6<br>5 | 56<br>58 | 50.0<br>88.9 |
| Ji 2021[24] | China | RCT | 12.0 | 250 | PB119<br>PB119<br>PB119<br>PB119<br>Placebo | 0.08<br>0.15<br>0.20<br>- | 3.20<br>3.28<br>3.48<br>- | 50.8<br>51.4<br>50.5<br>- | 77.4<br>65.1<br>49.2<br>- |

| Study | Country and region | Design | Follow-up duration (weeks) | Number of participants | Randomised treatment | Dose and frequency | Diabetes duration (years) | Age | Male (%) |
|---|---|---|---|---|---|---|---|---|---|
| Jiang 2022[46] | China | RCT | 12.0 | 42 | Dulaglutide<br>Mazdutide<br>Mazdutide<br>Mazdutide<br>Mazdutide<br>Placebo | 1.50<br>3.00<br>4.50<br>6.00<br>- | 3.3<br>2.1<br>4.7<br>6.0<br>- | 50.7<br>58.6<br>47.9<br>54.6<br>- | 1.000<br>0.750<br>0.625<br>0.625<br>- |
| Kadowaki 2022[47] | Multinational | RCT | 68.0 | 28 | Semaglutide<br>Semaglutide<br>Placebo | 1.70<br>2.40 | 7.7<br>7.8 | 52<br>51 | 0.057<br>0.630 |
| Kadowaki 2022[48] | Japan | RCT | 52.0 | 443 | Tirzepatide<br>Tirzepatide<br>Tirzepatide | 5.00<br>10.00<br>15.00 | 8.5<br>9.1<br>8.5 | 57.7<br>56.9<br>56.5 | 0.800<br>0.770<br>0.700 |
| Kadowaki 2011[49] | Japan | RCT | 24.0 | 179 | Exenatide<br>Exenatide<br>Placebo | 0.01<br>0.01 | 12.2<br>11.6 | 58.5<br>59.4 | 0.680<br>0.680 |
| Kendall 2005[50] | USA | RCT | 56.0 | 733 | Exenatide<br>Placebo | 0.01 | - | 45.2 | 0.213 |
| Kim 2007[51] | USA | RCT | 12.0 | 51 | Liraglutide<br>Metformin | 1.80 | - | - | - |
| Kim 2013[52] | USA | RCT | 68.0 | 68 | Liraglutide<br>Placebo | 1.80 | 0.33 | 58 | 27.0 |
| Knop 2023[53] | Multinational | RCT | 68.0 | 709 | Semaglutide<br>Placebo | 50.00 | 35.1 | - | 27.0 |
| Kosiborod 2023[54] | Multinational | RCT | 68.0 | 1961 | Semaglutide<br>Placebo | 2.40 | 46 | - | 21.0 |
| Koska 2021[55] | USA | RCT | 18.0 | 163 | Exenatide<br>Placebo | 2.00 | 6 | 63 | 0.871 |
| Krajnc 2023[56] | Multinational | RCT | 26.0 | 39 | Semaglutide,Liraglutide<br>usualcareweightmanagement | 3.30 | 33.6 | 0.109 | |
| Larsen 2017[57] | Denmark | RCT | 16.0 | 103 | Liraglutide<br>Placebo | 3.00 | 42.5 | 0.583 | |
| Lin 2022[58] | USA | RCT | 12.0 | 50 | Liraglutide+Empagliflozin<br>Liraglutide | 1.20 | 50.7 | 0.540 | |

| Study | Country and region | Design | Follow-up duration (weeks) | Number of participants | Randomised treatment | Dose and frequency | Diabetes duration (years) | Age | Male (%) |
|---|---|---|---|---|---|---|---|---|---|
| Liutkus 2010[59] | Multinational | RCT | 26.0 | 165 | Exenatide<br>Placebo | 0.01 | 6.3 | 55 | 0.600 |
| Lovshin 201[60] | Canada | RCT | 3.0 | 20 | Liraglutide<br>Placebo | 1.80 | 5.8 | 62 | 0.950 |
| Ludvik 2018[61] | Multinational | RCT | 24.0 | 423 | Dulaglutide<br>Dulaglutide<br>Placebo | 1.50<br>0.75 | 9.21<br>10.05 | 56.17<br>58.55 | 0.540<br>0.490 |
| Ludvik 2021[62] | Multinational | RCT | 52.0 | 1437 | Tirzepatide<br>Tirzepatide<br>Tirzepatide<br>Insulin Degludec | 5.00<br>10.00<br>15.00 | 8.5<br>8.4<br>8.5 | 57.2<br>57.4<br>57.5 | 0.560<br>0.540<br>0.540 |
| Masmiquel 2016[63] | Multinational | RCT | 260.0 | 9340 | Liraglutide<br>Placebo | 1.80 | 12.1 | 64.3 | 64.3 |
| Mensberg 2016[64] | Japan | RCT | 16.0 | 36 | Liraglutide<br>Placebo | 0.60 | - | - | 69.0 |
| Miyagawa 2015 [65] | Japan | RCT | 26.0 | 487 | Dulaglutide<br>Liraglutide<br>Placebo | 0.75<br>0.90 | 6.8<br>6.3 | 57.2<br>57.9 | 81.0<br>83.0 |
| Moretto 2008 [66] | Multinational | RCT | 24 | 232 | Exenatide<br>Exenatide<br>Placebo | 0.01<br>0.01 | 2<br>2 | 55<br>56 | 55.0<br>54.0 |
| Nauck 2007[67] | Multinational | RCT | 52.0 | 501 | Exenatide<br>Placebo | 0.01 | 2 | 55 | 0.540 |
| Nauck 2009[68] | Multinational | RCT | 26.0 | 1091 | Liraglutide<br>Liraglutide<br>Liraglutide<br>Glimepiride | 1.80<br>1.80<br>0.60 | 7<br>7<br>8 | 57<br>57<br>56 | 59.0<br>54.0<br>62.0 |
| Nauck 2016[69] | Multinational | RCT | 52.0 | 309 | Albiglutide<br>Albiglutide<br>Placebo | 30.00<br>50.00 | 3.4<br>4.2 | 53.6<br>52 | 57.4<br>50.5 |
| Nauck 2014[70] | Multinational | RCT | 26.0 | 1098 | Dulaglutide<br>Dulaglutide<br>Sitagliptin<br>Placebo | 1.50<br>0.75<br>100.00 | 7<br>7<br>7 | 54<br>54<br>54 | 48.0<br>44.0<br>48.0 |

| Study | Country and region | Design | Follow-up duration (weeks) | Number of participants | Randomised treatment | Dose and frequency | Diabetes duration (years) | Age | Male (%) |
|---|---|---|---|---|---|---|---|---|---|
| Odawara 2016[71] | Japan | RCT | 52.0 | 422 | Dulaglutide<br>Liraglutide | 0.75<br>0.90 | 6.8<br>6.3 | 57.2<br>57.9 | 81.0<br>83.0 |
| Palikhe 2013[72] | India | RCT | 54.0 | 31 | Exenatide<br>Gastrectomy | 0.01 | 8.5 | 49.6 | 26.0 |
| Pan 2014[73] | Multinational | RCT | 24.0 | 391 | Lixisenatide<br>Placebo | 0.02 | 6.5 | 54.5 | 51.5 |
| Pinget 2013[74] | Multinational | RCT | 24.0 | 484 | Lixisenatide<br>Placebo | 0.02 | 8.1 | 56 | 53.0 |
| PiSunyer 2015[75] | Multinational | RCT | 56.0 | 3731 | Liraglutide<br>Placebo | 3.00 | 0.213 | 45.1 | - |
| Pratley 2010[76] | Multinational | RCT | 26 | 665 | Liraglutide<br>Liraglutide<br>Sitagliptin | 1.80<br>1.20 | 6.4<br>6.3 | 55<br>55.9 | 52.0<br>52.0 |
| Pratley 2020[77] | Multinational | RCT | 52.0 | 1199 | Semaglutide<br>Dulaglutide | 5.00 | 10.7 | 60.3 | 52.5 |
| Pratley 2018[78] | Multinational | RCT | 40.0 | 1201 | Semaglutide<br>Dulaglutide | 14.00<br>1.80 | 7.8<br>7.3 | 56<br>56 | 52.0<br>53.0 |
| Pratley 2019[79] | Multinational | RCT | 52 | 711 | Semaglutide<br>Liraglutide<br>Placebo | 14.0<br>1.80 | 7.8<br>7.3 | 56<br>56 | 52.0<br>52.0 |
| Pratt 2023[80] | Multinational | RCT | 12.0 | 51 | Orforglipron<br>Orforglipron<br>Orforglipron<br>Orforglipron<br>Orforglipron<br>Placebo | 9.00<br>15.00<br>21.00<br>27.00<br>45.00<br>- | 13.48<br>15.02<br>9.48<br>7.6<br>10.38<br>- | 57.7<br>59.6<br>55.3<br>58.8<br>62.8<br>- | 70.0<br>71.4<br>77.8<br>44.0<br>62.7<br>- |
| Ratner 2010[81] | USA | RCT | 13.0 | 542 | Lixisenatide<br>Lixisenatide<br>Lixisenatide<br>Lixisenatide<br>Lixisenatide<br>Lixisenatide<br>Lixisenatide<br>Lixisenatide<br>Placebo | 0.005 QD<br>0.01 QD<br>0.02 QD<br>0.03 QD<br>0.005BID<br>0.01 BID<br>0.02 BID<br>0.03 BID | 7.2<br>6.2<br>6.4<br>6.0<br>6.2<br>6.4<br>6.6<br>7.0 | 56.8<br>55.4<br>55.4<br>56.5<br>57.1<br>56.0<br>56.7<br>55.3 | 47.3<br>59.6<br>50.9<br>50.0<br>47.2<br>51.8<br>37.0<br>42.6 |
| Riddle 2013[82] | Multinational | RCT | 24 | 495 | Lixisenatide<br>Placebo | 0.02 | 12.5 | 57 | 45.0 |

| Study | Country and region | Design | Follow-up duration (weeks) | Number of participants | Randomised treatment | Dose and frequency | Diabetes duration (years) | Age | Male (%) |
|---|---|---|---|---|---|---|---|---|---|
| Rodbard 2018[83] | Multinational | RCT | 30.0 | 396 | Semaglutide<br>Semaglutide<br>Placebo | 0.50<br>1.00 | 12.9<br>13.7 | 59.1<br>58.5 | 56.1<br>58.8 |
| Rosenstock 2014[84] | Multinational | RCT | 24.0 | 859 | Lixisenatide<br>Placebo | 0.02 | 9.1 | 59.1 | 0.495 |
| Rosenstock 2023[85] | USA | RCT | 36 | 162 | Retatrutide<br>Retatrutide<br>Dulaglutide<br>Placebo | 0.50<br>4.00<br>1.50 | 8.8<br>10.5<br>7.2 | 57.2<br>57.6<br>54.9 | 51.0<br>50.0<br>28.0 |
| Rosenstock 2018[86] | USA | RCT | 39.0 | 460 | ITCA650<br>ITCA650<br>Placebo | 0.04<br>0.06 | 9.1<br>8.9 | 55.5<br>54.7 | 58.2<br>59.5 |
| Rosenstock 2019[87] | Multinational | RCT | 12.0 | 252 | Efpeglenatide<br>Efpeglenatide<br>Efpeglenatide<br>Efpeglenatide<br>Efpeglenatide<br>Efpeglenatide<br>Liraglutide<br>Placebo | 0.30<br>1.00<br>2.00<br>3.00<br>4.00<br>1.80<br>1.80 | 7.1<br>5.1<br>5.8<br>5.9<br>6.1<br>6.4<br>6.4 | 56<br>55<br>56<br>54<br>56<br>54<br>54 | 0.650<br>0.510<br>0.550<br>0.640<br>0.640<br>0.440<br>0.440 |
| Rosenstock 2021[88] | Multinational | RCT | 40 | 478 | Tirzepatide<br>Tirzepatide<br>Tirzepatide<br>Tirzepatide<br>Placebo | 5.00<br>10.00<br>15.00<br>15.00 | 4.7<br>4.9<br>4.8<br>4.8 | 54.1<br>55.8<br>52.9<br>52.9 | 0.460<br>0.600<br>0.500<br>0.500 |
| Rubino 2021[89] | USA | RCT | 48.0 | 902 | Semaglutide<br>Placebo | 2.40 | 2.0 | 51.7 | 0.360 |
| Ryan 2020 [90] | Multinational | RCT | 68.0 | 17500 | Semaglutide | 2.40 | - | 51.7 | 36.0 |
| Seino 2008[91] | Japan | RCT | 14.0 | 225 | Liraglutide<br>Liraglutide<br>Liraglutide<br>Liraglutide<br>Placebo | 0.30<br>0.60<br>0.90<br>0.90 | 7.15<br>6.78<br>8.87<br>7.62 | 56.5<br>56.8<br>60<br>55.5 | 68.9<br>68.9<br>62.0<br>70.0 |
| Seino 2012 [92] | Multinational | RCT | 24.0 | 311 | Lixisenatide<br>Placebo | 0.02 | 13.7 | 58.7 | 44.8 |

| Study | Country and region | Design | Follow-up duration (weeks) | Number of participants | Randomised treatment | Dose and frequency | Diabetes duration (years) | Age | Male (%) |
|---|---|---|---|---|---|---|---|---|---|
| Seino 2014[93] | Japan | RCT | 16.0 | 212 | Albiglutide | 15.00 | 6.3 | 53.3 | 61.5 |
| | | | | | Albiglutide | 30Qwsc | 7.8 | 58 | 70.4 |
| | | | | | Albiglutide | 30BIWsc | 7.2 | 59.1 | 77.4 |
| | | | | | Placebo | | | | |
| Shuai 2021[94] | China | RCT | 24.0 | 361 | PEX168 | 0.10 | 1.0 | 50 | 67.0 |
| | | | | | PEX168 | 0.20 | 1.5 | 52.4 | 55.0 |
| | | | | | Placebo | | | | |
| Skrivanek 2014[95] | USA | RCT | 26.0 | 230 | Dulaglutide | 0.75 | 7 | - | 48.0 |
| | | | | | Dulaglutide | 1.50 | 9 | - | 40.0 |
| | | | | | Dulaglutide | 1.00 | 7 | - | 30.0 |
| | | | | | Dulaglutide | 2.00 | 7 | - | 27.0 |
| | | | | | Dulaglutide | 3.00 | 7 | - | 33.0 |
| | | | | | Dulaglutide | 0.25 | 52 | - | 28.0 |
| | | | | | Dulaglutide | 0.50 | 7 | - | - |
| | | | | | Placebo | | | | |
| Sorli 2017[96] | Multinational | RCT | 30.0 | 387 | Semaglutide | 0.50 | 4.81 | 54.6 | 47.0 |
| | | | | | Semaglutide | 1.00 | 3.62 | 52.7 | 62.0 |
| | | | | | Placebo | | | | |
| Terauchi 2014[97] | Japan | RCT | 12.0 | 145 | Dulaglutide | 0.25 | 4.3 | 52.3 | 75.0 |
| | | | | | Dulaglutide | 0.50 | 4.9 | 52.5 | 62.2 |
| | | | | | Dulaglutide | 0.75 | 4.6 | 52.2 | 82.0 |
| | | | | | Dulaglutide | 0.50 | 4.9 | 52.5 | 62.2 |
| | | | | | Placebo | | | | |
| Umpierrez 2014[98] | Multinational | RCT | 26.0 | 807 | Dulaglutide | 1.50 | 3 | 56 | 42.0 |
| | | | | | Dulaglutide | 0.75 | 3 | 56 | 44.0 |
| | | | | | Metformin | | | | |
| Urva 2022[99] | USA | RCT | 12.0 | 72 | Dulaglutide | 1.50 | 14.2 | 59.8 | 80.0 |
| | | | | | Retatrutide | 1.50 | 10.7 | 59.2 | 44.0 |
| | | | | | Retatrutide | 3.00 | 10.2 | 56.8 | 33.0 |
| | | | | | Retatrutide | 6.00 | 12.9 | 55.8 | 55.0 |
| | | | | | Retatrutide | 12.00 | 10.0 | 61.5 | 64.0 |
| | | | | | Retatrutide | 4.00 | 12.9 | 55.8 | 67.0 |
| | | | | | Retatrutide | 6.00 | 10.2 | 61.5 | |
| Vanderheiden 2016[100] | Multinational | RCT | 26.0 | 71 | Liraglutide | 1.80 | 16 | 52.8 | 34.0 |
| | | | | | Placebo | | | | |
| Wang 2018[101] | Multinational | RCT | 52.0 | 774 | Dulaglutide | 1.50 | 7.9 | 55 | 53.4 |
| | | | | | Dulaglutide | 0.75 | 8.1 | 54.5 | 56.7 |
| | | | | | InsulinGlargine | | | | |

| Study | Country and region | Design | Follow-up duration (weeks) | Number of participants | Randomised treatment | Dose and frequency | Diabetes duration (years) | Age | Male (%) |
|---|---|---|---|---|---|---|---|---|---|
| Wysham 2016[102] | Multinational | RCT | 20.0 | 110 | Exenatide QMS<br>Exenatide QMS<br>Exenatide QMS<br>ExenatideQW | 5.00<br>8.00<br>11.00 | 5.0<br>6.4<br>6.7 | 50.0<br>52.3<br>49.9 | 57.7<br>82.1<br>55.6 |
| Yabe 2020[103] | Japan | RCT | 52.0 | 458 | Semaglutide<br>Semaglutide<br>Semaglutide<br>Dulaglutide<br>Placebo | 7.00<br>7.00<br>14.00<br>0.75 | 9.3<br>9.1<br>7.9<br>9.9 | 58<br>57<br>61<br>61 | 76.0<br>68.0<br>77.0<br>78.0 |
| Yamada 2020[104] | Japan | RCT | 26.0 | 243 | Semaglutide<br>Semaglutide<br>Semaglutide<br>Liraglutide<br>Placebo | 3.00<br>7.00<br>14.00<br>0.90 | 7.4<br>7.4<br>7.9<br>6.7 | 58<br>60<br>61<br>59 | 73.0<br>73.0<br>83.0<br>81.0 |
| Yan 2024[105] | China | RCT | 52.0 | 273 | Visepegsenatide<br>Placebo | 0.15 | 2.20 | 50.3 | 75.9 |
| Zhaohu 2022[106] | China | RCT | 24.0 | 309 | Dapagliflozin<br>Liraglutide | 1.20 | 6.3 | 51.8 | 61.5 |
| Zinman 2007[107] | Canada | RCT | 64.0 | 233 | Exenatide<br>Placebo | 0.02 | - | 55 | - |
| Zinman 2009[108] | USA, Canada | RCT | 26.0 | 533 | Liraglutide<br>Liraglutide<br>Placebo | 1.80<br>1.20 | 9<br>9 | 55<br>55 | 57.0<br>58.9 |
| Zinman 2019[109] | Multinational | RCT | 30.0 | 302 | Semaglutide<br>Placebo | 1.00 | 9.8 | 57.5 | 58.9 |

Abbreviations: QD, once daily; QW, once weekly; BID, twice daily; QMS, once monthly suspension.

#### Supplementary Table 2. GLP-1RA list

Table 1: GLP-1RA List

| GLP-1RA Drug | Synonyms | Drug type | Company | Highest phase | Drug approved country/region | ATC | Note |
|---|---|---|---|---|---|---|---|
| Albiglutide | GSK-716155, PGC-GLP-1, Tanzeum, Eperzan, Syncria, Albugon | Fusion protein | GSK Plc | Approved | United States, European Union, Canada | A10BJ04 | - |
| CagriSema | Cagrilintide/Semaglutide | Recombinant polypeptide | Novo Nordisk | Phase 3 | - | - | GLP-1/Amylin receptor dual agonist |
| Dulaglutide | GLP-fc, LY-2189265, GLP-1-FC, LY 2189265, Trulicity | Fusion protein | Eli Lilly & Co. | Approved | United States, China, European Union, Japan, Canada, India, Iceland, Liechtenstein, Norway | A10BJ05 | - |
| Efpeglenatide | LAPS-Exendin, Langlenatide, LAPS-exendin 4 analogue, LAPSCA-Exendin4, LAPS-Exendin4, LAPS-EXD4, HM-11260C, SAR-439977 | Fusion protein | Hanmi Pharmaceutical Co., Ltd. | Phase 3 | - | - | - |

| | | | | | | | |
|---|---|---|---|---|---|---|---|
| Exenatide | Presendin, Exendin-4, EX-4, AC-2993, AC-2993-LAR, LY-2148568, LY2148568, Byetta, Bydureon | Synthetic peptide | Eli Lilly & Co.; Amylin Pharmaceuticals, Inc. | Approved | United States, China, European Union, Japan | A10BJ01 | - |
| ITCA 650 | - | Small molecule drug | Intarcia Therapeutics, Inc. | Phase 3 | - | - | Subdermal osmotic exenatide mini-pump |
| Lixisenatide | NN-2211, NN-9211, NNC-90-1170, NN-8022, NNC-901170, NNC 90-1170, NN 9211, NN 2211, LATIN T1D, Saxenda, SAXE, Victoza | Recombinant polypeptide | Novo Nordisk | Approved | United States, China, European Union, Japan, Canada | A10BJ02 | - |
| Liraglutide | des-3-proline-exendin-4 (Heloderma suspectum)-(1-39)-peptidyl-L-lysyl-L-lysinamide, DesPro38Exendin-4(1-39)-Lys6-NH2, ZP-10, AQVE-10010, AVE0100, AVE-0010, ZP10A PEPTIDE, Lyxumia, Adlyxin | Synthetic peptide | Zealand Pharma | Approved | United States, China, European Union, Japan, Canada, Australia, India, Argentina, Brazil, Switzerland, Chile, Colombia, Ecuador, Hong Kong, Israel, Iceland, South Korea, Kuwait, Liechtenstein, Mexico, Norway, Russia, Singapore, Turkey, Taiwan Province, Ukraine | A10BJ03 | - |

| | | | | | | | |
|---|---|---|---|---|---|---|---|
| Mazdutide | IBI-362, IBI362, OXM-3, OXM3, LY3305677, LY-3305677 | Synthetic peptide | Eli Lilly & Co. | Phase 3 | - | - | - |
| Orforglipron | LY3502970, LY-3502970, OWL 833 | Small molecule drug | Eli Lilly & Co. | Phase 3 | - | - | - |
| PEGylated exenatide | PEG-exenatide, PB-119, PB119 | Synthetic peptide | PegBio Co., Ltd | Phase 3 | - | - | - |
| Polyethylene glycol loxenatide | Polyethylene Glycol Loxenatide, PEG loxenatide, pegloxenatide, PEX-168, PEX168 | Synthetic peptide | Jiangsu Hansoh Pharmaceutical Co. Ltd. | Approved | China | A10B | - |
| Retatrutide | LY3437943 | Synthetic peptide | Eli Lilly & Co. | Phase 3 | - | - | GLP-1/GIP/GCG receptor triple agonist |
| Semaglutide | NN-9535, NN-9536, NN-9931, NN9924, NNC-0113-0217, Ozempic, Rybelsus, Wegovy | Synthetic peptide | Novo Nordisk | Approved | United States, China, European Union, Japan | A10BJ06 | - |
| Tirzepatide | LY3298176, LY-3298176, GIP/GLP-1 RA, Mounjaro | Synthetic peptide | Eli Lilly & Co. | Approved | United States, Japan | - | GLP-1/GIP receptor dual agonist |

Abbreviations: ATC: anatomical therapeutic chemical; GIP, gastric inhibitory polypeptide; GCG, glucagon.

#### Supplementary Result 1: PRISMA flow diagram for study selection

***Supplementary Figure 1. Flow diagram of preferred reporting items identified, included, and excluded for systematic reviews and meta-analyses (PRISMA)***[110]

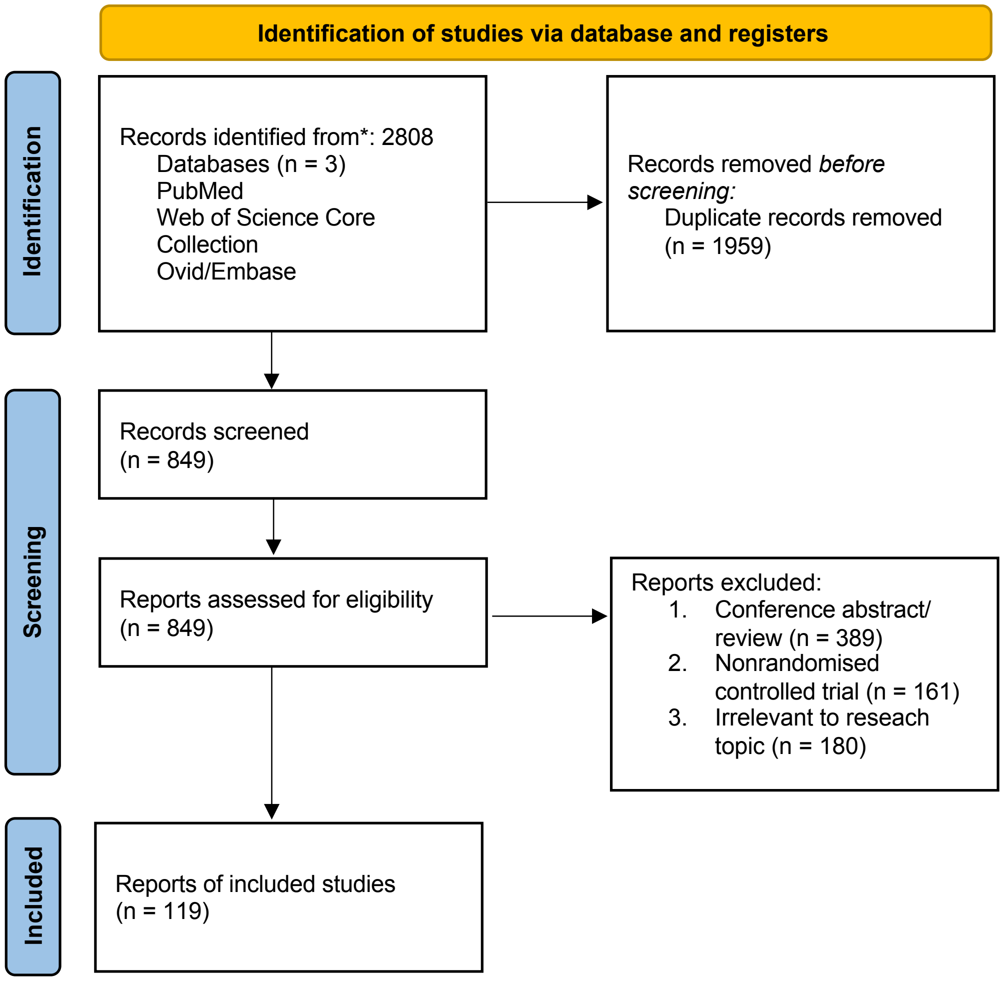

#### Supplementary Result 2: Network maps and forest plots of secondary outcomes

***Supplementary Figure 2. Forest plot of the network analysis of weight loss stratified by drug types***

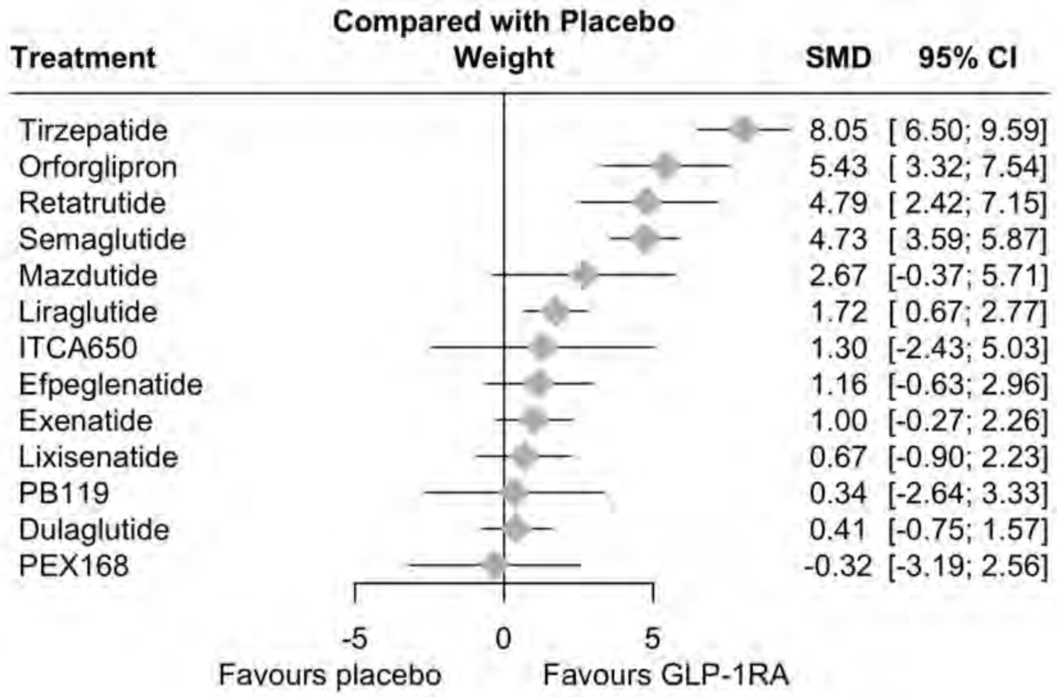

***Supplementary Figure 3. Netgraph of the network analysis of weight loss stratified by drug types***

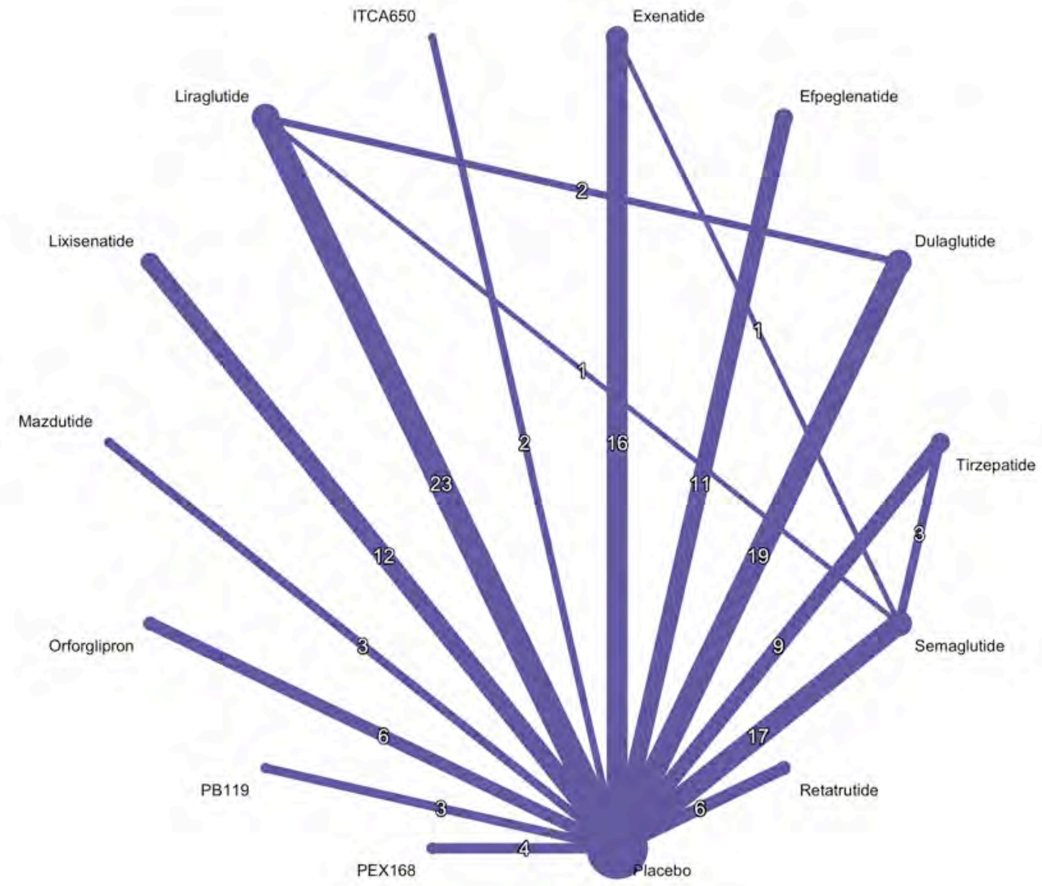

***Supplementary Figure 4. League table of the network analysis of weight loss stratified by drug types***

| | | | | | | | | | | | | | |
|---|---|---|---|---|---|---|---|---|---|---|---|---|---|
| **Tirzepatide** | | | | | | | | | | | | | |
| 2.61 (0.00, 5.23) | **Orforglipron** | | | | | | | | | | | | |
| 3.26 (0.43, 6.08) | 0.65 (-2.52, 3.81) | **Retatrutide** | | | | | | | | | | | |
| 3.31 (1.57, 5.06) | 0.70 (-1.70, 3.10) | 0.05 (-2.57, 2.68) | **Semaglutide** | | | | | | | | | | |
| 5.38 (1.97, 8.79) | 2.76 (-0.94, 6.46) | 2.12 (-1.73, 5.97) | 2.06 (-1.18, 5.31) | **Mazdutide** | | | | | | | | | |
| 6.33 (4.47, 8.19) | 3.72 (1.36, 6.07) | 3.07 (0.48, 5.66) | 3.01 (1.50, 4.53) | 0.95 (-2.26, 4.17) | **Liraglutide** | | | | | | | | |
| 6.75 (2.71, 10.78) | 4.13 (-0.15, 8.42) | 3.49 (-0.93, 7.90) | 3.43 (-0.46, 7.33) | 1.37 (-3.44, 6.18) | 0.42 (-3.45, 4.29) | **ITCA650** | | | | | | | |
| 6.88 (4.51, 9.25) | 4.27 (1.50, 7.04) | 3.62 (0.66, 6.59) | 3.57 (1.44, 5.69) | 1.51 (-2.02, 5.04) | 0.55 (-1.52, 2.63) | 0.14 (-4.00, 4.27) | **Efpeglenatide** | | | | | | |
| 7.05 (5.06, 9.04) | 4.43 (1.97, 6.90) | 3.79 (1.11, 6.47) | 3.73 (2.08, 5.39) | 1.67 (-1.62, 4.96) | 0.72 (-0.92, 2.36) | 0.30 (-3.64, 4.24) | 0.16 (-2.03, 2.36) | **Exenatide** | | | | | |
| 7.38 (5.18, 9.58) | 4.76 (2.13, 7.39) | 4.12 (1.28, 6.95) | 4.06 (2.13, 6.00) | 2.00 (-1.42, 5.42) | 1.05 (-0.84, 2.93) | 0.63 (-3.41, 4.67) | 0.49 (-1.89, 2.87) | 0.33 (-1.68, 2.34) | **Lixisenatide** | | | | |
| 7.70 (4.34, 11.06) | 5.09 (1.44, 8.74) | 4.44 (0.64, 8.25) | 4.39 (1.20, 7.58) | 2.33 (-1.93, 6.58) | 1.37 (-1.79, 4.53) | 0.96 (-3.82, 5.73) | 0.82 (-2.66, 4.30) | 0.66 (-2.58, 3.89) | 0.33 (-3.04, 3.69) | **PB119** | | | |
| 7.64 (5.71, 9.57) | 5.02 (2.62, 7.43) | 4.38 (1.75, 7.01) | 4.32 (2.70, 5.94) | 2.26 (-0.99, 5.51) | 1.31 (-0.18, 2.80) | 0.89 (-3.01, 4.79) | 0.76 (-1.38, 2.89) | 0.59 (-1.12, 2.31) | 0.26 (-1.69, 2.21) | -0.06 (-3.26, 3.13) | **Dulaglutide** | | |
| 8.36 (5.09, 11.63) | 5.75 (2.18, 9.32) | 5.10 (1.38, 8.83) | 5.05 (1.95, 8.14) | 2.98 (-1.20, 7.17) | 2.03 (-1.03, 5.10) | 1.61 (-3.10, 6.32) | 1.48 (-1.91, 4.87) | 1.31 (-1.83, 4.46) | 0.98 (-2.29, 4.26) | 0.66 (-3.49, 4.80) | 0.72 (-2.38, 3.83) | **PEX168** | |
| 8.05 (6.50, 9.59) | 5.43 (3.32, 7.54) | 4.79 (2.42, 7.15) | 4.73 (3.59, 5.87) | 2.67 (-0.37, 5.71) | 1.72 (0.67, 2.77) | 1.30 (-2.43, 5.03) | 1.16 (-0.63, 2.96) | 1.00 (-0.27, 2.26) | 0.67 (-0.90, 2.23) | 0.34 (-2.64, 3.33) | 0.41 (-0.75, 1.57) | -0.32 (-3.19, 2.56) | **Placebo** |

*Supplementary Figure 5. Forest plot of the network analysis of hba1c stratified by drug types*

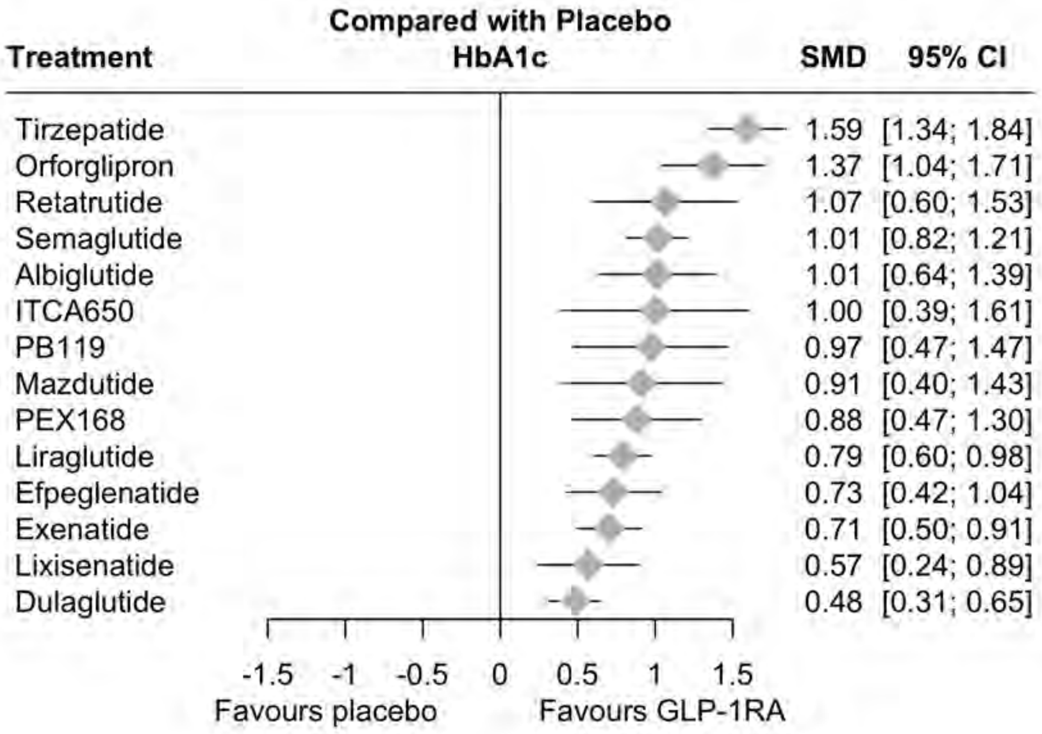

***Supplementary Figure 6. Netgraph of the network analysis of hba1c loss stratified by drug types***

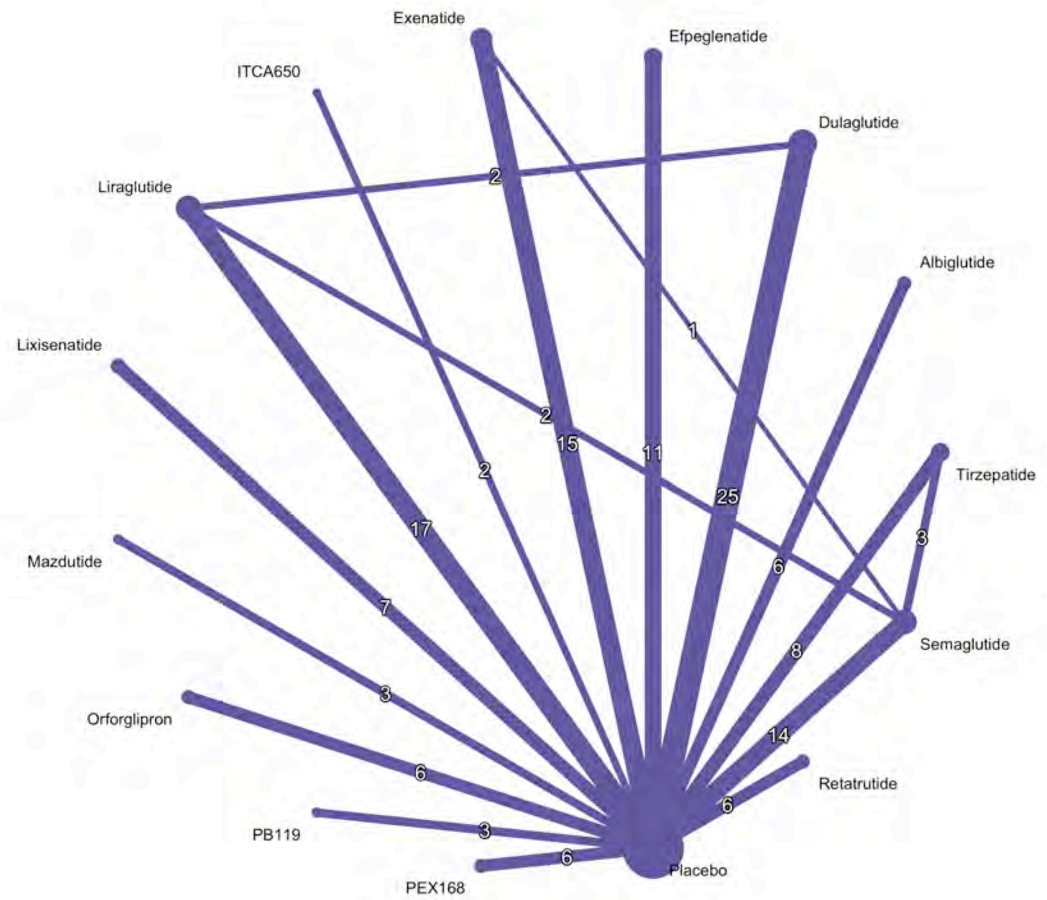

***Supplementary Figure 7. League table of the network analysis of HbA1c stratified by drug types***

| | | | | | | | | | | | | | | |
|---|---|---|---|---|---|---|---|---|---|---|---|---|---|---|
| Tirzepatide | | | | | | | | | | | | | | |
| 0.22 (-0.20, 0.64) | Orforglipron | | | | | | | | | | | | | |
| 0.53 (0.00, 1.05) | 0.31 (-0.26, 0.88) | Retatrutide | | | | | | | | | | | | |
| 0.58 (0.29, 0.86) | 0.36 (-0.03, 0.75) | 0.05 (-0.45, 0.55) | Semaglutide | | | | | | | | | | | |
| 0.58 (0.13, 1.03) | 0.36 (-0.14, 0.86) | 0.05 (-0.54, 0.65) | 0.00 (-0.42, 0.42) | Albiglutide | | | | | | | | | | |
| 0.59 (-0.07, 1.25) | 0.37 (-0.32, 1.07) | 0.07 (-0.70, 0.83) | 0.01 (-0.62, 0.65) | 0.01 (-0.70, 0.73) | ITCA650 | | | | | | | | | |
| 0.62 (0.06, 1.18) | 0.40 (-0.20, 1.00) | 0.09 (-0.59, 0.77) | 0.04 (-0.49, 0.58) | 0.04 (-0.58, 0.67) | 0.03 (-0.76, 0.81) | PB119 | | | | | | | | |
| 0.68 (0.11, 1.25) | 0.46 (-0.15, 1.07) | 0.15 (-0.54, 0.84) | 0.10 (-0.45, 0.65) | 0.10 (-0.54, 0.73) | 0.09 (-0.71, 0.88) | 0.06 (-0.66, 0.77) | Mazdutide | | | | | | | |
| 0.71 (0.22, 1.19) | 0.49 (-0.04, 1.02) | 0.18 (-0.44, 0.80) | 0.13 (-0.33, 0.59) | 0.13 (-0.43, 0.69) | 0.12 (-0.62, 0.85) | 0.09 (-0.56, 0.74) | 0.03 (-0.63, 0.69) | PEX168 | | | | | | |
| 0.80 (0.49, 1.11) | 0.58 (0.20, 0.97) | 0.27 (-0.22, 0.77) | 0.22 (-0.03, 0.48) | 0.22 (-0.20, 0.64) | 0.21 (-0.43, 0.84) | 0.18 (-0.35, 0.71) | 0.12 (-0.42, 0.67) | 0.09 (-0.36, 0.55) | Liraglutide | | | | | |
| 0.86 (0.46, 1.26) | 0.64 (0.19, 1.10) | 0.34 (-0.22, 0.89) | 0.28 (-0.08, 0.65) | 0.28 (-0.20, 0.77) | 0.27 (-0.41, 0.95) | 0.24 (-0.34, 0.83) | 0.18 (-0.41, 0.78) | 0.15 (-0.36, 0.67) | 0.06 (-0.30, 0.42) | Efpeglenatide | | | | |
| 0.89 (0.56, 1.21) | 0.67 (0.27, 1.06) | 0.36 (-0.15, 0.87) | 0.31 (0.03, 0.59) | 0.31 (-0.12, 0.74) | 0.29 (-0.35, 0.94) | 0.27 (-0.28, 0.81) | 0.21 (-0.35, 0.76) | 0.18 (-0.29, 0.64) | 0.09 (-0.20, 0.37) | 0.02 (-0.35, 0.40) | Exenatide | | | |
| 1.02 (0.61, 1.44) | 0.81 (0.34, 1.27) | 0.50 (-0.07, 1.06) | 0.45 (0.07, 0.83) | 0.45 (-0.05, 0.94) | 0.43 (-0.26, 1.12) | 0.40 (-0.19, 1.00) | 0.35 (-0.26, 0.95) | 0.32 (-0.21, 0.84) | 0.22 (-0.15, 0.60) | 0.16 (-0.29, 0.61) | 0.14 (-0.25, 0.53) | Lixisenatide | | |
| 1.11 (0.81, 1.41) | 0.89 (0.52, 1.27) | 0.58 (0.09, 1.07) | 0.53 (0.28, 0.79) | 0.53 (0.12, 0.94) | 0.52 (-0.11, 1.15) | 0.49 (-0.04, 1.02) | 0.43 (-0.11, 0.97) | 0.40 (-0.04, 0.85) | 0.31 (0.07, 0.55) | 0.25 (-0.10, 0.60) | 0.22 (-0.04, 0.49) | 0.09 (-0.28, 0.45) | Dulaglutide | |
| 1.59 (1.34, 1.84) | 1.37 (1.04, 1.71) | 1.07 (0.60, 1.53) | 1.01 (0.82, 1.21) | 1.01 (0.64, 1.39) | 1.00 (0.39, 1.61) | 0.97 (0.47, 1.47) | 0.91 (0.40, 1.43) | 0.88 (0.47, 1.30) | 0.79 (0.60, 0.98) | 0.73 (0.42, 1.04) | 0.71 (0.50, 0.91) | 0.57 (0.24, 0.89) | 0.48 (0.31, 0.65) | Placebo |

#### Supplementary Result 3: Funnel plots

***Supplementary Figure 8. Funnel plot of SBP***

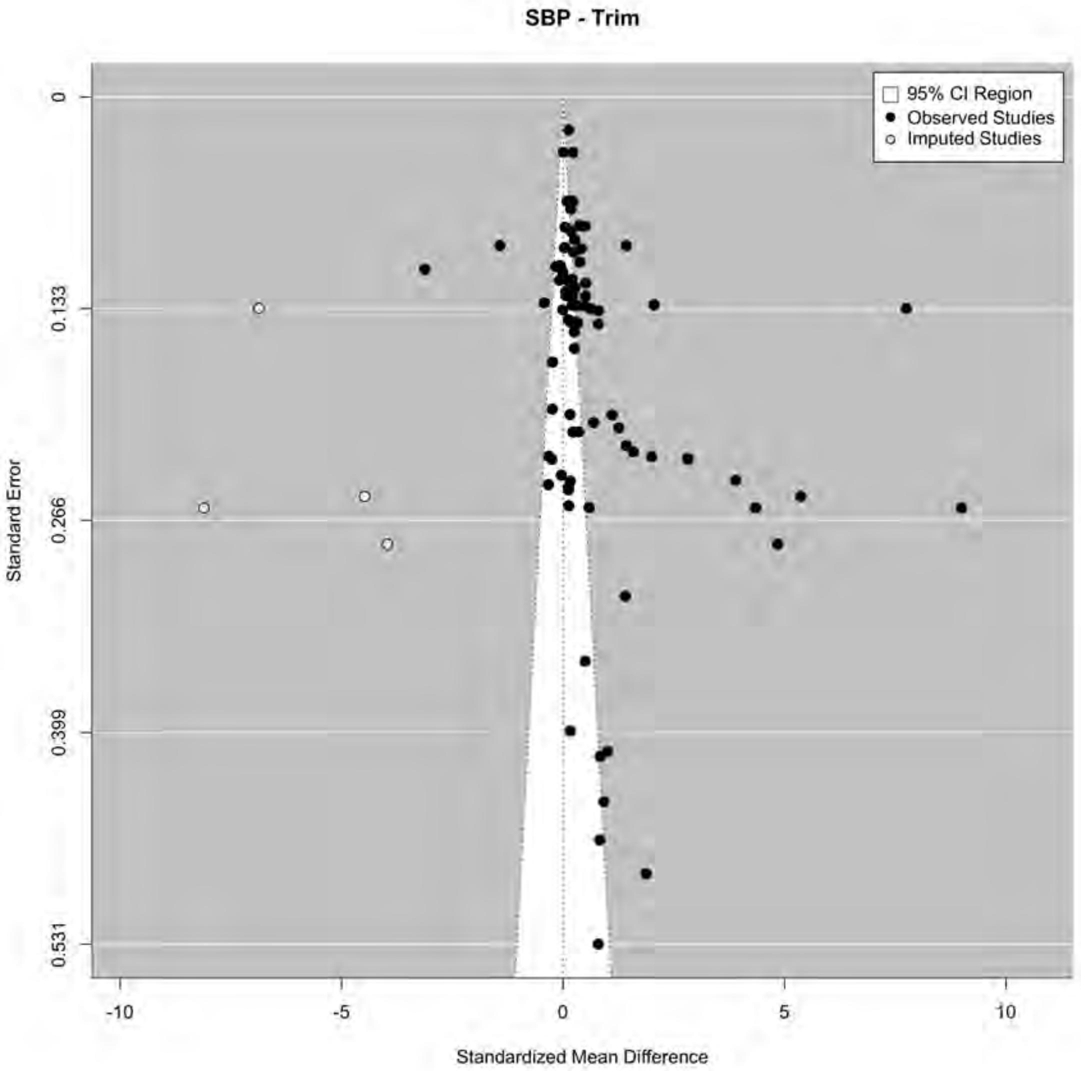

Egger's test: z = 2.7228, p = 0.0065 b = -0.0370 (CI: -0.6681, 0.5941)
Begg's test: Kendall's tau = 0.3261, p < .0001
Trimfill:
Egger's test: z = 1.4155, p = 0.1569 b = -0.0724 (CI: -0.9066, 0.7619)
Begg's test: Kendall's tau = 0.0172, p = 0.9115

*Supplementary Figure 9. Funnel plot of DBP*

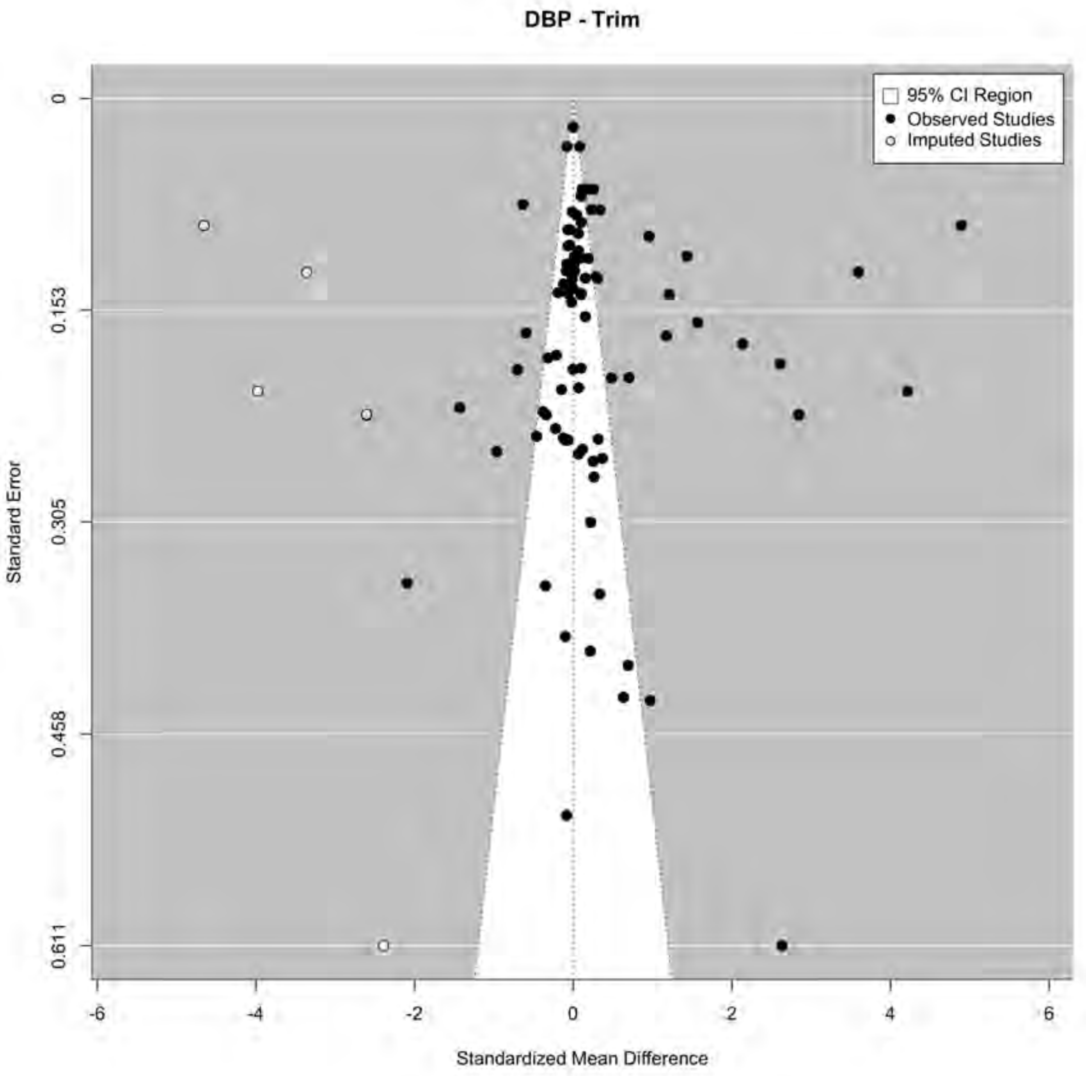

Egger's test: z = 0.3674, p = 0.7133 b = 0.2673 (CI: -0.1666, 0.7013)
Begg's test: Kendall's tau = 0.1596, p = 0.0259
**Trimfill:**
Egger's test: z = -0.1498, p = 0.8809 b = 0.1510 (CI: -0.3779, 0.6800)
Kendall's tau = 0.0110, p = 0.8742

***Supplementary Figure 10. Funnel plot of weight loss***

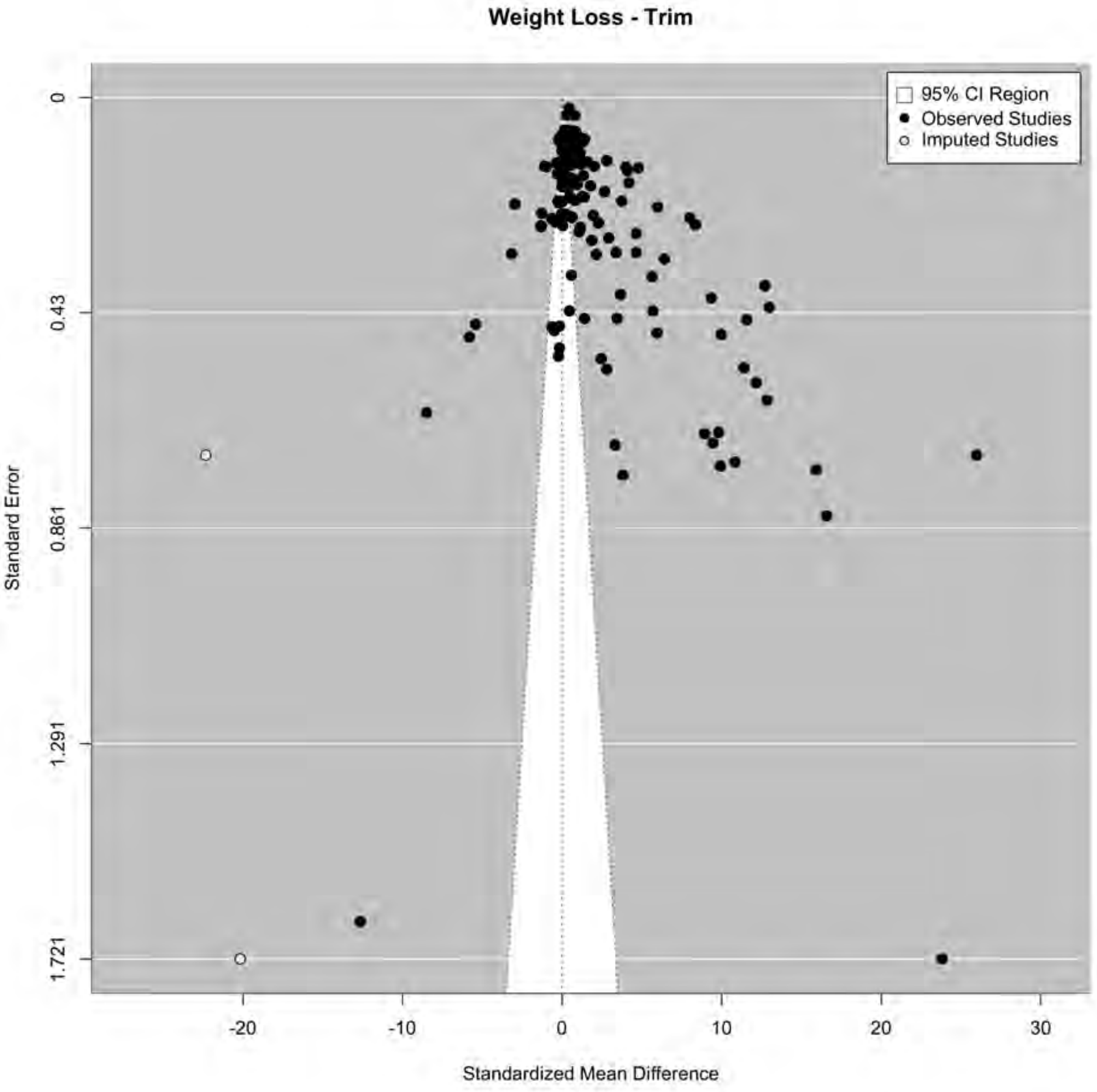

Egger's test: z = 6.8760, p < .0001 b = -0.3141 (CI: -1.2265, 0.5984)
Begg's test: Kendall's tau = 0.2559, p < .0001
**Trimfill:**
Egger's test: z = 2.5004, p = 0.0124 b = 0.7907 (CI: -0.3221, 1.9035)
Kendall's tau = 0.2272, p < .0001

***Supplementary Figure 11. Funnel plot of HbA1c***

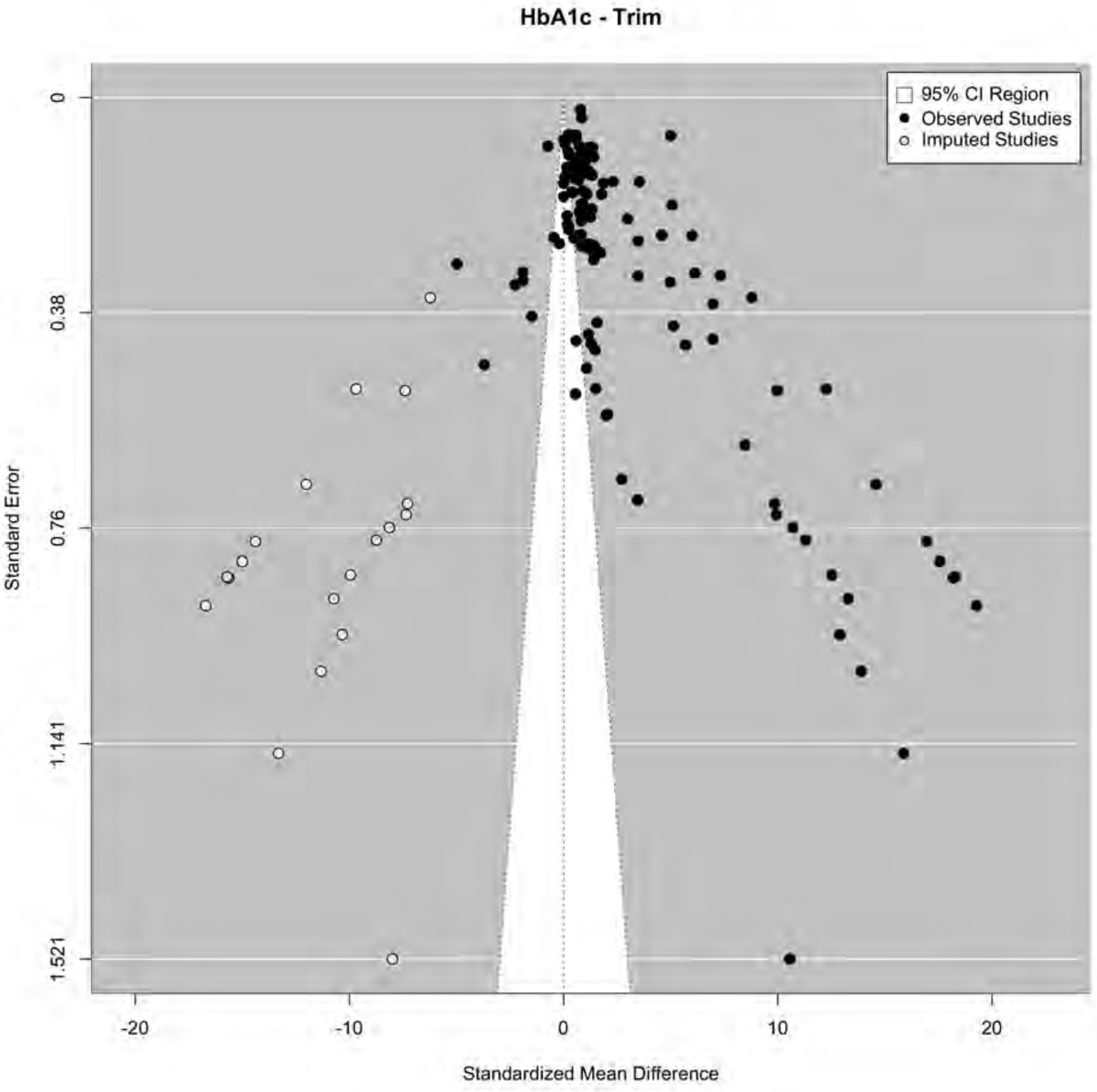

Egger's test: z = 14.8171, p < .0001 b = -1.4139 (CI: -2.1648, -0.6631)
Begg's test: Kendall's tau = 0.3955, p < .0001
**Trimfill:**
Egger's test: z = 0.3763, p = 0.7067 b = 1.0545 (CI: -0.5282, 2.6373)
Kendall's tau = 0.1131, p = 0.0358

#### Supplementary Result 4: Sensitivity analysis

Supplementary Table 3. Sensitivity analyses of primary outcomes with all trials comparing GLP-1 receptor against other classes of glucose-lowering drugs.

| GLP-1 RA | Weight | | HbA1c (%) | | SBP | | DBP | |
|---|---|---|---|---|---|---|---|---|
| | Main estimate | Sensitivity analyses | Main estimate | Sensitivity analyses | Main estimate | Sensitivity analyses | Main estimate | Sensitivity analyses |
| Tirzepatide | 8.05 (6.50, 9.59) | 8.04 (6.83, 9.26) | 1.59 (1.34, 1.84) | 1.62 (1.41, 1.84) | 5.18 (3.51, 6.85) | 7.81 (5.94, 9.68) | 1.73 (0.88, 2.57) | 2.28 (1.24, 3.32) |
| Orforglipron | 5.43 (3.32, 7.54) | 5.44 (3.34, 7.54) | 1.37 (1.04, 1.71) | 1.37 (1.11, 1.63) | 2.51 (0.50, 4.52) | 2.20 (0.68, 3.73) | 0.06 (-0.73, 0.85) | -0.12 (-0.90, 0.66) |
| Retatrutide | 4.79 (2.42, 7.15) | 4.82 (3.31, 6.32) | 1.07 (0.60, 1.53) | 1.06 (0.65, 1.47) | 6.98 (3.45, 10.50) | 6.89 (3.69, 10.09) | 1.38 (-0.38, 3.15) | 1.35 (-0.30, 2.99) |
| Semaglutide | 4.73 (3.59, 5.87) | 3.86 (2.22, 5.51) | 1.01 (0.82, 1.21) | 1.14 (0.77, 1.50) | 3.39 (2.07, 4.71) | 2.79 (1.50, 4.08) | 0.84 (0.24, 1.44) | 1.07 (0.55, 1.59) |
| Albiglutide | NA | NA | 1.01 (0.64, 1.39) | 1.10 (0.69, 1.31) | 1.76 (-1.65, 5.17) | 1.62 (-0.43, 3.68) | 0.90 (-1.27, 3.07) | 0.90 (-1.02, 2.82) |
| ITCA650 | 1.30 (-2.43, 5.03) | 1.30 (-1.94, 4.54) | 1.00 (0.39, 1.61) | 1.00 (0.47, 1.53) | 1.70 (-2.37, 5.77) | 1.71 (-0.65, 4.06) | 0.30 (-1.56, 2.17) | 0.30 (-1.25, 1.86) |
| Mazdutide | 2.67 (-0.37, 5.71) | 2.67 (1.02, 4.33) | 0.91 (0.40, 1.43) | 0.91 (0.49, 1.33) | NA | NA | NA | NA |
| Liraglutide | 1.72 (0.67, 2.77) | 0.42 (-1.33, 2.17) | 0.79 (0.60, 0.98) | 0.37 (-0.02, 0.76) | 2.67 (1.45, 3.88) | 1.69 (0.33, 3.05) | 0.49 (-0.07, 1.05) | 0.52 (-1.14, 2.17) |
| Efpeglenatide | 1.16 (-0.63, 2.96) | 1.19 (-0.40, 2.29) | 0.73 (0.42, 1.04) | 0.73 (0.48, 0.98) | 3.55 (-0.88, 7.98) | 3.56 (0.05, 7.07) | -0.71 (-3.04, 1.62) | -0.71 (-3.00, 1.59) |
| Exenatide | 1.00 (-0.27, 2.26) | 0.16 (-3.08, 3.40) | 0.71 (0.50, 0.91) | 0.54 (-0.22, 1.29) | 1.48 (-0.12, 3.08) | 0.39 (-2.12, 2.90) | 0.56 (-0.16, 1.27) | -0.89 (-2.99, 1.21) |
| Lixisenatide | 0.67 (-0.90, 2.23) | 0.73 (-0.70, 2.17) | 0.57 (0.24, 0.89) | 0.58 (0.25, 0.90) | NA | NA | NA | NA |
| PB119 | 0.34 (-2.64, 3.33) | 0.34 (-2.23, 2.97) | 0.97 (0.47, 1.47) | 0.97 (0.53, 1.41) | NA | NA | NA | NA |
| Dulaglutide | 0.41 (-0.75, 1.57) | 0.57 (-0.32, 1.47) | 0.48 (0.31, 0.65) | 0.48 (0.34, 0.62) | 1.69 (0.13, 3.24) | 1.47 (0.48, 2.46) | 0.35 (-0.20, 0.89) | -0.06 (-0.53, 0.40) |
| PEX168 | -0.32 (-3.19, 2.56) | -0.31 (-2.86, 2.23) | 0.88 (0.47, 1.30) | 0.88 (0.52, 1.24) | -1.22 (-3.51, 1.06) | -1.00 (-2.74, 0.74) | -0.03 (-1.05, 1.00) | -0.03 (-0.93, 0.87) |

Supplementary Table 4. Sensitivity analyses of primary outcomes after removing high heterougenity stuides

| GLP-1 RA | Weight | | HbA1c (%) | | SBP | | DBP | |
|---|---|---|---|---|---|---|---|---|
| | Main estimate | Sensitivity analyses | Main estimate | Sensitivity analyses | Main estimate | Sensitivity analyses | Main estimate | Sensitivity analyses |
| Tirzepatide | 8.05 (6.50, 9.59) | 7.65 (6.74, 8.57) | 1.59 (1.34, 1.84) | 1.62 (1.41, 1.84) | 5.18 (3.51, 6.85) | 5.01 (4.03, 5.98) | 1.73 (0.88, 2.57) | 1.45 (0.60, 2.31) |
| Orforglipron | 5.43 (3.32, 7.54) | 5.44 (3.34, 6.54) | 1.37 (1.04, 1.71) | 1.37 (1.11, 1.63) | 2.51 (0.50, 4.52) | 2.30 (1.78, 2.81) | 0.06 (-0.73, 0.85) | 0.00 (-0.51, 0.52) |
| Retatrutide | 4.79 (2.42, 7.15) | 4.82 (3.31, 6.32) | 1.07 (0.60, 1.53) | 1.06 (0.65, 1.47) | 6.98 (3.45, 10.50) | 6.76 (3.88, 9.64) | 1.38 (-0.38, 3.15) | 1.35 (-0.30, 2.99) |
| Semaglutide | 4.73 (3.59, 5.87) | 3.86 (2.22, 5.51) | 1.01 (0.82, 1.21) | 1.14 (0.77, 1.50) | 3.39 (2.07, 4.71) | 2.79 (1.50, 4.08) | 0.84 (0.24, 1.44) | 0.01 (-1.22, 1.24) |
| Albiglutide | NA | NA | 1.01 (0.64, 1.39) | 1.10 (0.69, 1.31) | 1.76 (-1.65, 5.17) | 1.62 (-0.43, 3.68) | 0.90 (-1.27, 3.07) | 0.90 (-1.02, 2.82) |
| ITCA650 | 1.30 (-2.43, 5.03) | 1.30 (-0.74, 3.33) | 1.00 (0.39, 1.61) | 1.00 (0.51, 1.49) | 1.70 (-2.37, 5.77) | 1.71 (-0.65, 4.06) | 0.30 (-1.56, 2.17) | 0.30 (-1.25, 1.86) |
| Mazdutide | 2.67 (-0.37, 5.71) | 2.67 (1.02, 4.33) | 0.91 (0.40, 1.43) | 0.91 (0.49, 1.33) | NA | NA | NA | NA |
| Liraglutide | 1.72 (0.67, 2.77) | 0.42 (-1.33, 2.17) | 0.79 (0.60, 0.98) | 0.37 (-0.02, 0.76) | 2.67 (1.45, 3.88) | 1.69 (0.33, 3.05) | 0.49 (-0.07, 1.05) | 0.52 (-1.14, 2.17) |
| Efpeglenatide | 1.16 (-0.63, 2.96) | 1.25 (0.21, 2.29) | 0.73 (0.42, 1.04) | 0.73 (0.48, 0.98) | 3.55 (-0.88, 7.98) | 3.56 (0.05, 7.07) | -0.71 (-3.04, 1.62) | -0.71 (-2.88, 1.47) |
| Exenatide | 1.00 (-0.27, 2.26) | 0.16 (-3.08, 3.40) | 0.71 (0.50, 0.91) | 0.54 (-0.22, 1.29) | 1.48 (-0.12, 3.08) | 0.39 (-2.12, 2.90) | 0.56 (-0.16, 1.27) | -0.89 (-2.99, 1.21) |
| Lixisenatide | 0.67 (-0.90, 2.23) | 0.65 (-0.22, 1.52) | 0.57 (0.24, 0.89) | 0.57 (0.31, 0.83) | NA | NA | NA | NA |
| PB119 | 0.34 (-2.64, 3.33) | 0.34 (-1.20, 1.89) | 0.97 (0.47, 1.47) | 0.97 (0.57, 1.37) | NA | NA | NA | NA |
| Dulaglutide | 0.41 (-0.75, 1.57) | 0.32 (-0.33, 0.97) | 0.48 (0.31, 0.65) | 0.43 (0.30, 0.57) | 1.69 (0.13, 3.24) | 1.23 (0.60, 1.87) | 0.35 (-0.20, 0.89) | 0.43 (0.00, 0.85) |
| PEX168 | -0.32 (-3.19, 2.56) | -0.31 (-1.97, 1.35) | 0.88 (0.55, 1.21) | 0.88 (0.47, 1.30) | -1.22 (-3.51, 1.06) | -0.16 (-0.84, 0.53) | -0.03 (-1.05, 1.00) | 0.32 (-0.41, 1.04) |

### Supplementary Result 5: Meta-regression analysis

***Supplementary Figure 12. Meta-regression of the effects of GLP1a on systolic blood pressure stratified by diabetes duration***

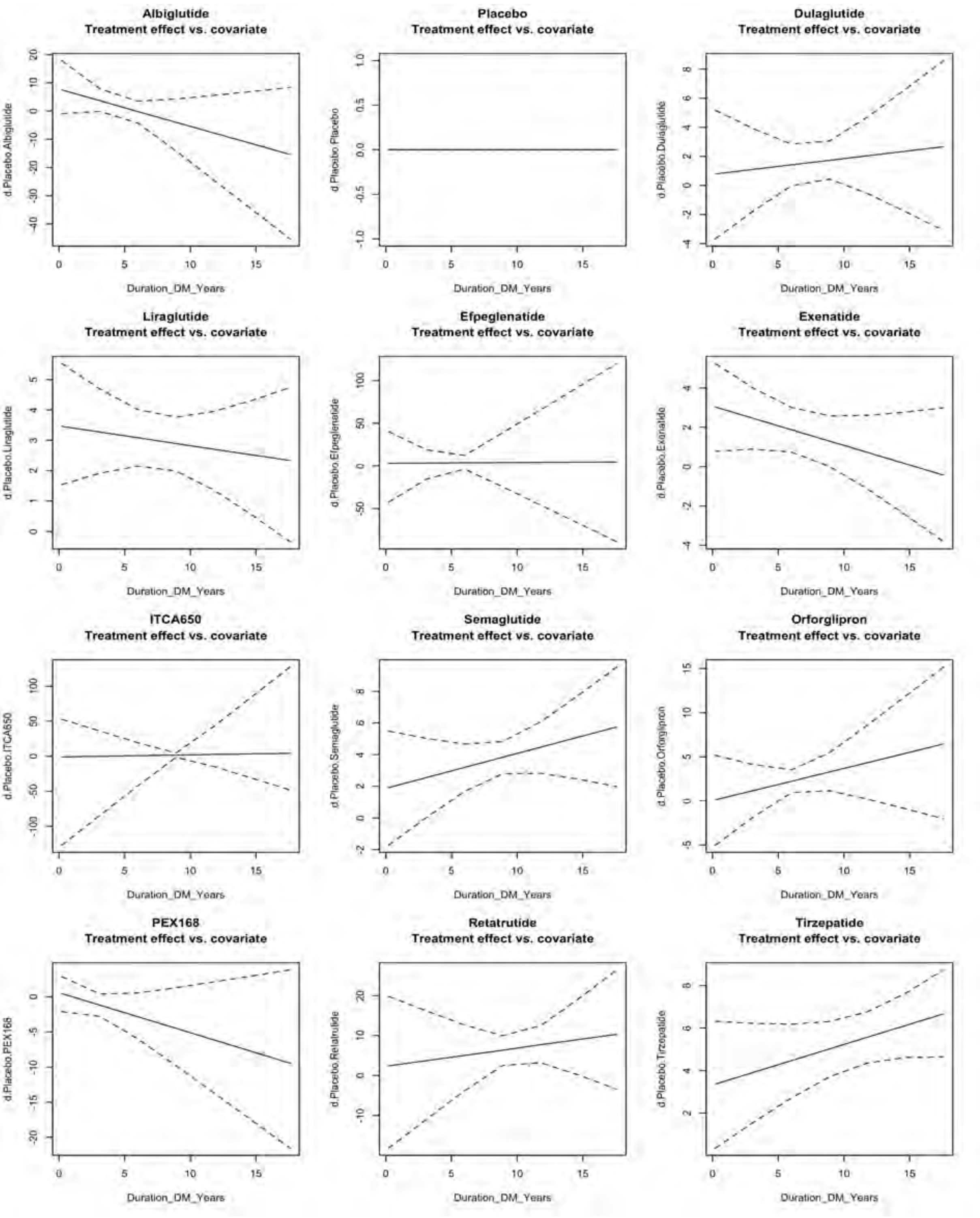

*Supplementary Figure 13. Meta-regression of the effects of GLP1a on systolic blood pressure stratified by age*

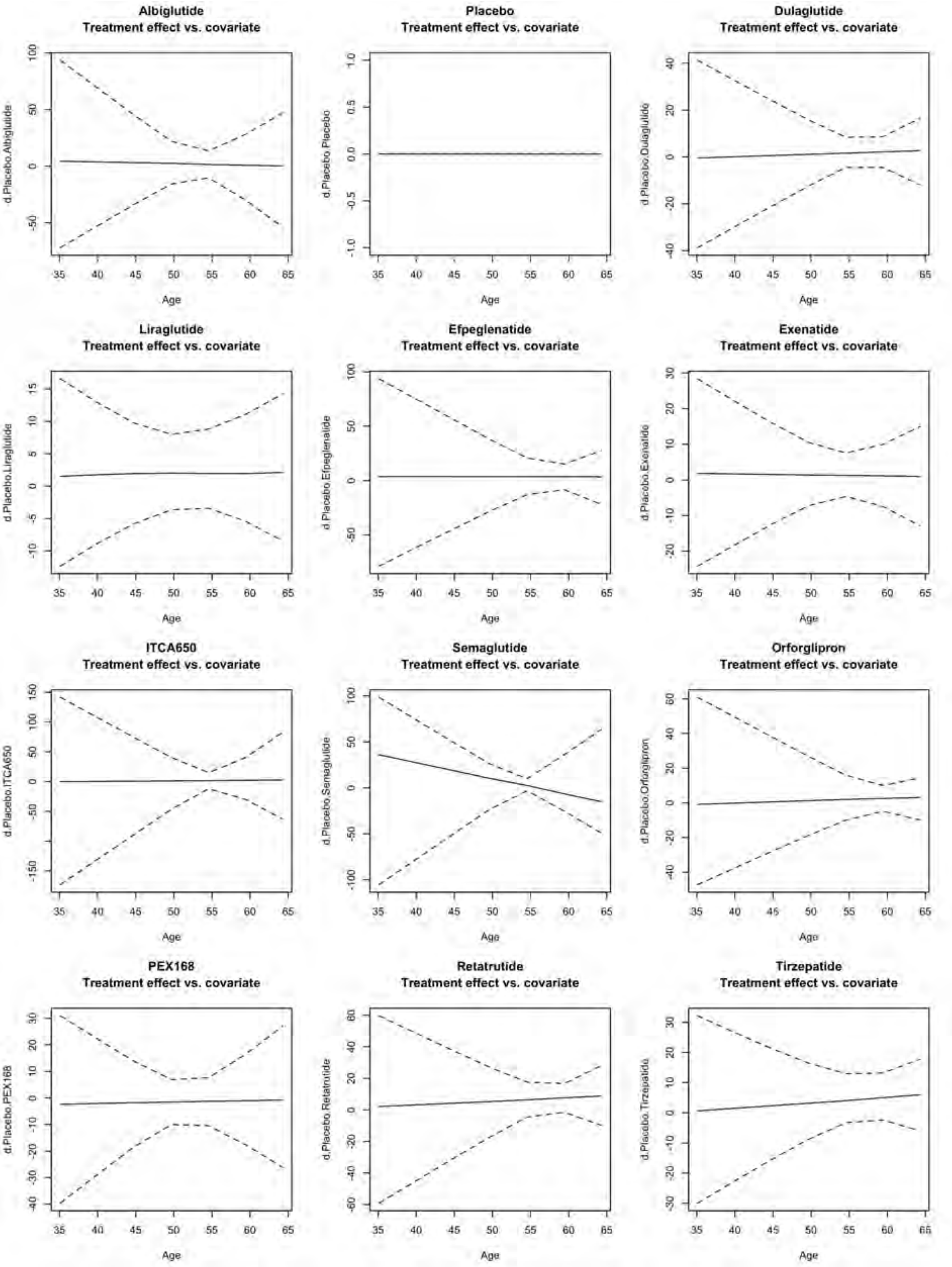

***Supplementary Figure 14. Meta-regression of the effects of GLP1a on systolic blood pressure stratified by weight changes***

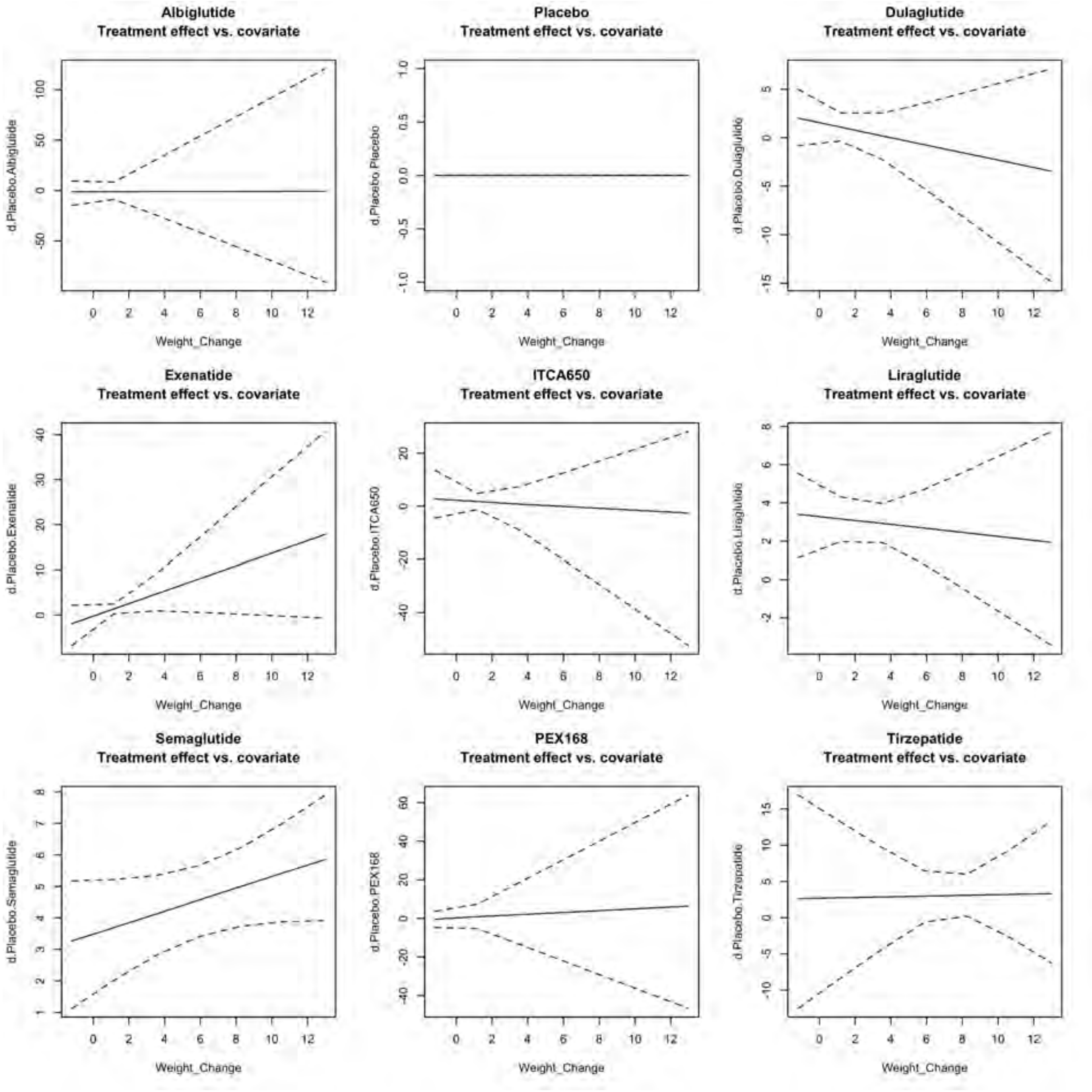

*Supplementary Figure 15. Meta-regression of the effects of GLP1a on diastolic blood pressure stratified by weight changes*

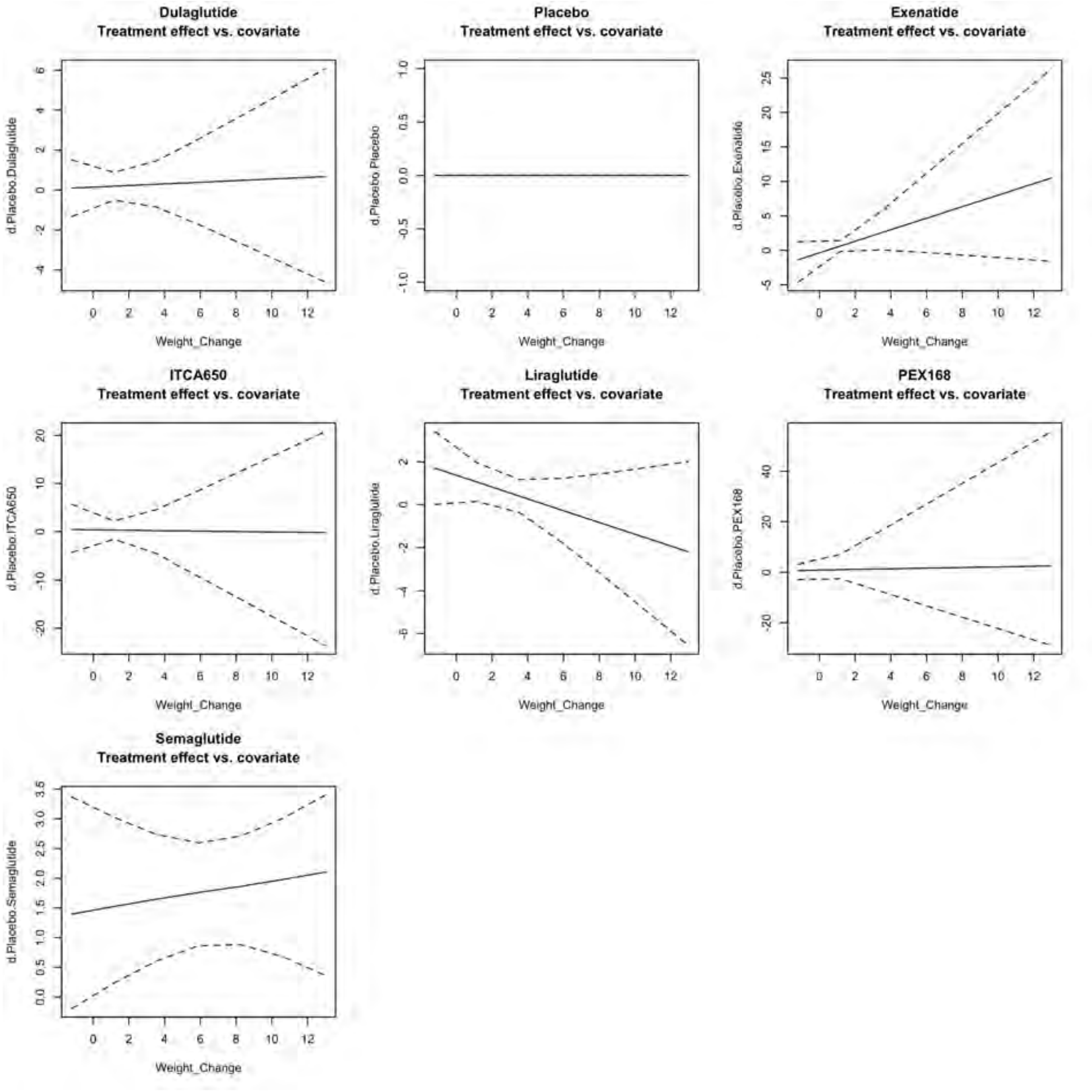

#### Supplementary Result 6: Mediation analysis

***Supplementary Figure 16. Mediation analysis of the GLP1a effects on systolic blood pressure mediated by weight changes***

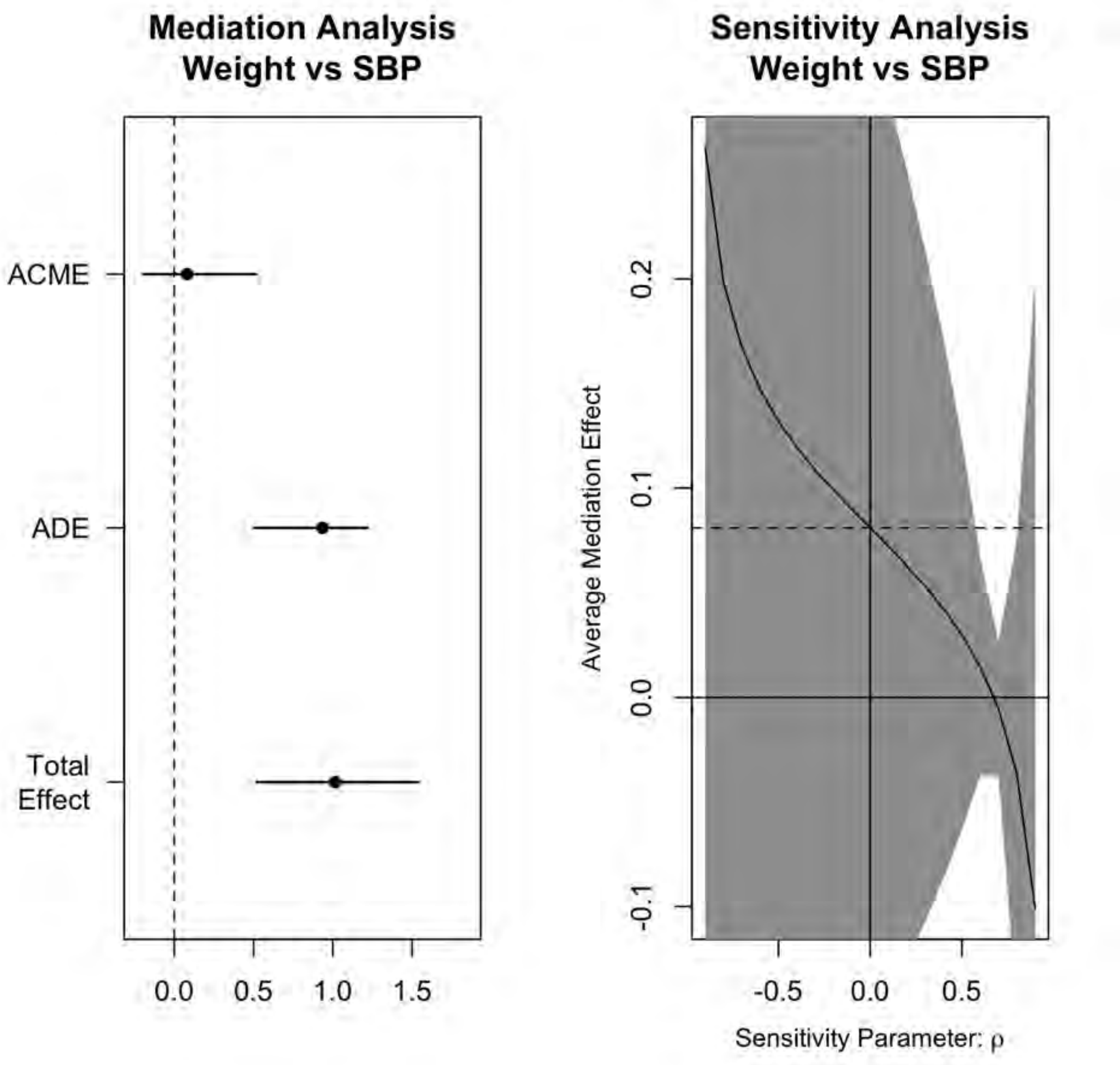

*Supplementary Figure 17. Mediation analysis of the GLP1a effects on systolic blood pressure mediated by HbA1c*

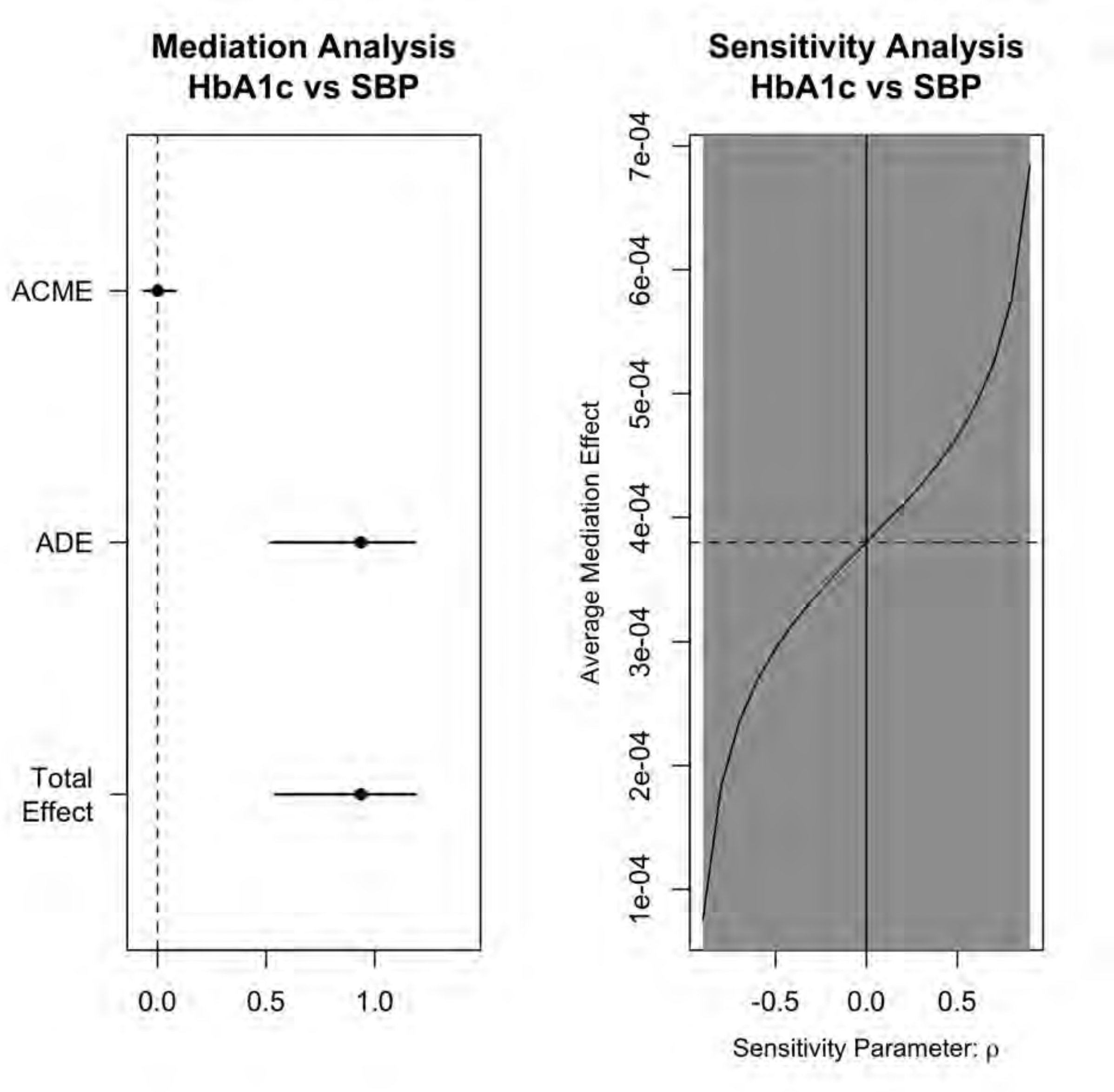

***Supplementary Figure 18. Bayes model of the mediation effects of GLP1a on systolic blood pressure***

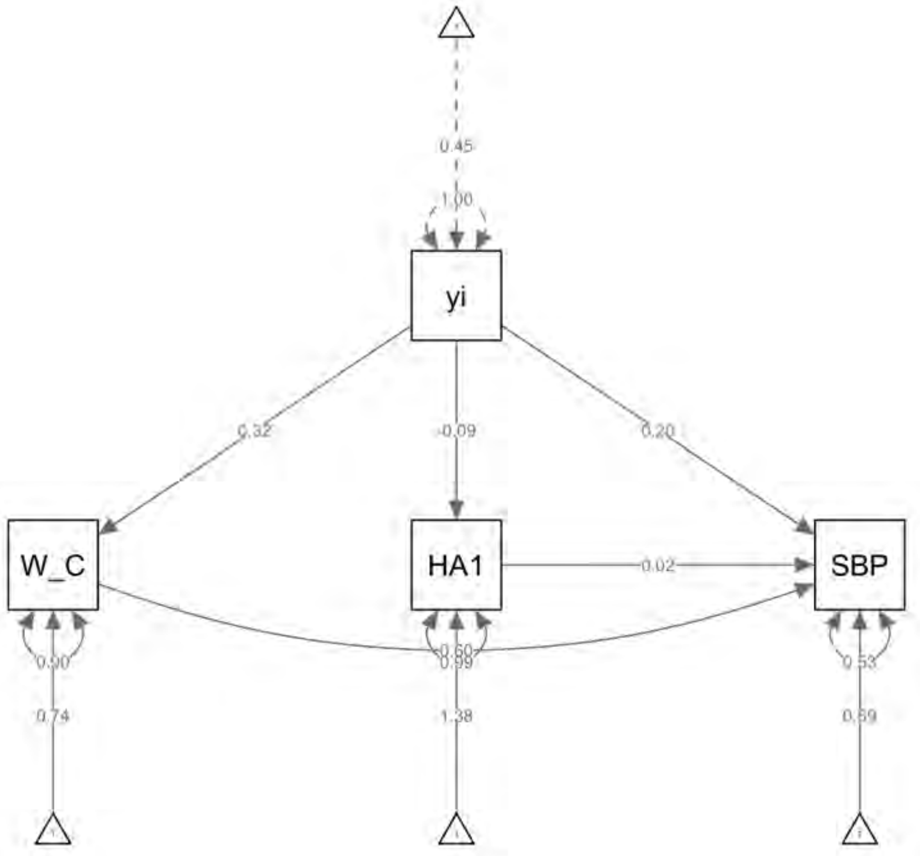

*Supplementary Figure 19. Mediation analysis of the GLP1a effects on diastolic blood pressure mediated by weight changes*

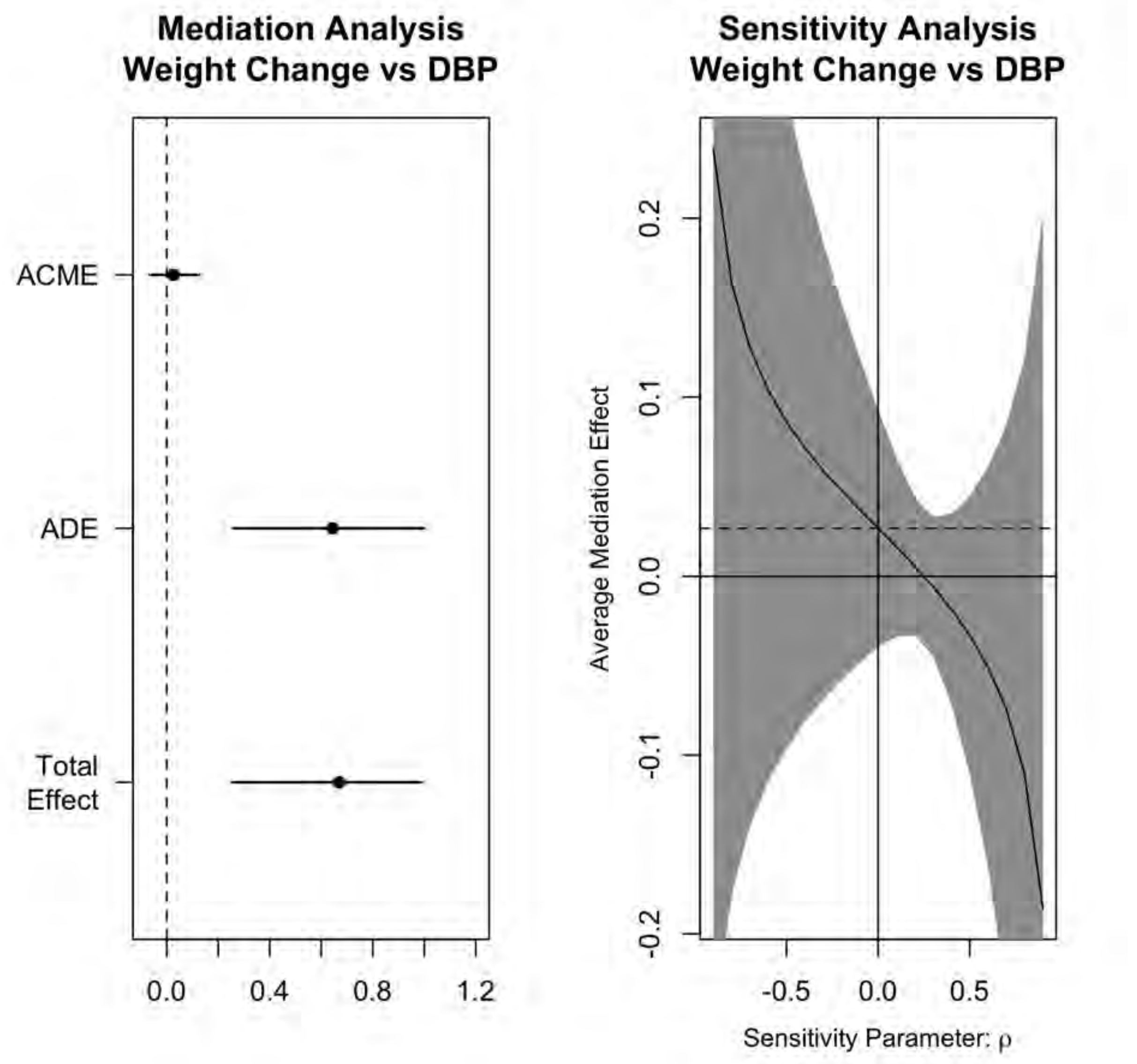

*Supplementary Figure 20. Mediation analysis of the GLP1a effects on diastolic blood pressure mediated by HbA1c*

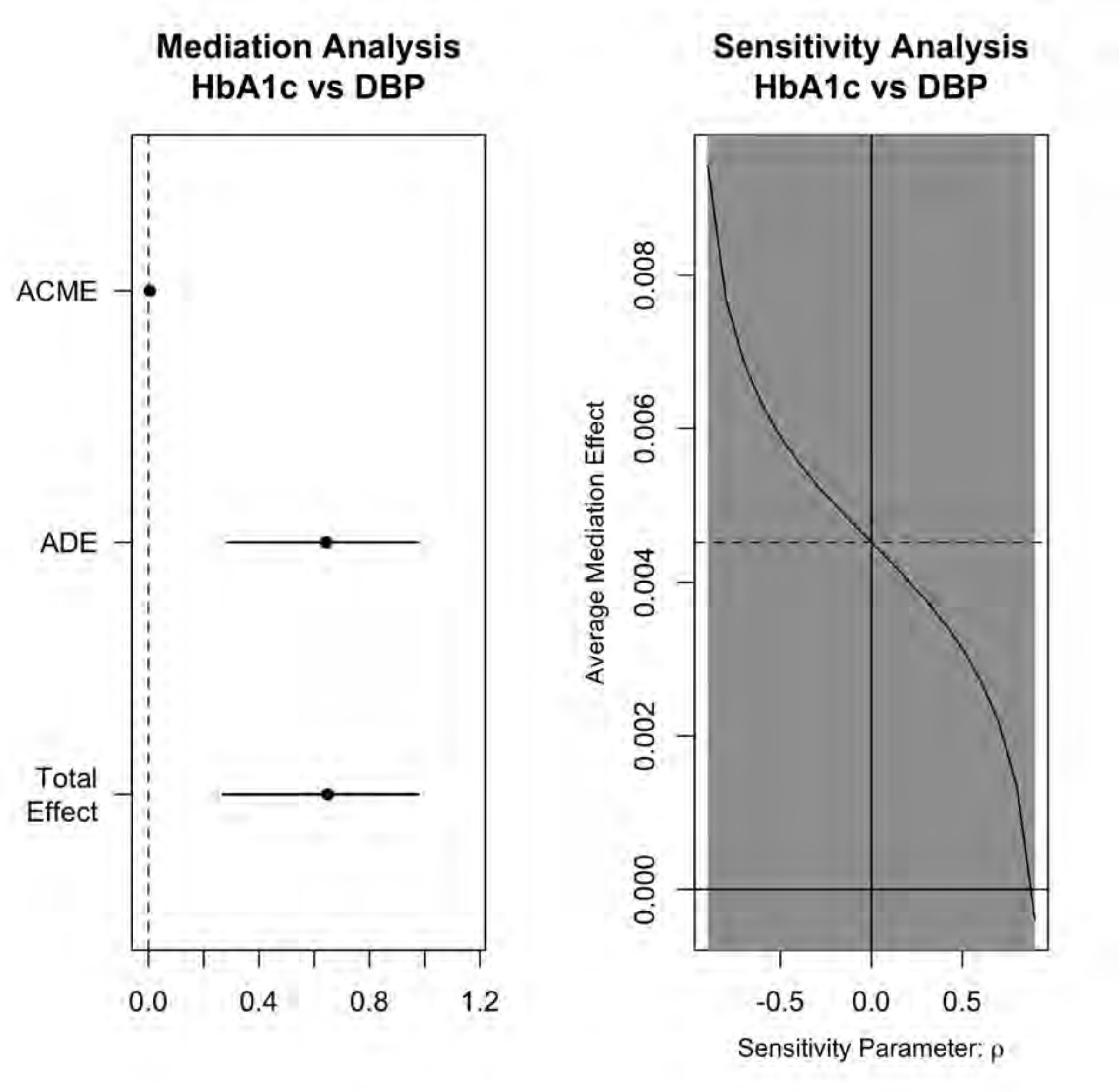

***Supplementary Figure 21. Bayes model of the mediation effects of GLP1a on diastolic blood pressure***

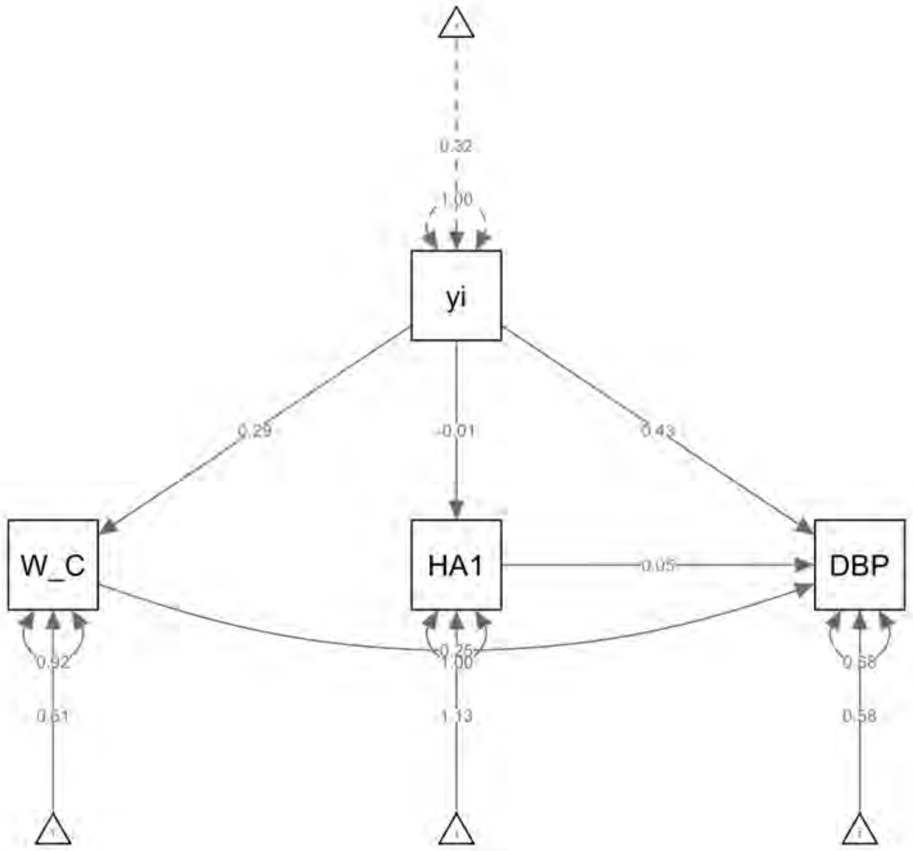

#### Supplementary Result 7: Subgroup analysis by agonist status

***Supplementary Figure 22. Forest plot of the network analysis of systolic blood pressure stratified by single, dual or triple agonist***

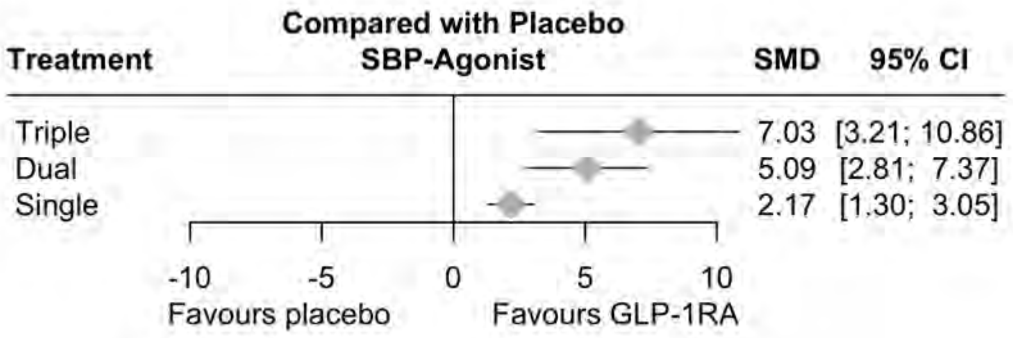

***Supplementary Figure 23. Netgraph of the network analysis of systolic blood pressure stratified by single, dual or triple agonist***

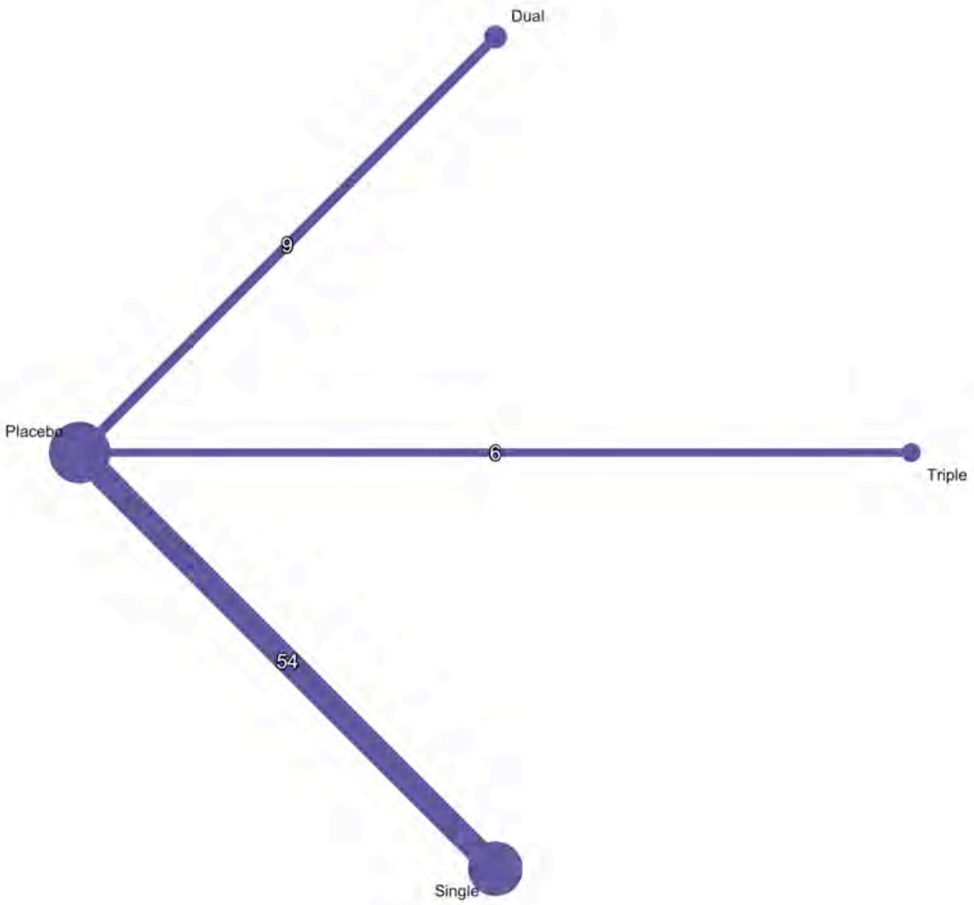

*Supplementary Figure 24. League table of the network analysis of systolic blood pressure stratified by single, dual or triple agonist*

| | | | |
|---|---|---|---|
| Triple | | | |
| 1.95 ( -2.51, 6.40) | Dual | | |
| 4.86 ( 0.94, 8.78) | 2.91 ( 0.47, 5.36) | Single | |
| 7.03 ( 3.21, 10.86) | 5.09 ( 2.81, 7.37) | 2.17 ( 1.30, 3.05) | Placebo |

***SSupplementary Figure 25. Forest plot of the network analysis of diastolic blood pressure stratified by single, dual or triple agonist***

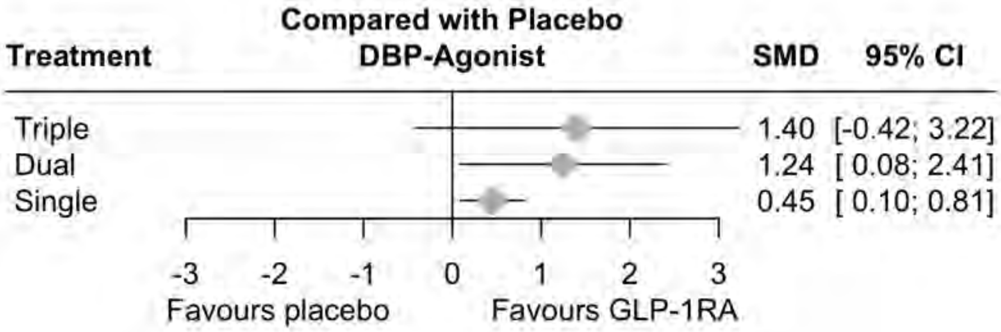

***Supplementary Figure 26. Netgraph of the network analysis of diastolic blood pressure stratified by single, dual or triple agonist***

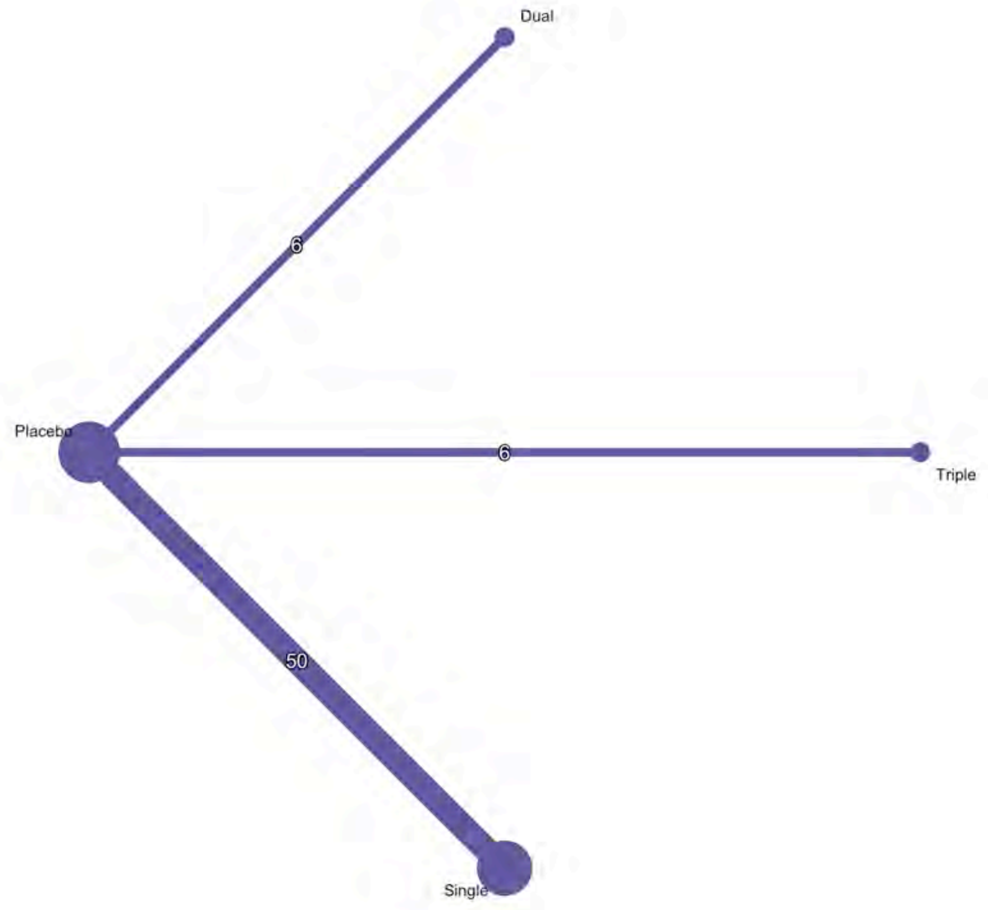

***Supplementary Figure 27. League table of the network analysis of diastolic blood pressure stratified by single, dual or triple agonist***

| | | | |
|---|---|---|---|
| **Triple** | | | |
| 0.16 (-2.00, 2.32) | **Dual** | | |
| 0.95 (-0.91, 2.80) | 0.79 (-0.42, 2.01) | **Single** | |
| 1.40 (-0.42, 3.22) | 1.24 ( 0.08, 2.41) | 0.45 ( 0.10, 0.81) | **Placebo** |

Supplementary Figure . League table of the network analysis of diastolic blood pressure stratified by single, dual or triple agonist

***Supplementary Figure 28. Forest plot of the network analysis of weight loss stratified by single, dual or triple agonist***

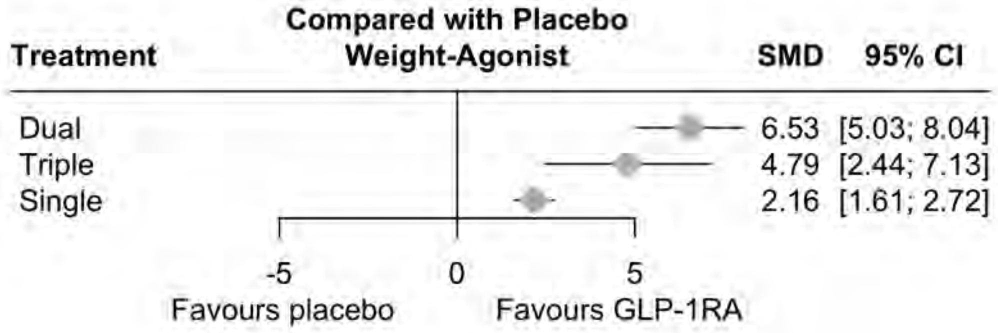

***Supplementary Figure 29. Netgraph of the network analysis of weight loss stratified by single, dual or triple agonist***

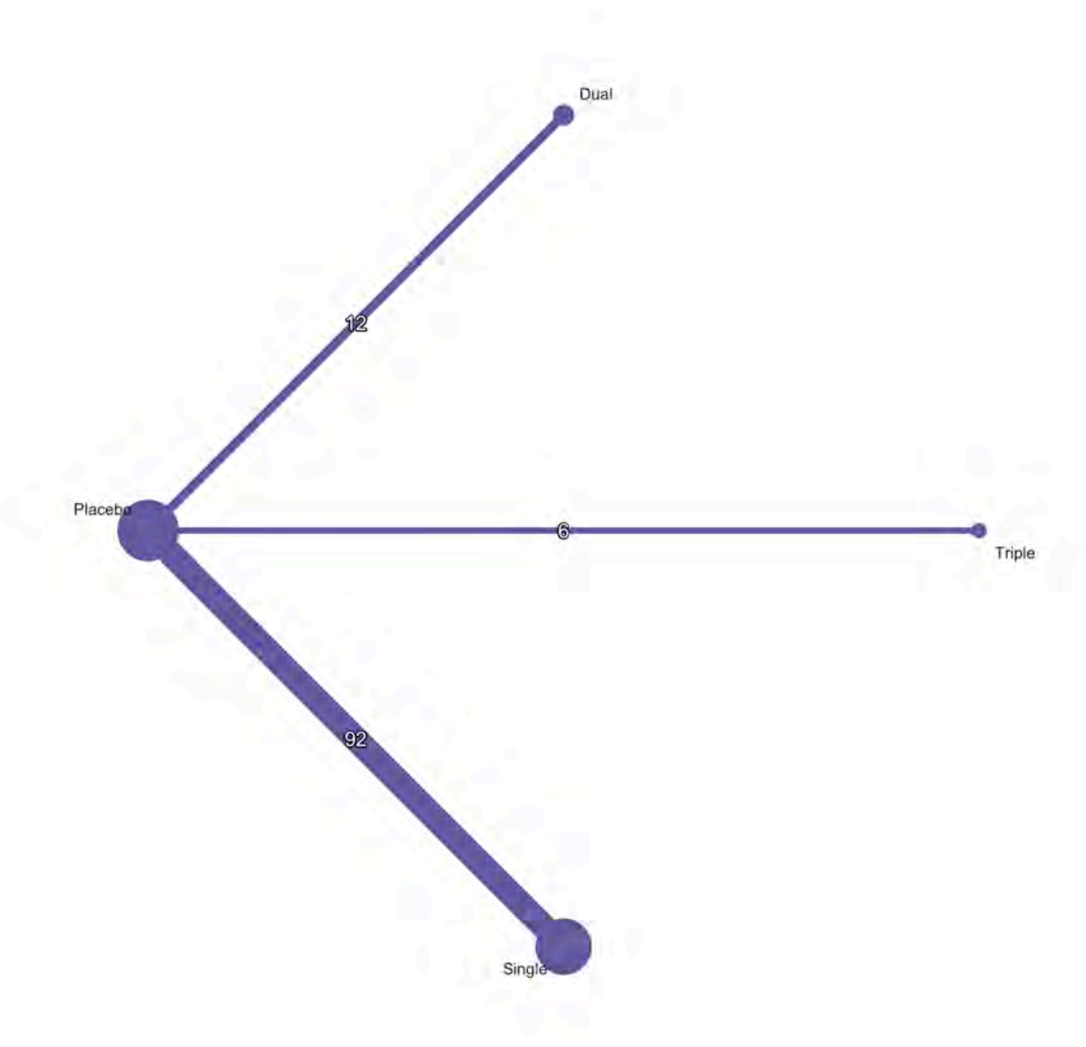

***Supplementary Figure 30. League table of the network analysis of weight loss stratified by single, dual or triple agonist***

| | | | |
|---|---|---|---|
| Dual | | | |
| 1.75 (-1.04, 4.53) | Triple | | |
| 4.37 ( 2.77, 5.97) | 2.62 ( 0.22, 5.03) | Single | |
| 6.53 ( 5.03, 8.04) | 4.79 ( 2.44, 7.13) | 2.16 ( 1.61, 2.72) | Placebo |

*Supplementary Figure 31. Forest plot of the network analysis of HbA1c stratified by single, dual or triple agonist*

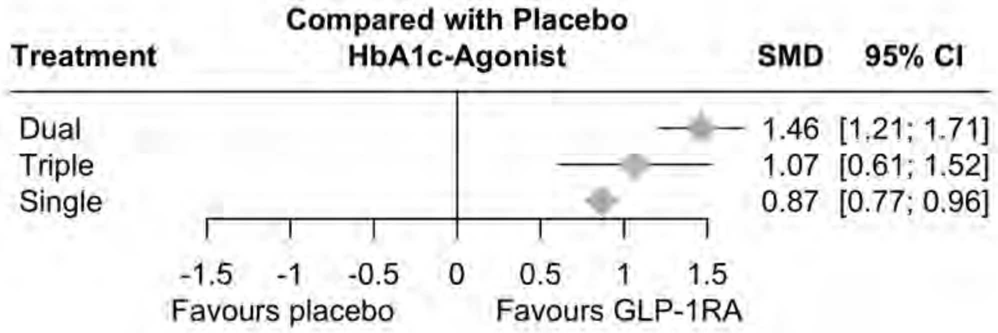

***Supplementary Figure 32. Netgraph of the network analysis of HbA1c stratified by single, dual or triple agonist***

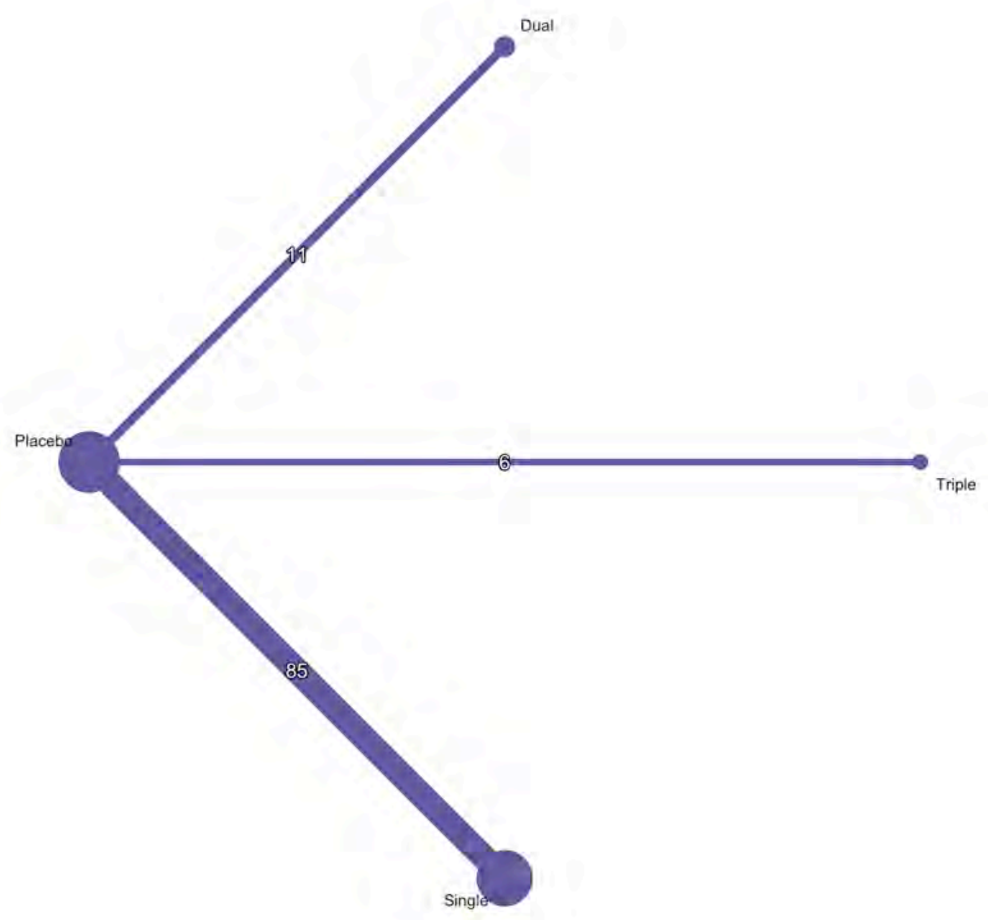

*Supplementary Figure 33. League table of the network analysis of HbA1c stratified by single, dual or triple agonist*

| | | | |
|---|---|---|---|
| Dual | | | |
| 0.40 (-0.12, 0.92) | Triple | | |
| 0.60 ( 0.33, 0.86) | 0.20 (-0.27, 0.67) | Single | |
| 1.46 ( 1.21, 1.71) | 1.07 ( 0.61, 1.52) | 0.87 ( 0.77, 0.96) | Placebo |

### Supplementary Result 8: Subgroup analysis by different dosage of GLP1A

***Supplementary Figure 34. Forest plot of the network analysis of GLP1A effects on systolic blood pressure stratified by different dosage***

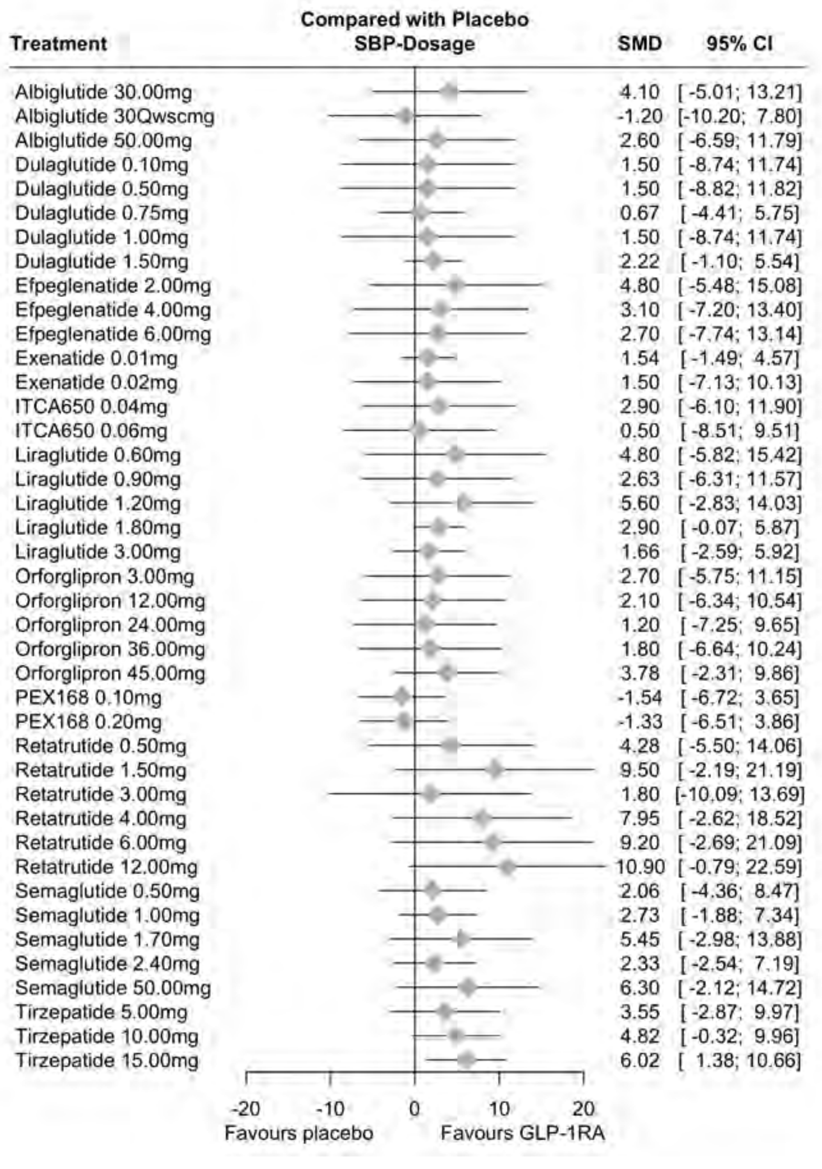

***Supplementary Figure 35. Netgraph of the network analysis of GLP1A effects on systolic blood pressure stratified by different dosage***

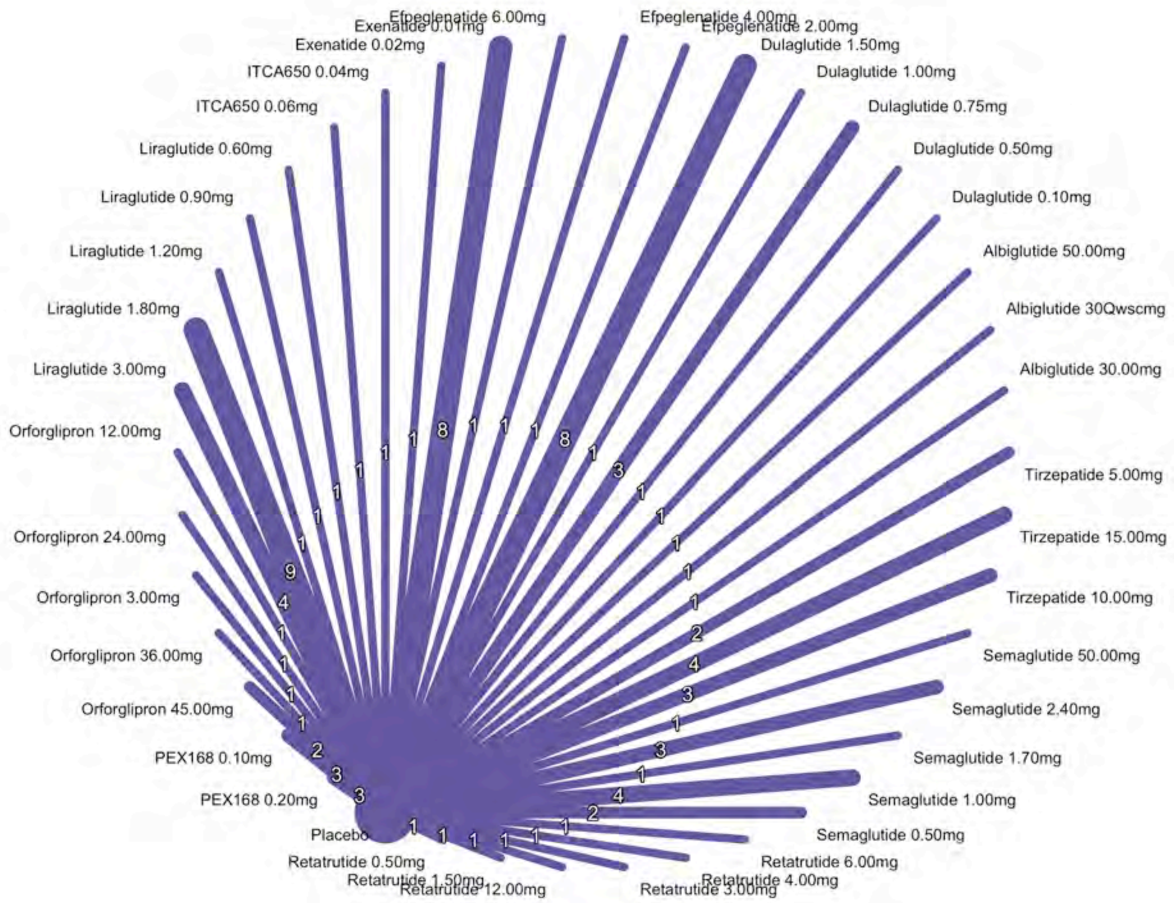

***Supplementary Figure 36. Forest plot of the network analysis of GLP1A effects on diastolic blood pressure stratified by different dosage***

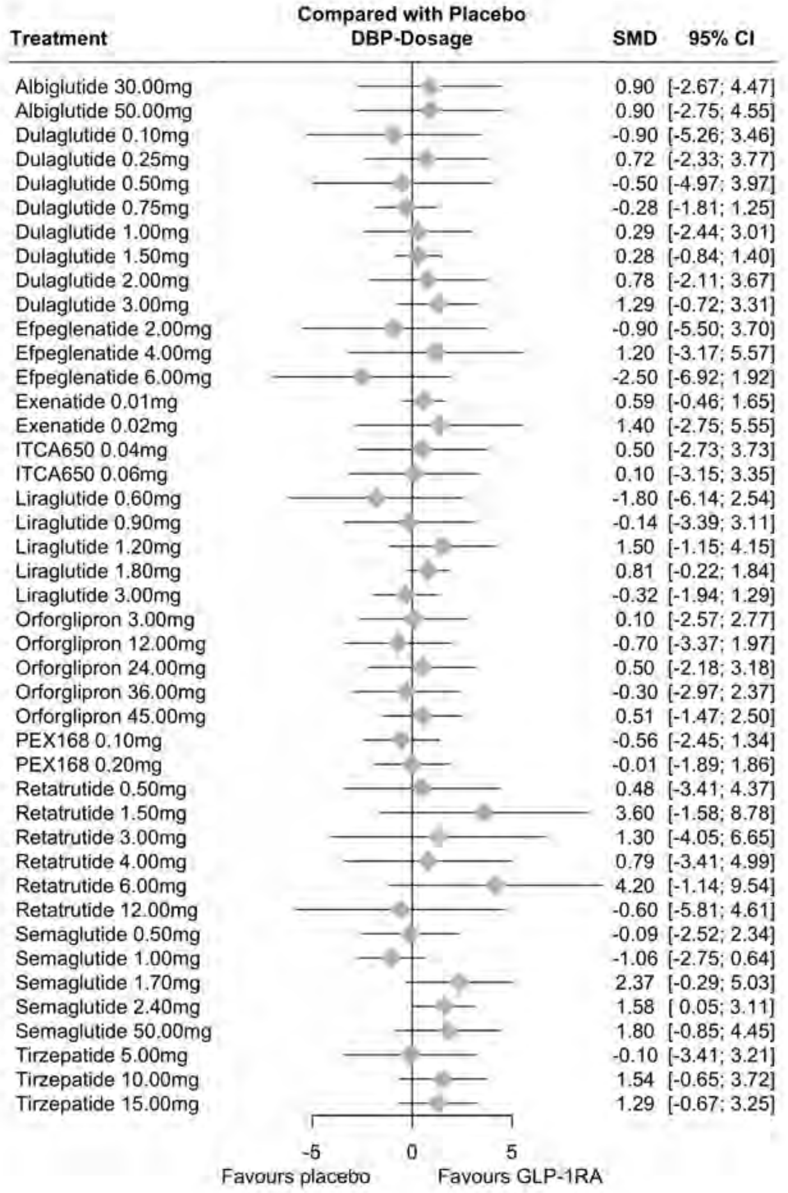

***Supplementary Figure 37. Netgraph of the network analysis of GLP1A effects on diastolic blood pressure stratified by different dosage***

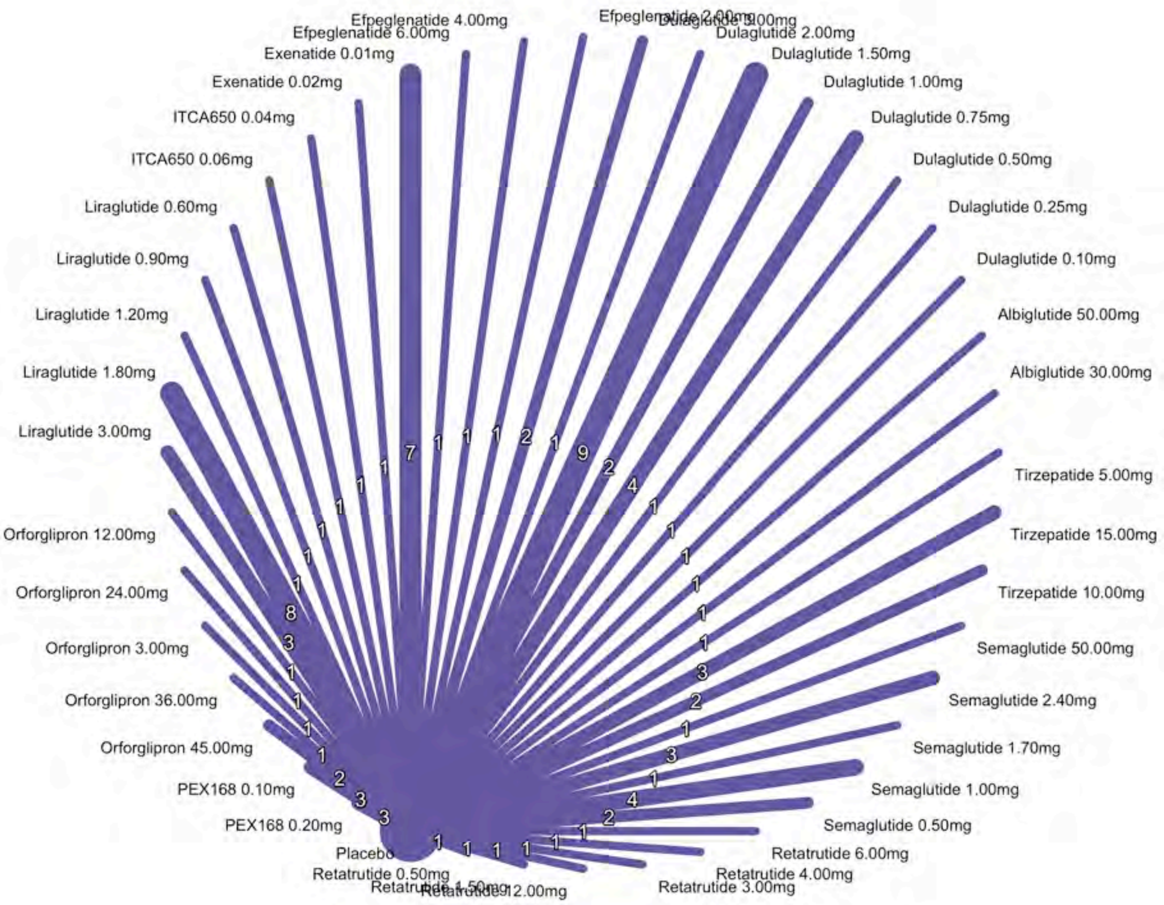

***Supplementary Figure 38. Forest plot of the network analysis of GLP1A effects on weight loss stratified by different dosage***

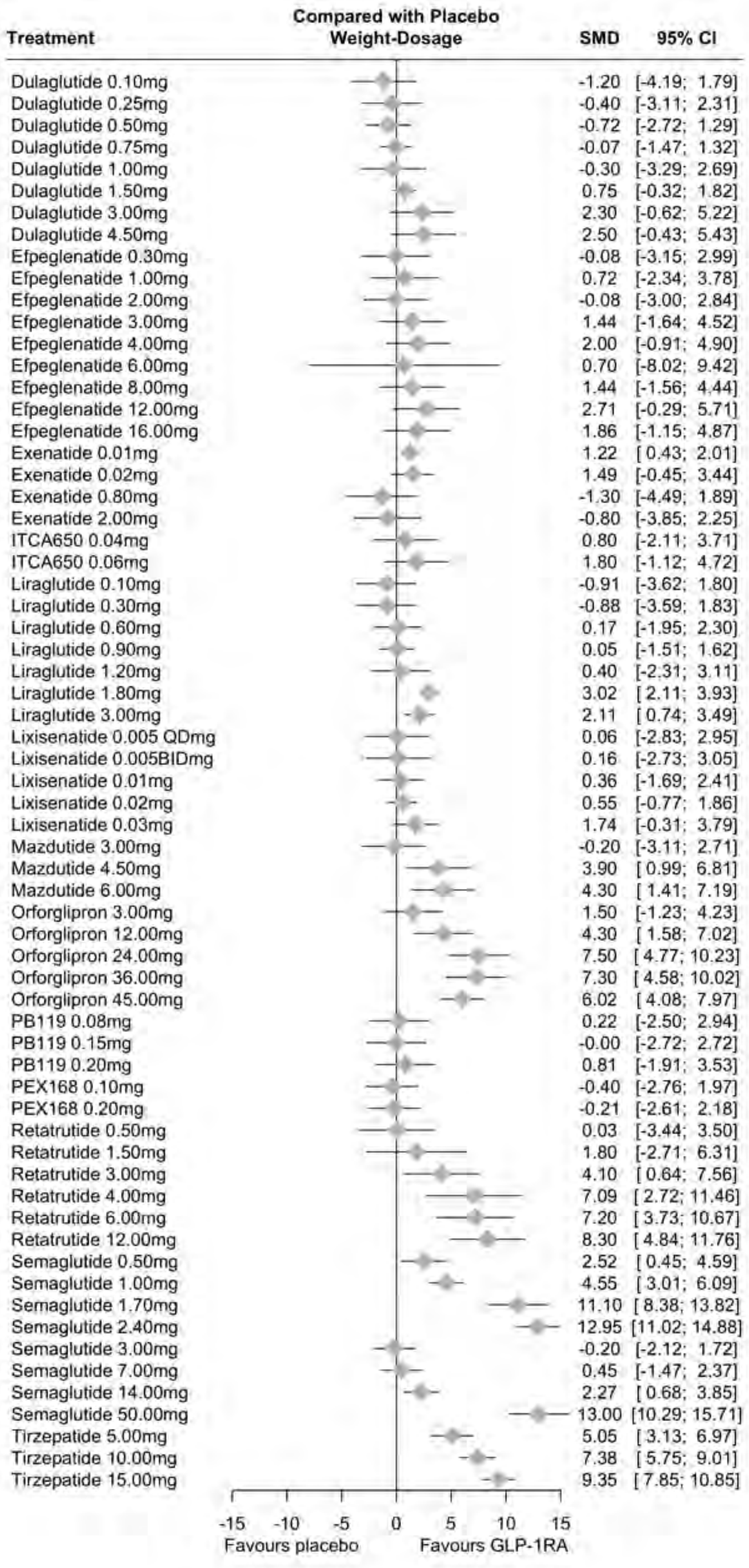

***Supplementary Figure 39. Netgraph of the network analysis of GLP1A effects on weight loss stratified by different dosage***

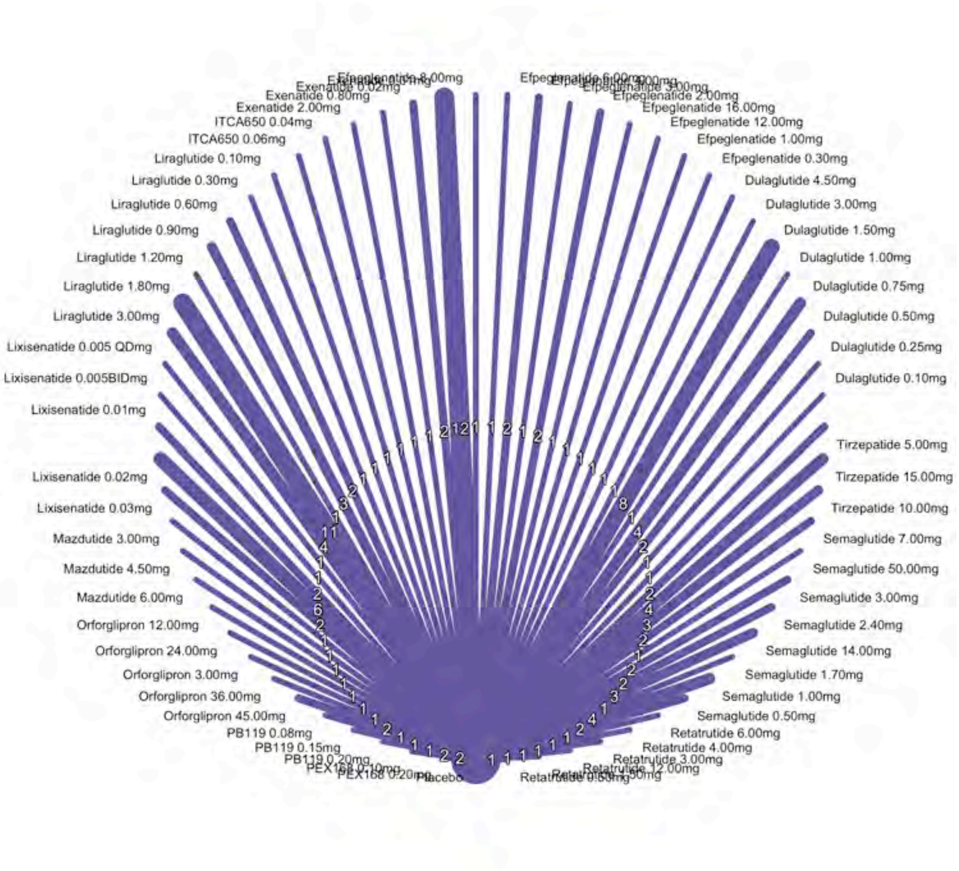

***Supplementary Figure 40. Forest plot of the network analysis of GLP1A effects on HbA1c stratified by different dosage***

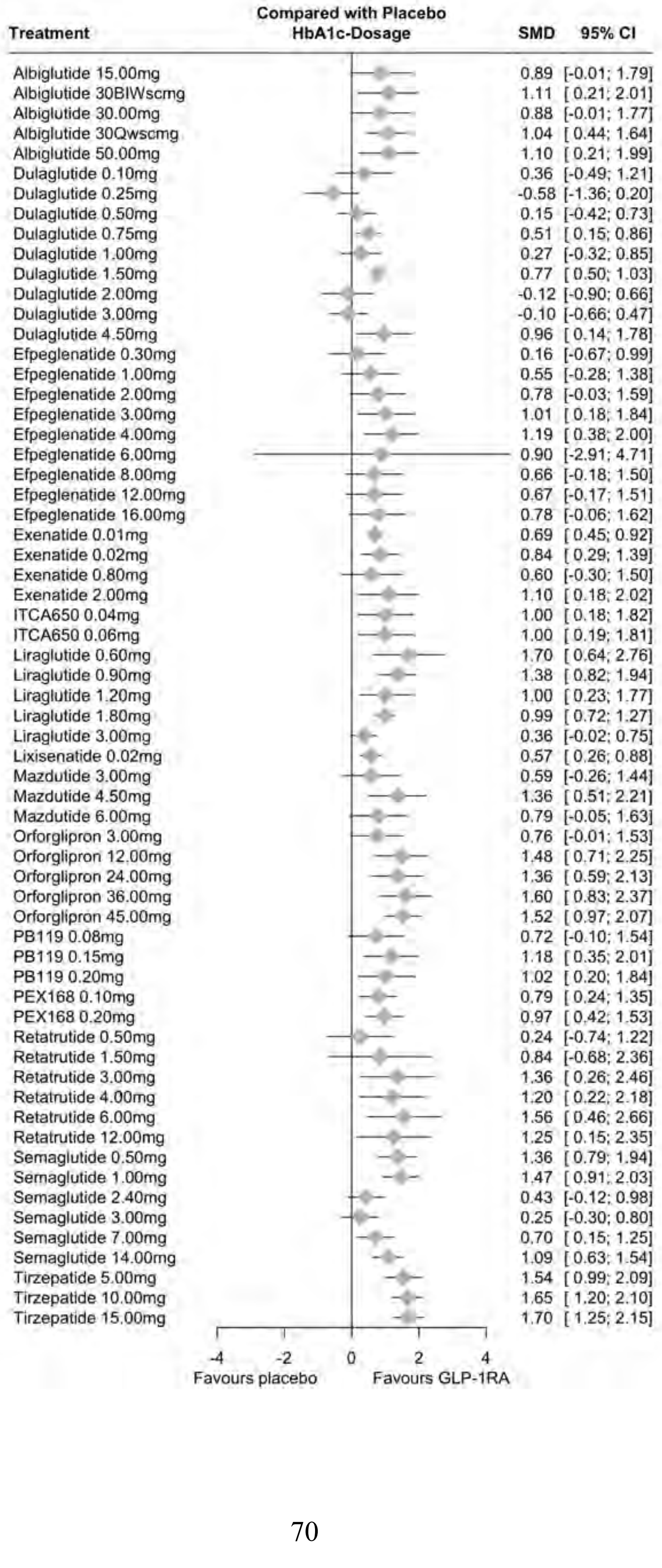

***Supplementary Figure 41. Netgraph of the network analysis of GLP1A effects on HbA1c stratified by different dosage***

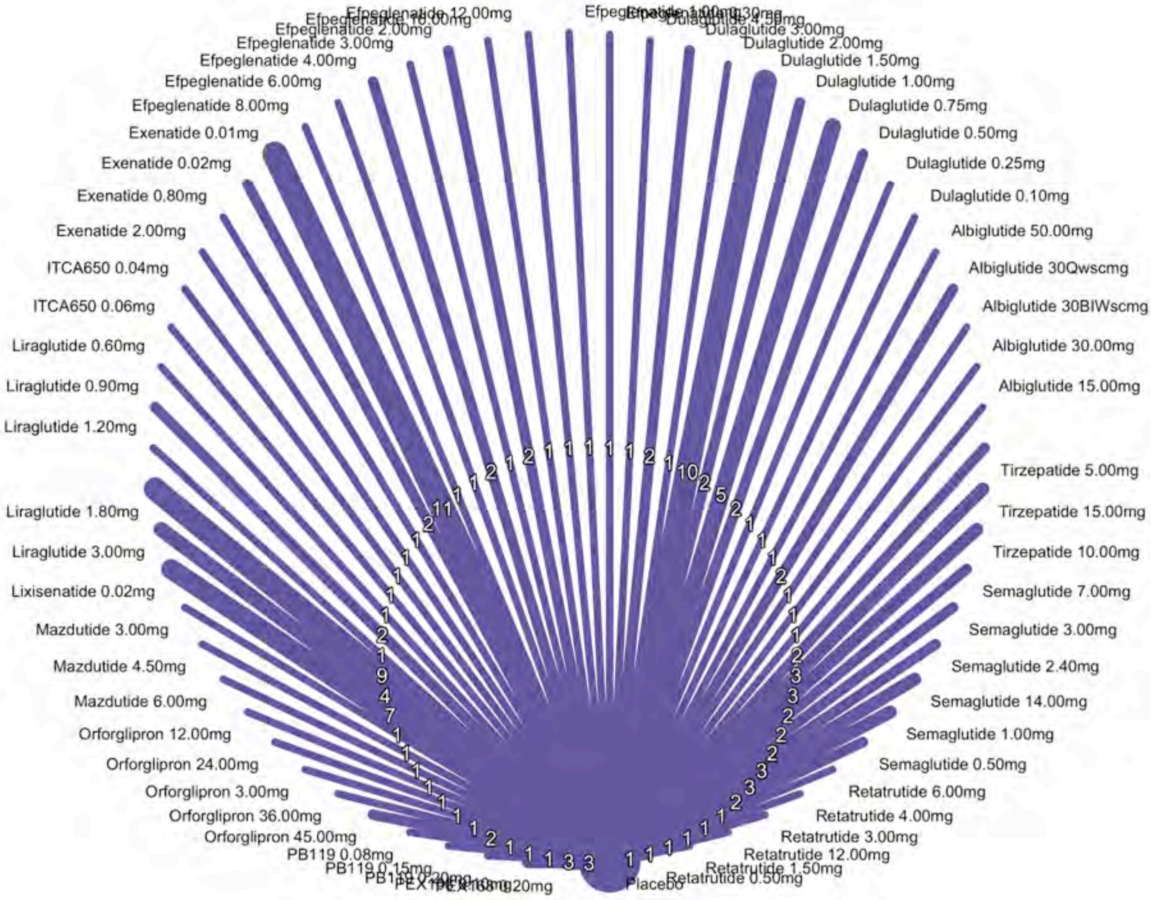

#### Supplementary Result 9: Subgroup analysis of selected drugs

***Supplementary Figure 42. Forest plot of the network analysis of Tirzepatide effects on systolic blood pressure stratified by different dosage***

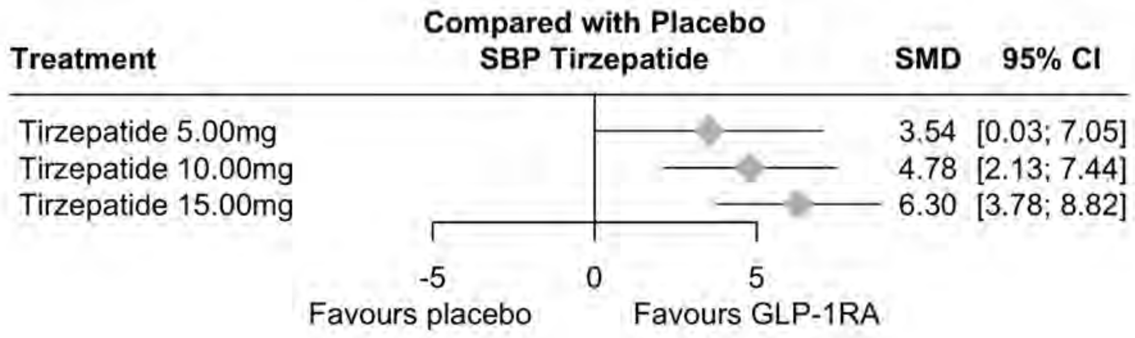

***Supplementary Figure 43. Netgraph of the network analysis of Tirzepatide effects on systolic blood pressure stratified by different dosage***

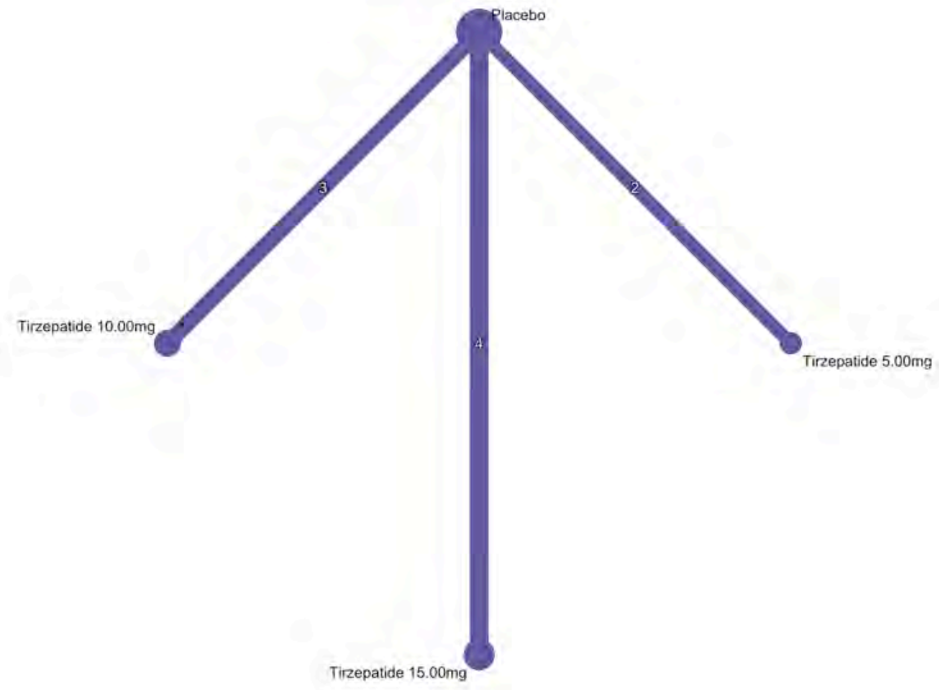

***Supplementary Figure 44. Forest plot of the network analysis of Tirzepatide effects on diastolic blood pressure stratified by different dosage***

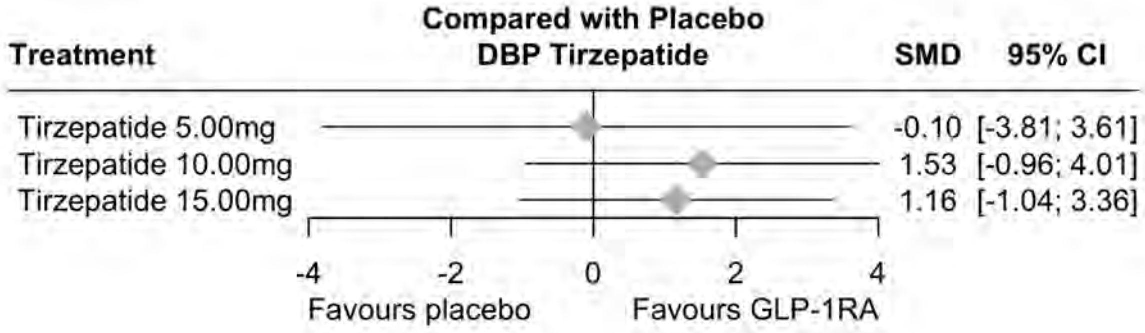

***Supplementary Figure 45. Netgraph of the network analysis of Tirzepatide effects on diastolic blood pressure stratified by different dosage***

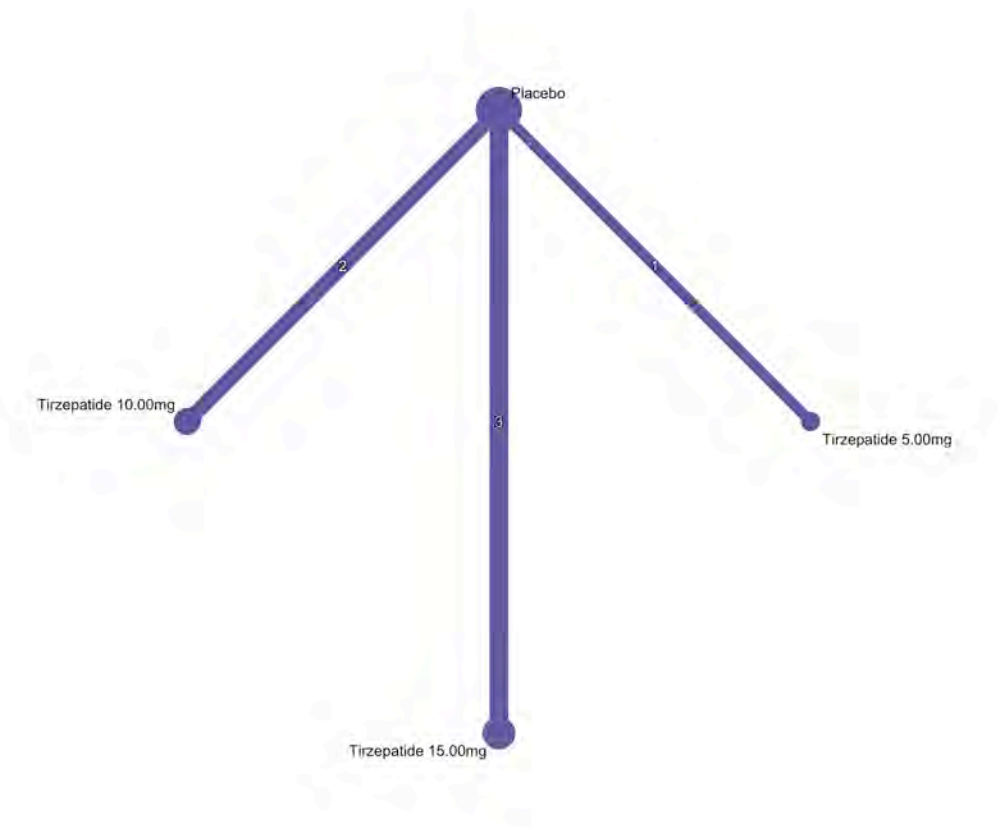

*Supplementary Figure 46. Mean age plot on blood pressure*

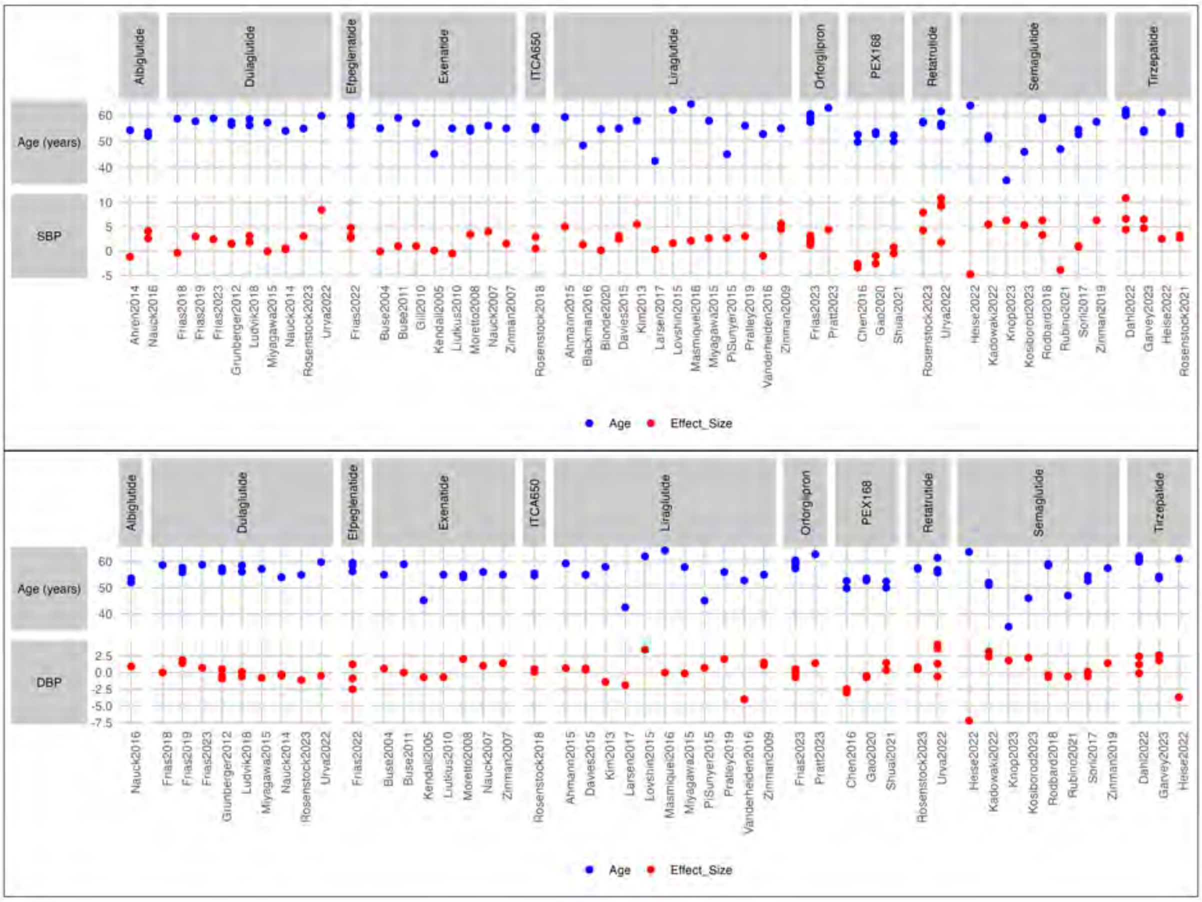
